# The consequences of a year of the COVID-19 pandemic for the mental health of young adult twins in England and Wales

**DOI:** 10.1101/2021.10.07.21264655

**Authors:** Kaili Rimfeld, Margherita Malanchini, Ryan Arathimos, Agnieszka Gidziela, Oliver Pain, Andrew McMillan, Rachel Ogden, Louise Webster, Amy E. Packer, Nicholas G. Shakeshaft, Kerry L. Schofield, Jean-Baptiste Pingault, Andrea G. Allegrini, Argyris Stringaris, Sophie von Stumm, Cathryn M. Lewis, Robert Plomin

## Abstract

The COVID-19 pandemic has impacted all our lives, not only through the infection itself, but also through the measures taken to control the virus’s spread (e.g., lockdown). Here we investigated how the COVID-19 pandemic and unprecedented lockdown affected the mental health of young adults in England and Wales. We compared the mental health symptoms of up to 4,000 twins in their mid-twenties in 2018 prior to the COVID-19 pandemic (T1) to those in a four-wave longitudinal data collection during the pandemic in April, July, and October 2020, and in March 2021 (T2-T5). The average changes in mental health were small-to-medium and mainly occurred from 2018 (T1) to March 2020 (T2, one month following the start of lockdown; average Cohen d=0.14). Despite the expectation of catastrophic effects on the pandemic on mental health of our young adults, we did not observe trends in worsening mental health during the pandemic (T3-T5). Young people with pre-existing mental health problems were adversely affected at the beginning of the pandemic, but their increased problems largely subsided as the pandemic persisted. Twin analyses indicated that the aetiology of individual differences did not change during the lockdown. The average heritability of mental health symptoms was 33% across 5 waves of assessment, and the average genetic correlation between T1 and T2-T5 was .95, indicating that genetic effects before the pandemic (T1) are substantially correlated with genetic effects up to a year later (T2-T5). We conclude that on average the mental health of young adults in England and Wales has been remarkably resilient to the effects of the pandemic and associated lockdown.

## Introduction

The COVID-19 pandemic has affected both those infected by the virus as well as those spared from the infection who had to adhere to strict lockdowns. The COVID-19 pandemic has changed the everyday lives of all, through full or partial forced isolation (lockdown), closure of schools and public spaces, and associated economic consequences. Several studies have reported that the pandemic negatively affected mental health, which seems reasonable given the scale of the crisis^1–4^. This pandemic might worsen mental health on average in the population, but young people are likely to be affected disproportionately more^5–8^. Indeed, young people’s mental health has been reported to be worse following the pandemic-related lockdown compared to the adult population even after the first restrictions were lifting^9^.

Young adulthood (the period between early twenties and early thirties) is a time marked by instability and fragility in several aspects of life, and is a critical age for the development of psychopathologies^10–12^. Even before COVID-19, Labour Force Survey of the Office on National Statistics reported that 17.9M days of work are lost every year to stress, depression and anxiety^13^. The ∼12M young adults living in the UK make up over a third of the current workforce and will be nearly half of the workforce within the next two decades. The instability and financial hardship generated by the COVID-19 pandemic are likely to exacerbate the struggles of young adults, with cascading effects on the health, wellbeing, and economic stability of the nation.

Although several studies have reported that the pandemic negatively affects mental health, the extent of these effects remains an open question because most reports focused on statistically significant average differences rather than effect size. When reported, the effect sizes are typically small to medium^14^, and some studies find little change in mental health following the COVID-19 pandemic and lockdown^15–18^. Some reports suggest that population levels of anxiety and depression were elevated immediately after lockdown, but then decreased again as the pandemic continued^14,16^. There is also some evidence for improvement in mental health symptoms during the pandemic^19,20^. For example, some people seemed to thrive during the pandemic and lockdown, reporting increased wellbeing^21,22^. These seemingly inconsistent findings might be due to methodological differences between the studies, such as differences in age, measures, timing of the lockdown, and whether pre-pandemic measures were available.

Notably, the majority of the research to date focuses on average psychological changes during the COVID-19 crisis and the experience of lockdown, but this crisis is likely to affect individuals differently^23^. Individual differences are likely to include both negative changes such as increased anxiety and depression, and positive changes such as increased wellbeing^22^, which could cancel out average changes. For this reason, it is important to study individual differences as well as average differences (means) in response to the pandemic and the experience of lockdown.

A fundamental aspect of individual differences that is often ignored when studying the response to the COVID-19 crisis is genetic variation. Yet, it has been demonstrated over decades of twin studies that inherited DNA differences contribute substantially to most psychological traits^24,25^. Moreover, research has shown that sensitivity to environmental changes has a heritable component^26^. It is therefore reasonable to assume that individual differences in the response to the COVID-19 crisis are also partially driven by genetic variation between individuals.

Our first study^27^, using the Twins Early Development Study (TEDS) sample, compared a wide range of psychological measures (30 traits) from T1 (2018) to T2 (April 2020) and found modest and unsystematic mean changes in these traits after one month of the COVID-19 lockdown. The largest negative effects of the pandemic were reduced volunteering (d=0.84), decreased achievement motivation (d=0.47) and increased hyperactivity-inattention (d=0.42). However, we also observed many positive changes in response to the pandemic, most notably reduced peer victimisation, reduced alcohol (quantity) consumption and decreased self-harming. It is, however, possible that one month of lockdown was an insufficient time frame to examine the negative consequences of this unprecedented restrictions. Furthermore, the lockdowns did not end after one month and the restrictions continued until March 2021; it is therefore important to study the longer-term effects of COVID-19 pandemic on mental health.

Our first study also examined the aetiology of mental health symptoms before and one month after the lockdown. We found that genetic factors accounted for around half of the reliable variance in diverse psychological traits both at T1 and T2. Furthermore, we found that genetic factors at T1 are highly correlated with genetic factors at T2. We also investigated possible moderators, for example, family socioeconomic factors, living conditions, financial difficulties, but found no evidence of gene-environment interaction. Here, we investigate the extent to which the aetiology of mental health symptoms changes over a longer period, from T1 to T5.

We use longitudinal data collected over five waves (2018 and four pandemic waves of assessment one, four, seven, and eleven months after the first lockdown in Britain began in March 2020). The sample consisted of twins in TEDS^17^ assessed when the twins were in their mid-twenties. Our focus is on tracking individual differences in mental health trajectories over the course of the crisis compared to 2018 using diverse measures of mental health symptoms, including conduct problems, emotional problems, hyperactivity, peer problems, prosocial behaviour, general anxiety, depression, and self-harm. The genetic and environmental origins of these measures were assessed by the classical twin method that compares the resemblance of identical and non-identical twins. It is reasonable to expect that a major environmental shift, such as the COVID-19 pandemic, reshuffles individual differences, for example, the genetic effects could be either reduced or amplified, alternatively this environmental shift could evoke innovative genetic effects^28–30^. We also investigated factors influencing changes in mental health using genome-wide polygenic scores, aggregated scores that capture genetic predisposition towards risk and protective factors based on previous genome-wide association studies (GWAS)^31^, which allowed us to establish if individuals with higher risk on psychiatric diseases are disproportionately affected by the COVID-19 pandemic. We also investigated the extent to which family socioeconomic status, lockdown conditions, life changes and home environment affect response to the pandemic.

Several studies have indicated that lockdown measures could be especially detrimental for young adults with existing psychological and psychiatric vulnerabilities^8,22,32^. Here we capitalised on data collected in TEDS in T1 (2018) to identify young adults who experienced mental health problems prior to the COVID-19 pandemic. We assessed the extent to which these individuals are disproportionately affected by the pandemic and associated lockdown. We also addressed this issue using polygenic scores as indicators of genetic vulnerabilities.

Our hypotheses and plan for analyses were preregistered in the Open Science Framework (https://osf.io/gzrk3) prior to accessing the data. Our main hypotheses were that mean changes from T1 to T5 will be significant, with small to modest effect sizes for the entire sample but stronger in vulnerable groups. Heritability was predicted to be substantial at each assessment from T1 to T5. We also hypothesised that mental health changes from T1 to T5 will show significant but modest genetic influence, with most of the changes environmental in origin. Polygenic scores were predicted to yield a similar pattern of results as the twin analyses.

## Methods

### Sample

Our sample consists of young adults enrolled in the Twins Early Development Study (TEDS)^33^. TEDS is a twin study that recruited twins born between 1994 and 1996 in England and Wales, as identified through birth records. The invitations were sent to families by the UK Office for National Statistics after screening for infant mortality, and 16,810 families expressed interest in taking part. TEDS conducted the first wave of data collection when twins were around 18 months old, obtaining demographic information, data about pregnancy and childbirth, and questions related to zygosity. The sample was, and remains, reasonably representative of the population in England and Wales in terms of ethnicity and socio-economic factors for this birth cohort^33^.

The current study uses data from the TEDS data collection wave conducted when the youngest twins were aged 21 (completed in 2018), together with COVID-19 data collection completed in April, July and October 2020, and March 2021. The sample size includes up to 4,773 unrelated individuals (one randomly selected twin per pair), up to 1,501 complete MZ twin pairs and up to 2,380 complete DZ twin pairs; these sample sizes vary across measures and measurement occasions (see Supplementary Table S1). All available data were used.

### Measures

In 2018, when the twins were aged 21-25 years (Mean age=22.27, SD=0.9), they completed a battery of mental health measures, which are listed in Table 1. These serve as a baseline to examine how the COVID-19 crisis has changed the mental health of young adults during the four subsequent waves of assessment, when the twins completed the same battery of mental health measures: April 2020 (one month after the first COVID-19 lockdown began), July 2020, October 2020 and March 2021. In addition, the **C**o**R**onav**I**ru**S** Health **I**mpact **S**urvey (CRISIS)^34^ was administered at T2-T5. CRISIS assesses mental health, behaviours, home environment, physical health and life changes (see Figure 1).

**Table 1.**
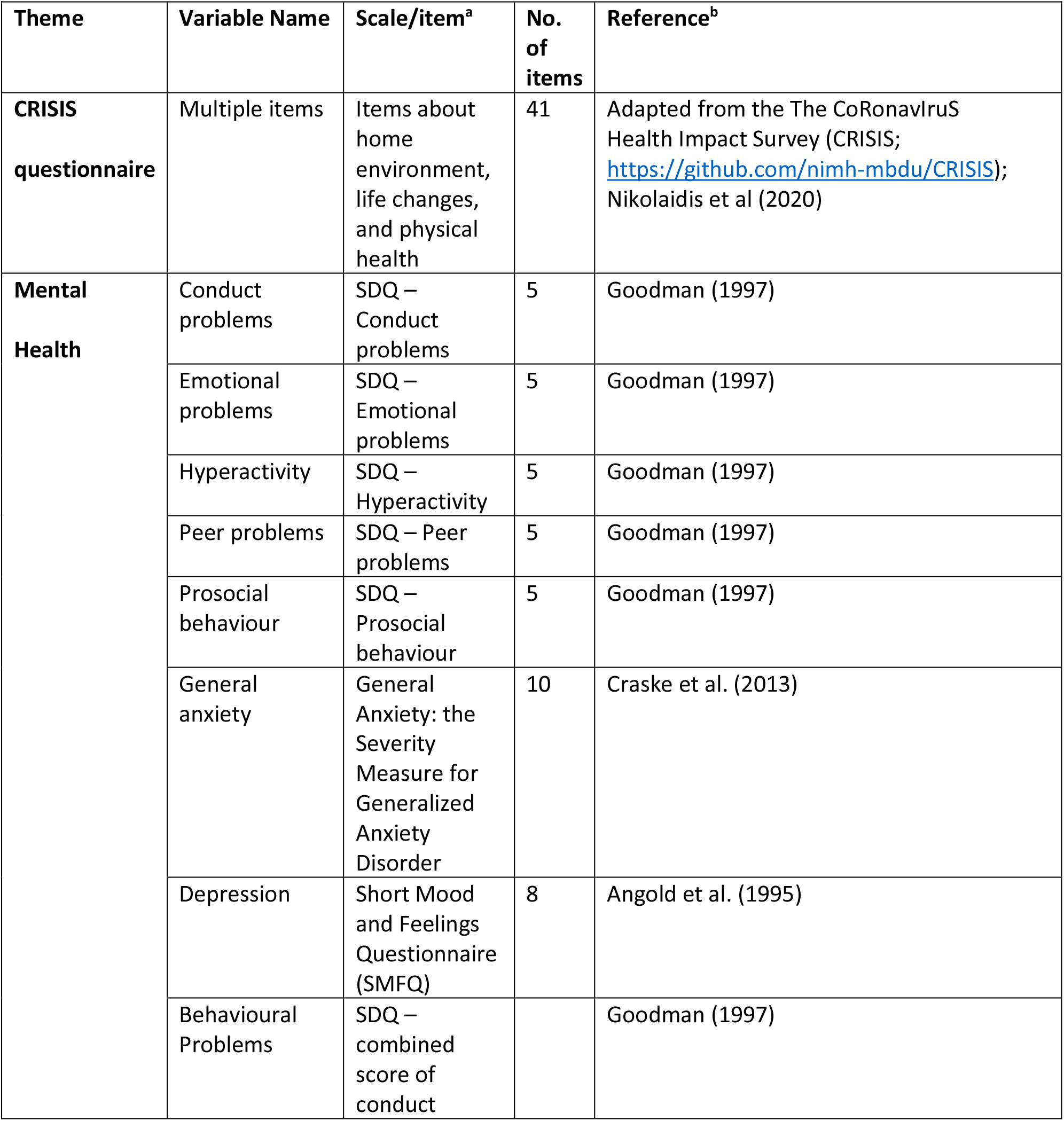
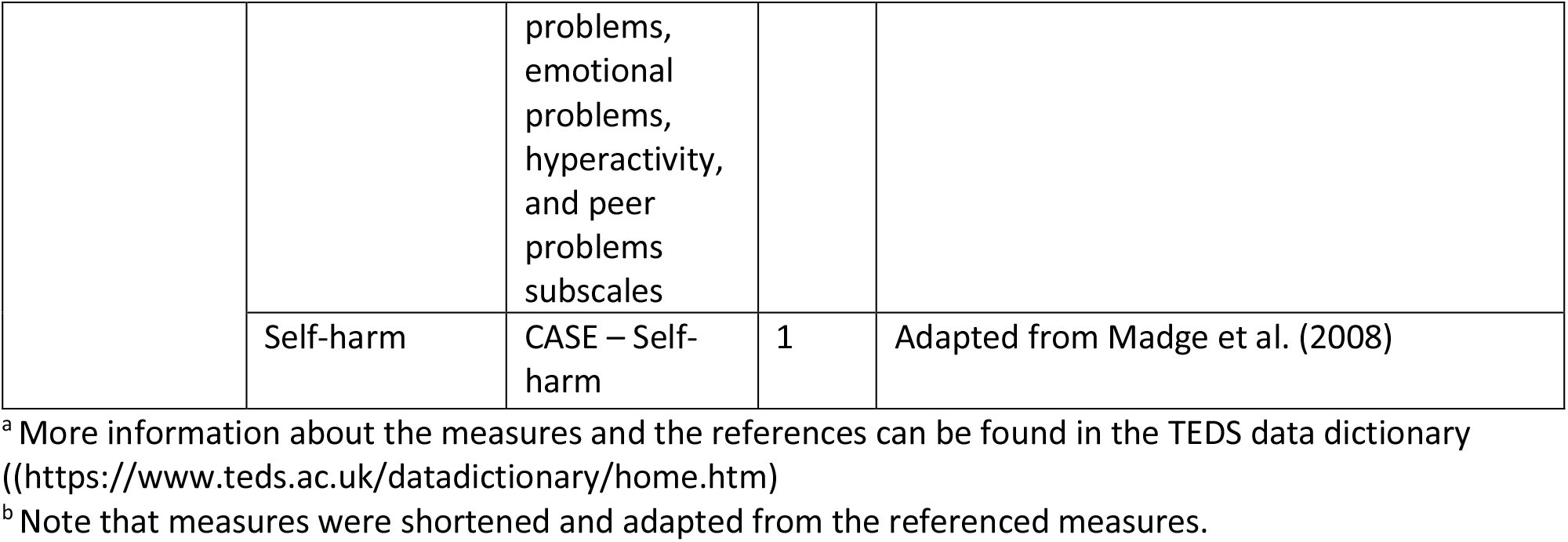
Measured variables

| Theme | Variable Name | Scale/item<sup>a</sup> | No. of items | Reference<sup>b</sup> |
| --- | --- | --- | --- | --- |
| **CRISIS questionnaire** | Multiple items | Items about home environment, life changes, and physical health | 41 | Adapted from the The CoRonaviruS Health Impact Survey (CRISIS; https://github.com/nimh-mbdu/CRISIS); Nikolaidis et al (2020) |
| **Mental Health** | Conduct problems | SDQ – Conduct problems | 5 | Goodman (1997) |
|  | Emotional problems | SDQ – Emotional problems | 5 | Goodman (1997) |
|  | Hyperactivity | SDQ – Hyperactivity | 5 | Goodman (1997) |
|  | Peer problems | SDQ – Peer problems | 5 | Goodman (1997) |
|  | Prosocial behaviour | SDQ – Prosocial behaviour | 5 | Goodman (1997) |
|  | General anxiety | General Anxiety: the Severity Measure for Generalized Anxiety Disorder | 10 | Craske et al. (2013) |
|  | Depression | Short Mood and Feelings Questionnaire (SMFQ) | 8 | Angold et al. (1995) |
|  | Behavioural Problems | SDQ – combined score of conduct problems, emotional problems, hyperactivity, and peer problems subscales |  | Goodman (1997) |
|  | Self-harm | CASE – Self-harm | 1 | Adapted from Madge et al. (2008) |

<sup>a</sup> More information about the measures and the references can be found in the TEDS data dictionary (https://www.teds.ac.uk/datadictionary/home.htm)
<sup>b</sup> Note that measures were shortened and adapted from the referenced measures.

**Figure 1.**
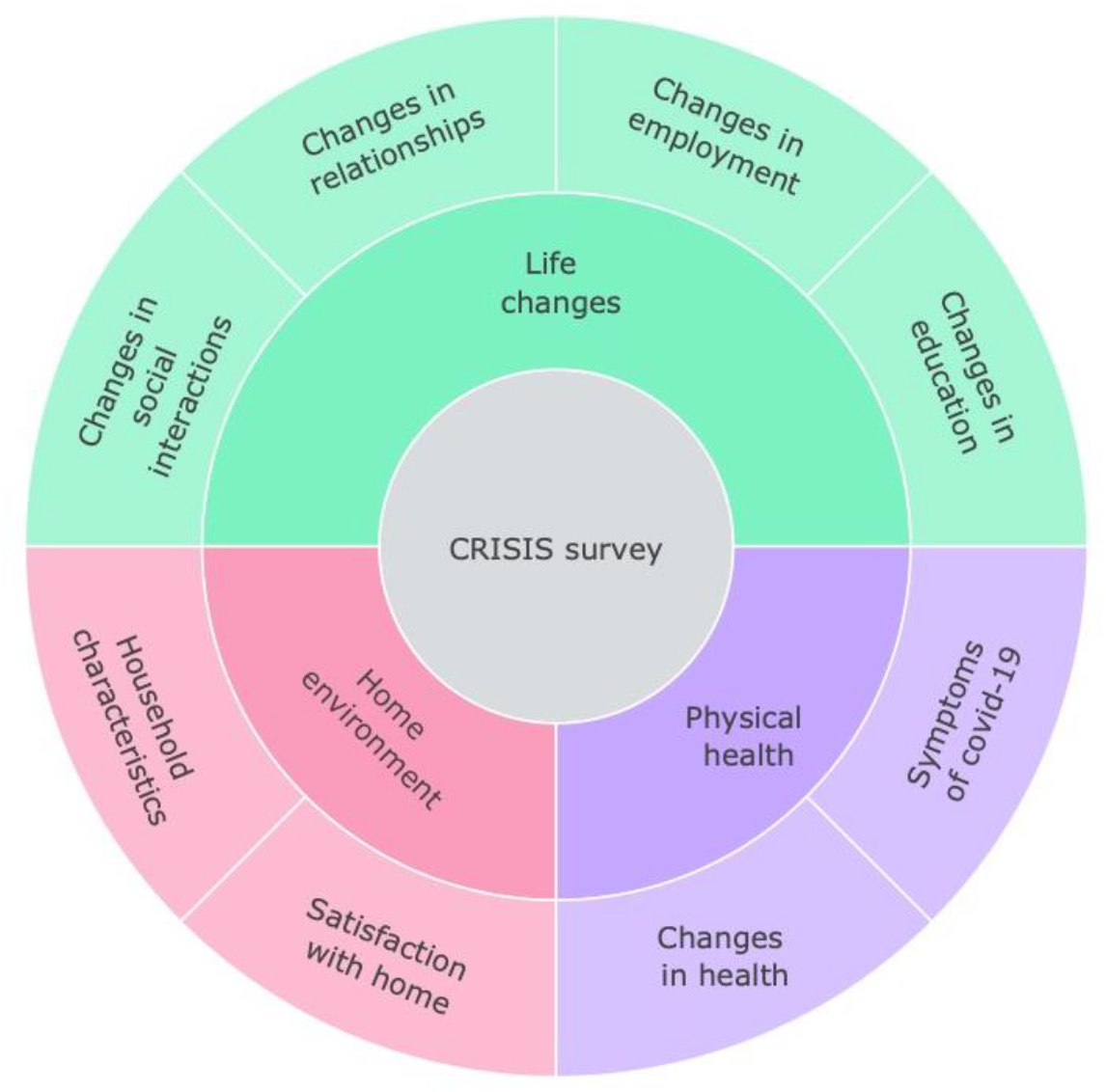
Summary of the constructs measured by the CRISIS questionnaire.

All data were collected using online questionnaires or there was also an option to contribute to the data collection via the paper and pencil questionnaires (data collection in 2018 only). More information is available on TEDS Data Dictionary (https://www.teds.ac.uk/datadictionary/home.htm).

### Genotyping

DNA samples have been obtained from 12,500 individuals and genotyped on one of two DNA microarrays (Affymetrix GeneChip 6.0 or Illumina HumanOmniExpressExome chips). After stringent quality control, the total sample size available for genomic analyses was 10,346 (including 7,026 unrelated individuals and 3,320 additional DZ co-twins). Of these, 7,289 individuals were genotyped on Illumina arrays, and 3,057 individuals were genotyped on Affymetrix arrays^33^ (see^35^ for a detailed description of genotyping and quality control).

### Polygenic scores

DNA was used to calculate polygenic scores as an index for genetic vulnerabilities for psychiatric diseases.

We constructed GPS (genome-wide polygenic scores) using LDpred^36^, which corrects GWAS summary statistics for local linkage disequilibrium (LD). See Allegrini et al. 2019^37^ for a detailed description of LDpred analytic strategies used in calculating GPS in the TEDS sample. Here we used fraction 1.0 for all GPS, which assumes that all markers have non-zero effects. This faction of causal markers has shown to be the most predictive for psychiatric traits.

The following GWAS summary statistics were used to create GPS: Schizophrenia^38^, Bipolar disorder^39^, Depression^40^, Anxiety^41^, ADHD (attention deficit hyperactivity disorder)^42^, ASD (autism spectrum disorder)^43^, EA3 (educational attainment)^44^, Risk taking Sensitivity^45^, Cross-disorder (combining eight psychiatric disorders)^46^, OCD (obsessive compulsive disorder)^47^, PTSD (post-traumatic stress disorder)^48^, Neuroticism^49^ and Insomnia^50^. These summary statistics were selected because they represent the most powerful GWAS for behavioural traits at the time of the analyses. Note that some summary statistics contained 23andMe data, the summary statistics employed here do not include 23andMe data due to their data availability policy.

In addition to examining the genetic vulnerabilities of each GPS, we also used the first principal component of the 11 psychiatric GPS (excluding EA3 to limit the final score to psychiatric traits; and cross-disorder GWAS summary statistics as this already includes psychiatric disorders included in this composite measure) to index general genetic vulnerability to behavioural problems. This approach comes with limitations, mainly the sample overlap between some of the GWAS (mostly from shared controls), which may inflate the correlation between the GWAS results. It has been shown, however, that different methods used to derive a genomic p factor yield similar results^51^.

### Statistical analyses

Our statistical analysis plan was registered with the Open Science Framework (https://osf.io/gzrk3) prior to accessing the data. Scripts have been made available on the OSF site. All analyses were completed using SPSS Statistics software^52^ and R version 4.0^53^.

#### Phenotypic analyses

We assessed mental health traits in a sample of up to 4,773 unrelated individuals in their mid-twenties during four waves of data collection (T2-T5) during the COVID-19 lockdown and compared their responses to the same traits assessed in 2018 (T1). We compared the average differences in these traits from T1 to T2-T5 by comparison of means and SD at T1-T5, and we did this also separately for males and females. MANOVA was used to test for the significance of mean differences between time points, differences between males and females at all time points, also testing for interactions between time and sex.

Because significant, though small, sex differences emerged, we corrected all scores for mean sex differences using the regression method. Correcting for sex and age is important in the analysis of twin data because members of a twin pair are identical in age and identical twins are identical for sex, which, if uncorrected, would inflate twin estimates of shared environment^54^. These age- and-sex adjusted standardised residuals were used in all subsequent analyses.

Several variables were skewed. In our previous study^55^, sensitivity analyses were performed to examine the influence of the van der Waerden transformation on results. The results remained highly similar to untransformed data analyses; for this reason, we decided to use untransformed values in all our analyses.

We used latent growth curve (LGC) models to extract stable individual differences from before the COVID-19 pandemic started (T1), evaluate individual differences at the starting point (2018 data collection) and extract the rate of change over the pandemic (T2-T5) for the same traits. We fitted LGC models for all mental health measures separately, testing for linear, quadratic and piecewise trends in the data. We used the R package lavaan for LGC. FIML (full information maximum likelihood) was used to account for missing data^56^.

In addition, we conducted latent profile analyses (LPA) to identify any subpopulations in our sample that responded differentially to the COVID-19 pandemic. LPA was fitted to each of the mental health measures separately. LPA is a statistical method for identifying homogenous subgroups of individuals based on a set of continuous measured variables called indicators. Classification of individuals into latent classes is probabilistic and LPA allows for selection of the optimum numbers of classes (the optimum model), through comparison of model fit indices. Longitudinal LPA is a special case of LPA where the indicators consist of repeated measures of the same variable across time (T1-T5). For each outcome (symptoms of mental health, see Table 1), models with stepwise increases in numbers of classes, from 2 to 7 classes, were fitted. In all models, variances were equated and covariances fixed to 0 and missing values across timepoints were imputed (single imputation using mix package alongside tidyLPA package). We used the tidyLPA package ^57^, which draws on functionality of mclust^58^ in R 3.6.2 for all analyses.

#### Genetic analyses

The classical twin design was used to assess genetic and environmental contributions to individuals’ traits at each wave of assessment. The twin method capitalises on the genetic differences between twins – that is, MZ (monozygotic, identical) twins, who are 100% similar genetically, while DZ (dizygotic, non-identical) twins share on average 50% of their segregating genes. Environmental factors that make members of twin pairs similar to each other are defined as shared environmental factors, and environmental factors that do not contribute to similarities between twin pairs are defined as non-shared environmental factors. Using these family relatedness coefficients, it is possible to estimate the relative influence of additive genetic (A), shared environmental (C), and non-shared environmental (E) effects on the variance and covariance of phenotypes, by comparing intraclass correlations for MZ and DZ twins^24^. In the model, non-shared environmental variance also includes any measurement error. These parameters can be estimated more accurately using structural equation modelling (SEM), which also provides 95% confidence intervals and estimates of model fit. The SEM program OpenMx was used for all twin model-fitting analyses^59^.

The univariate model can be extended to a multivariate model to investigate the aetiology of the covariance between two traits. This method also enables the estimation of the genetic correlation (rG), indicating the extent to which the same genetic variants influence two phenotypes. The shared environmental correlation (rC) and non-shared environmental correlation (rE) can also be estimated ^24,60^. A correlated factor solution was used to calculate genetic correlations between mental health measures from T1-T5. Cholesky Decomposition analysis, which is conceptually similar to hierarchical regression, was used to estimate the extent to which genetic and environmental effects at T2, T3, T 4 and T5 are independent of T1, indicating the aetiology of changes in mental health symptoms during COVID-19. We conducted these analyses for each mental health measure.

Genome-wide polygenic scores (GPS) were used to estimate the variance explained in mental health measures from T1 to T5 using linear regression adjusted for sex, age, the first ten PCs, genotyping-batch and genotyping-chip effects. In our preregistered plan we had specified that we would use GPS to predict both intercept and slope in mental health traits (LGC model), but since we did not find significant slope in the models, we were not able to complete this step in our analyses.

#### Extremes analyses

To assess whether extreme groups are differentially affected by the COVID-19 pandemic and associated lockdown, we compared extreme groups defined as <1SD and >1SD of the standardised scores for the following variables: SES (socioeconomic status assessed when twins joined TEDS in infancy; SES is stable across development, the correlation between SES from first contact and SES collected when twins were 16 is 0.71); phenotypic psychopathology factor (1^st^ principal component of mental health for data collected (see Table 1) in 2018 using PCA approach; see Cheesman, Allegrini et al. (2020)^61^ for details on PCA approach on mental health measures using the same data); and genomic psychopathology factors (1^st^ principal component of all psychiatric polygenic scores), and also separately for Depression^40^, Anxiety^41^, Risk taking Sensitivity^45^, Cross-disorder^46^ and educational attainment (EA)^62^, as these GPS were closely related to the phenotypes studied here.

#### Environmental correlates of COVID-19 lockdown

We also studied the association between mental health traits from T2-T5 and environmental factors (e.g., lockdown conditions), physical health (e.g., COVID-19 symptoms, short and long-term), and life changes (e.g., mental health or financial worries) using CRISIS measures (see Figure 1). In our preregistered analyses plan we decided to test if CRISIS measures predict intercept and slope in mental health, however, we did not detect significant linear slope in mental health measures. Therefore, we examined the effect of environmental extremes on mental health outcomes during the COVID-19 pandemic and lockdown. For example, we compared low versus high (+/-one SD) family socioeconomic status (SES) obtained when the twins were infants. For quantitative measures (e.g., worries about mental health or financial wellbeing) we report the phenotypic correlations.

## Results

### Descriptive statistics for mental health from T1 to T5

Figure 2 illustrates the means and standard errors for the mental health measures from T1 to T5. The average changes in mental health symptoms were small to medium (average partial eta squared from T1 to T5 =0.026) and mainly occurred over the 2-year period from 2018 to March 2020 (average Cohen’s d from T1 to T2= 0.14), with no evidence of average worsening mental health as the pandemic persisted. The largest negative effects on mental health were increased (worsening) general anxiety (Cohen’s d = 0.17 from T1 to T2), increased hyperactivity (Cohen’s d=0.37) and decreased prosocial behaviour (Cohen’s d=0.36). These worsening mental health effects are considered to be small to medium effect sizes (a Cohen’s d of 0.35 accounts for 3% of the variance)^63^. For anxiety and prosocial behaviour, the negative effect (worse symptoms on average) persisted from T2 to T5; for hyperactivity, the effect decreased with time. Moreover, some changes were positive for mental health: conduct problems, emotional problems and self-harm decreased. However, these small to medium effect sizes based on Cohen’s criteria are still meaningful^64^. Based on the current evidence an effect size of 0.3 for psychiatric traits is considered to be medium to large, and similar that would be expected after natural disasters (e.g., earthquake)^65^.

**Figure 2.**
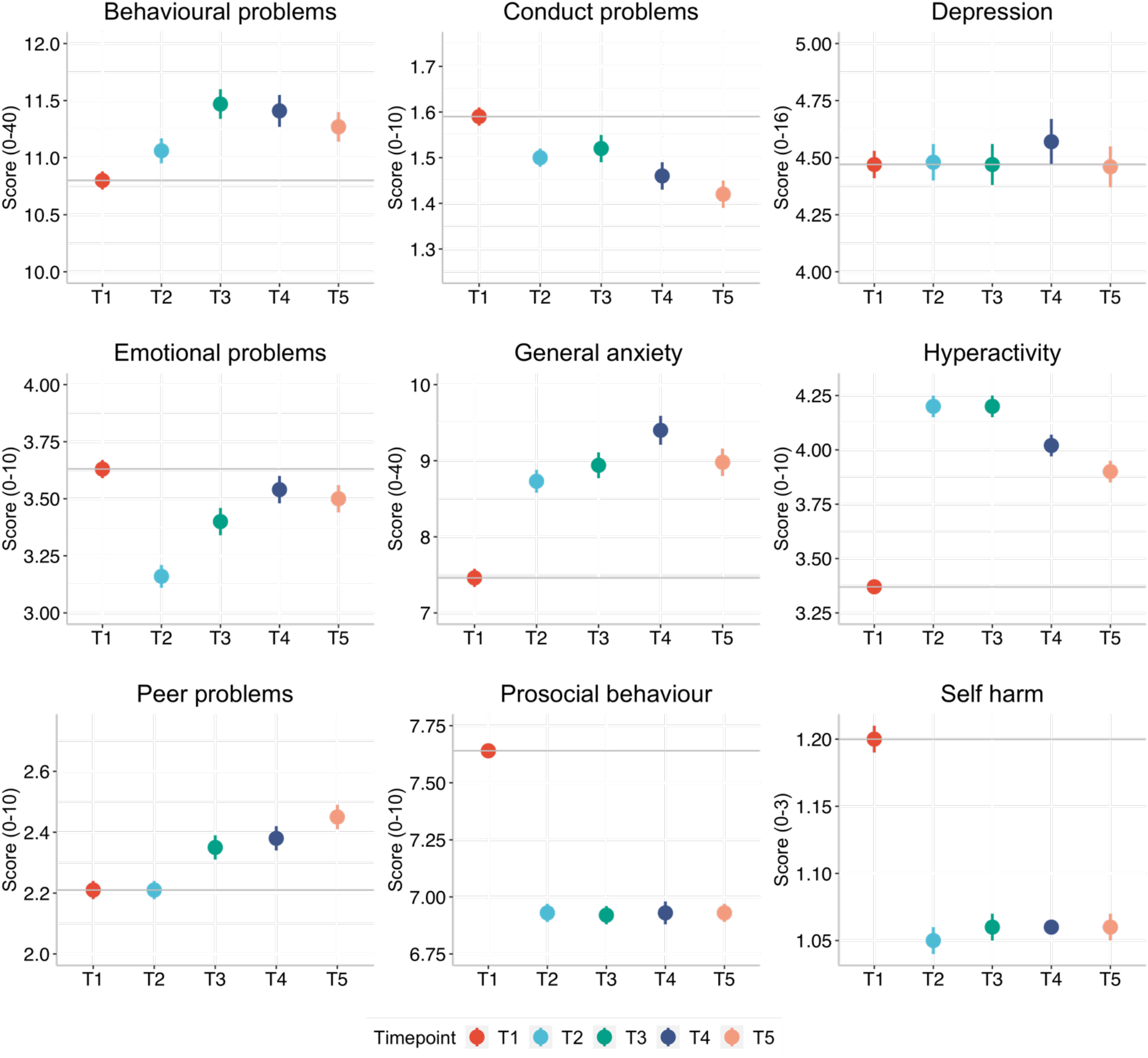
Descriptive statistics (means and standard errors) for all mental health measures from T1 to T5.

These descriptive statistics are based on one randomly selected twin per pair, but the results were virtually identical when the analysis was repeated with their co-twins. Details of descriptive statistics results of MANOVA are included in Supplementary Table S1. On average, females reported more mental health problems than males, but these mean sex differences only explained on average 3% of the variance in these measures (average partial eta squared 0.035). There were no significant time*sex interactions. Descriptive statistics for males and females separately, and results for MANOVA, are presented in Supplementary Table S1.

Supplementary Table S2 presents descriptive statistics for CRISIS measures from T2 to T5. On average the possible COVID-19 symptoms were slightly lower at the start of the pandemic compared to later data collection points; however, the worries (both worries about COVID-19, mental health, and finances) were slight elevated at the start of the pandemic and declined slightly as the pandemic continued.

In our preregistered analyses plan (https://osf.io/gzrk3) we specified LGC models to estimate individual differences at the starting point (2018 data collection, T1) and individual differences in the rate of change that occurs over time for the same traits (slope T1 to T5). We predicted that there would be a significant linear slope from T1-T5 for all mental health measures. We had plans to test whether GPS and CRISIS measures (Covid worries, mood states, life changes during the pandemic) predict intercept and slope in mental health symptoms. However, while the LGC identified a significant change (slope) from T1-T5 (see Supplementary Table S3 for results), variance in these slopes was very small. In addition, the linear trend across T1-T5 is not really fitting data well given the rupture happening at T2 for many phenotypes. It was therefore not reasonable to predict the change from T1-T5. The quadratic trends in the data fitted better than the model with just one slope. We could not test piecewise trends in the data as there was only one datapoint before change happened in T2, and a minimum of three data points is required to appropriately estimate a linear slope. We decided not to predict the change from T1 to T2 for several reasons: 1) it is uncertain if changes over a 2-year period can be due to pandemic as the maturation hypothesis could fit equally well; 2) the increase only happened between T1 and T2, we did not observe worsening of mental health symptoms over the pandemic; 3) the predictor variables (COVID-19 worries, mood states, life changes during the pandemic) were collected from T2, therefore not applicable for prediction of change from T1 to T2. Here, we opted to use latent profile analyses instead to check for the presence of subgroups in each mental health outcome.

### Latent profile analyses

Latent profile analyses, using standardized measures of mental health symptoms at each time point (mean of 0 and SD1), are in agreement with the descriptive statistics showing little change in mental health symptoms during the COVID-19 pandemic., even though subgroups were identified. For most measures, the optimal model identified 7 profiles. Most differences across profiles were due to changes from T1 to T2, and the profiles did not change much from T2-T5. Most of the sample were in the middle profiles, as illustrated in Figure 3, which showed no changes in mental health symptoms. The sample sizes for the profiles showing an increase or decrease in mental health were small as was the change in mental health symptoms (less than half a SD on average). For example, for hyperactivity, individuals in latent profile 2 (5.5% of the sample) showed an increase in symptoms from T1 to T2, and this remained elevated through T3, T4 and T5. Similarly, profile 3 (21.6% of the sample) showed an increase from T1 to T2, but the symptoms returned to pre-pandemic level. Model fit statistics for all LPA are presented in Supplementary Table S4.

**Figure 3.**
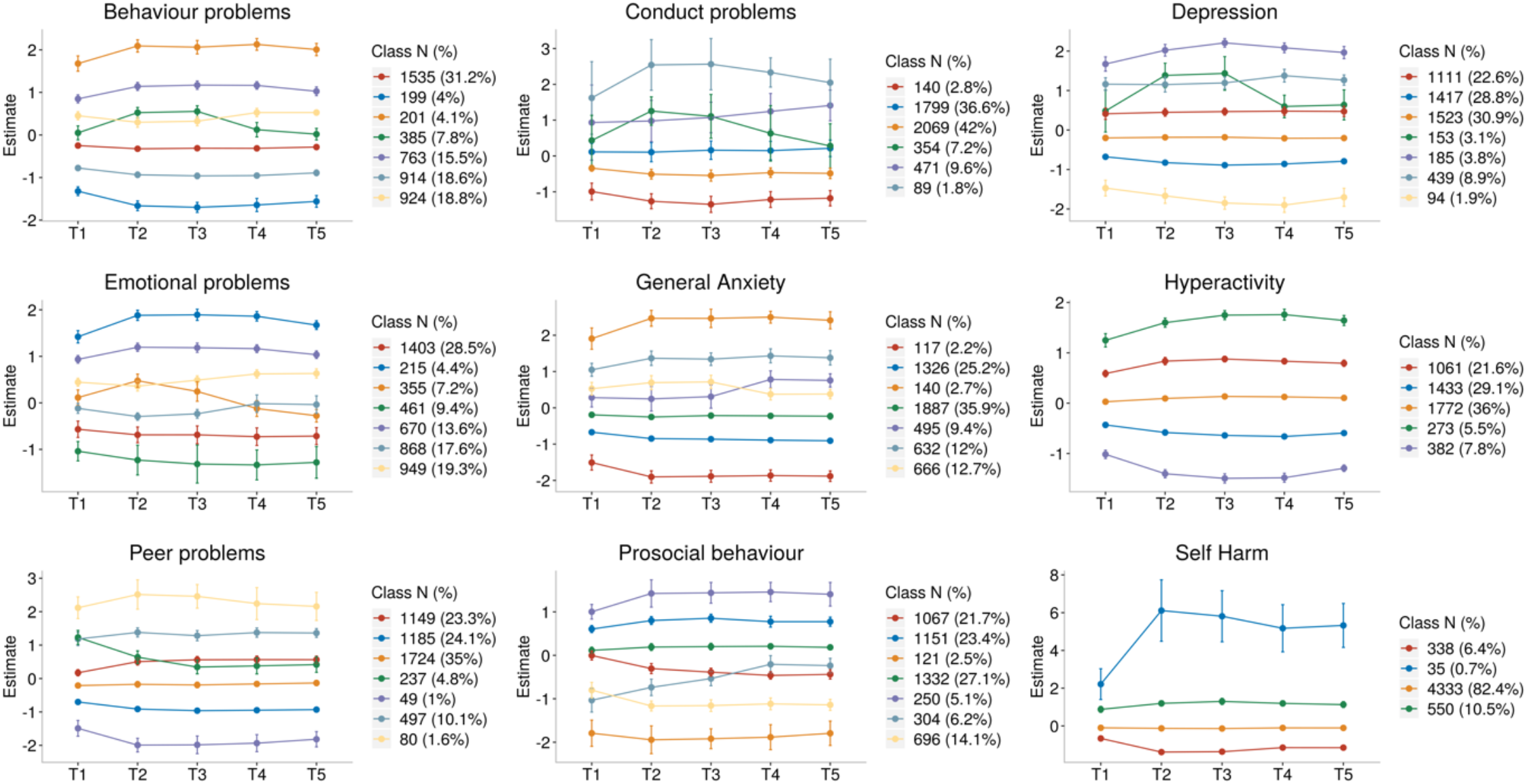
Latent profile analyses presenting the optimum model for each mental health outcomes (95% CI as error bars).

Although self-harm dramatically increased for one latent class, only 35 individuals (fewer than 1% of the sample) were in this profile. In addition, this measure was highly skewed. For these reasons we did not perform additional analyses with the self-harm measure, although we plan to study the predictors of self-harm and its changes in our future research.

### Individual differences

These small unsystematic mean differences, as well as latent profiles, could mask large individual differences. Figure S1 presents the pattern of individual variability in mental health measures from T1 to T5, which shows large individual differences at every data collection as well as individual differences in changes in mental health measures during the COVID-19 pandemic. The overall (mean) trajectories are presented in black (similar to means in Figure 2) and show no change from T1-T5, except for hyperactivity and prosocial behaviour which change for the worse from T1 to T2.

If there are large differences in how individuals respond to the COVID-19 pandemic and associated lockdown then we might predict increased variance. We have already shown in our previous report that we did not observe variance differences from T1 to T2 ^27^. Here we found that the variance remains similar from T1 to T5 (see Supplementary Table S1 for standard deviations across measures from T1-T5). We did not observe an increase in variance following the COVID-19 crisis.

In addition, if COVID-19 re-shuffled individual differences in mental health symptoms then we would expect to see little correlation between mental health symptoms from T1 to T5. However, in our previous report, we noted substantial correlations between measures across the three years from T1 to T2 (average correlation = 0.59). Here we found even larger correlations from T2-T5 (see Figure 4), which had an average interval of three months (average correlation = 0.65). The one-month interval between T2 and T3 is larger still (average correlation of 0.69) and comparable to the average two-week test-retest reliability from TEDS’ preparatory work for our 2018 (T1) assessment of the twins (average test-retest reliability = 0.75).

**Figure 4.**
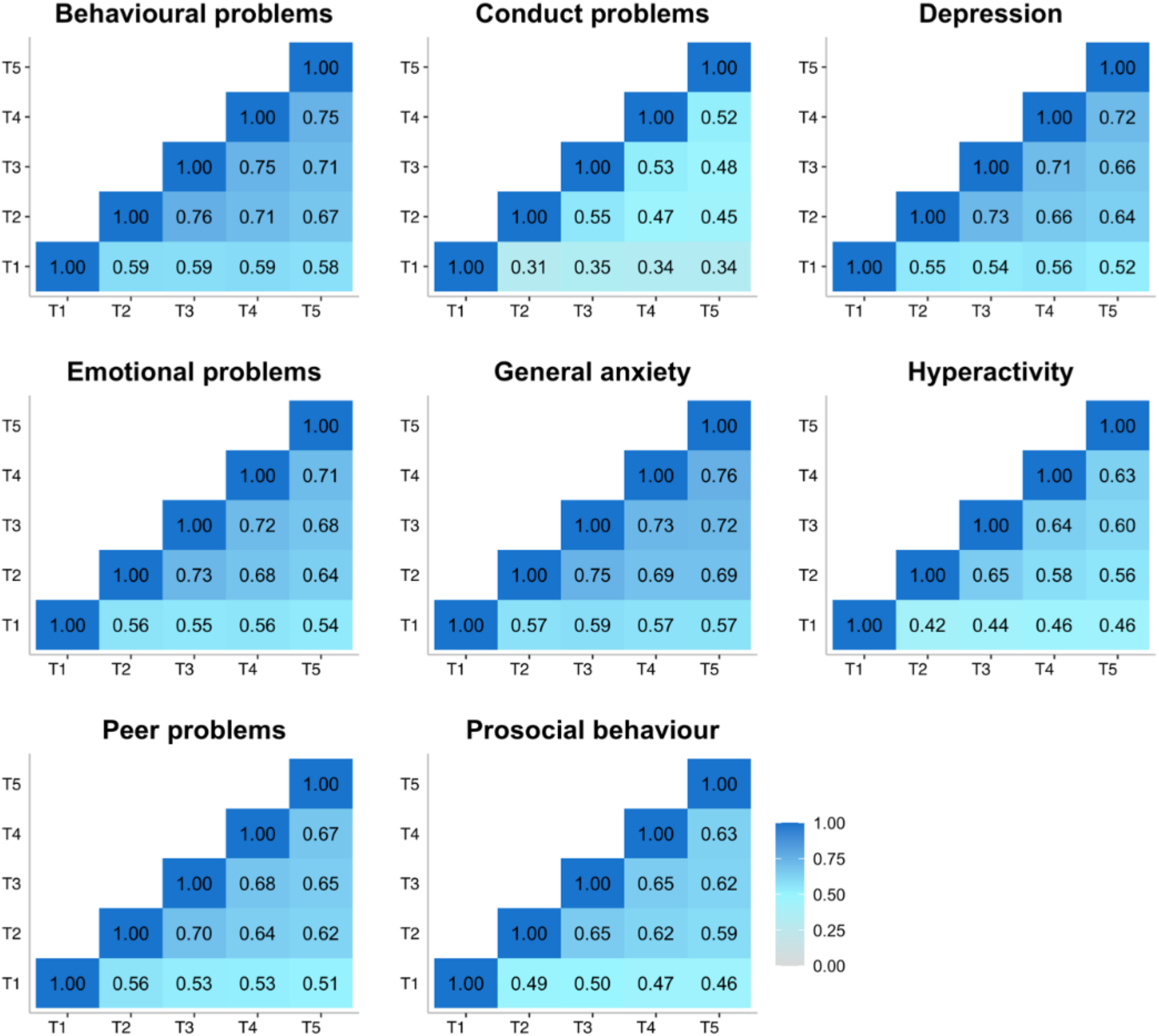
Phenotypic correlations from across time points.

See Supplementary Table S5 for correlation coefficients with 95% confidence intervals.

### Genetic analyses

As illustrated in Figure 5a, twin estimates of heritability for mental health measures were on average 33%, with the majority of the variance explained by non-shared environmental components. Importantly, the heritability of mental health symptoms studied here did not change systematically despite the COVID-19 pandemic and lockdown. One exception is conduct problems that only showed significant heritability at T1 and T4, although the heritability estimates from T1 to T5 have overlapping confidence intervals. Full univariate model-fitting results with 95% confidence intervals are presented in Supplementary Table S6. Twin model-fitting also showed that variance did not increase from T1 to T5, as indicated by the unstandardised variance components (See Supplementary Figure S2).

The most striking genetic result was that the average genetic correlation was .95 between T1 and T2-T5 (Figure 5b). Non-shared environmental correlations were moderate with the average correlation of .41, indicating that the non-shared environmental factors that explain individual differences in mental health symptoms at one time, were correlated with environmental factors at another time during the COVID-19 pandemic. (See Supplementary Table S7 for genetic, shared and nonshared environmental correlations with 95% confidence intervals.) Supplementary Table S8 presents the results for multivariate Cholesky analyses, which show that there is negligible genetic variance at T2-T5 independent of genetic variance at T1.

**Figure 5.**
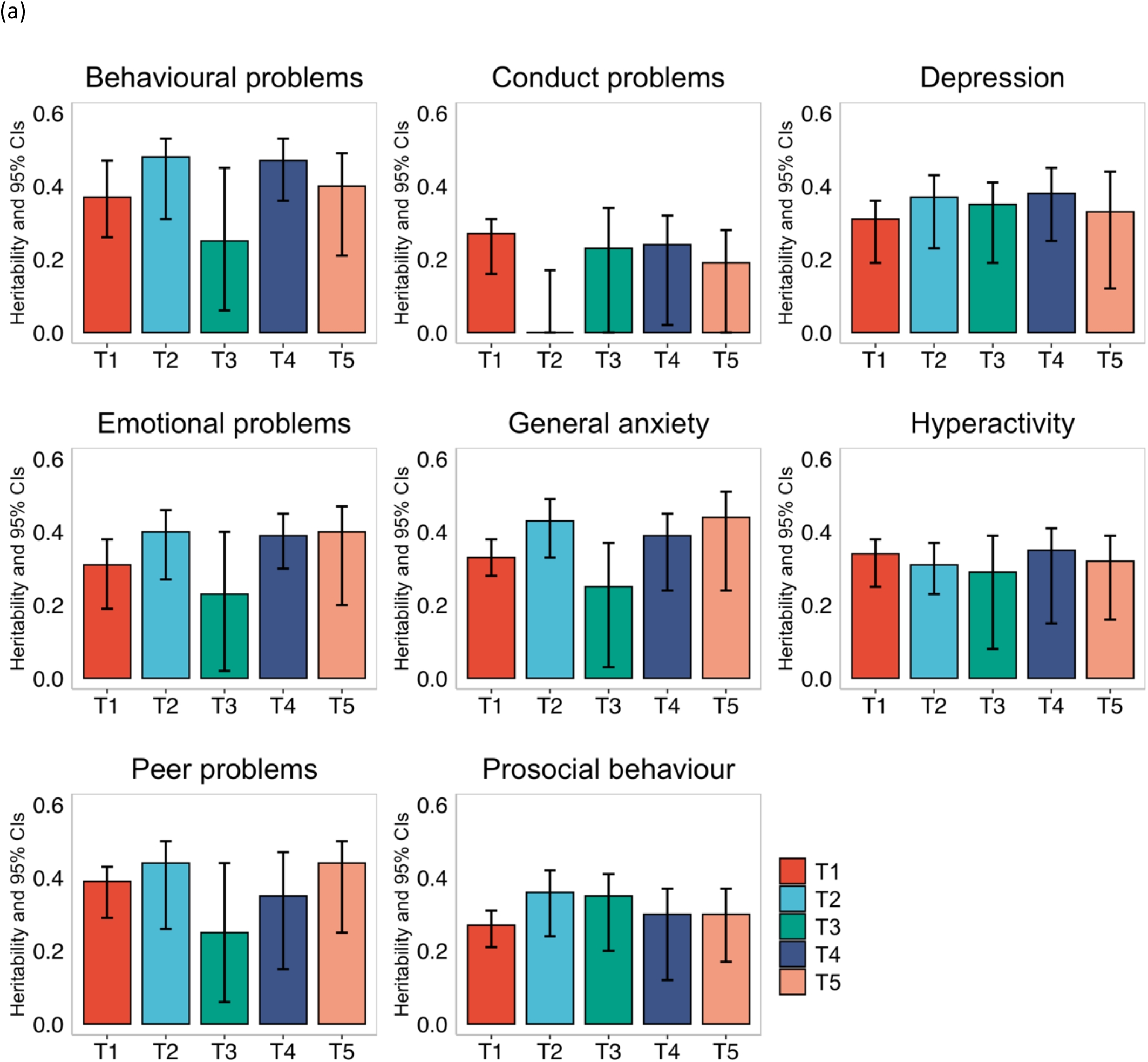
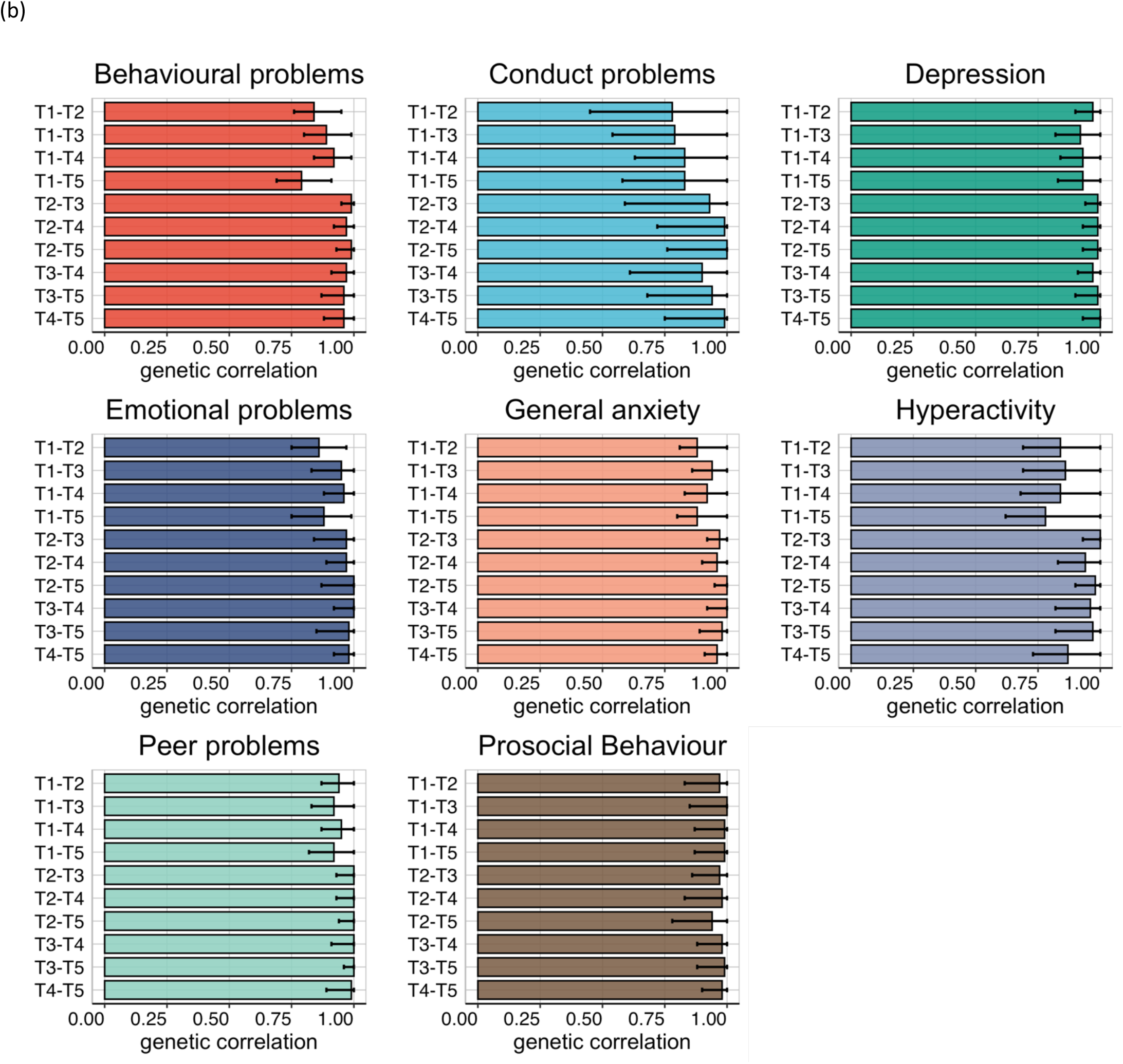
a) Univariate twin model-fitting results; b) Genetic correlations with 95% confidence intervals.

### Polygenic score analysis

We also calculated the variance explained by all polygenic scores (see Methods). We hypothesised that GPS would significantly predict variance in mental health measures, but while some of the models were significant after multiple testing correction, many were not, and the average prediction was low. On average, the polygenic scores (both genomic p factor and individual GPS) explained less than 1% of variance in mental health measures; importantly, the variance explained did not change from T1 to T5. See Supplementary Tables S9-S14 for the results of GPS prediction longitudinally from T1 to T5.

### Extremes analyses of individuals with pre-existing mental health problems

We compared two groups of individuals: those with pre-existing mental health problems (+1SD on the 1^st^ PC of mental health problems in 2018; N=265-429; see Supplementary Table S15 for details) and those who had lower than average mental health problems before the pandemic started (−1SD on the 1^st^ PC of mental health problems in 2018; N=227-329; see Supplementary Table S15 for details). As illustrated in Figure 6, individuals with pre-existing mental health problems reported increased problems at T2 for most measures (average d = .47 across all measures). However, these increases in mental health problems returned to pre-pandemic levels at T3, T4 and T5 and only remained slightly elevated for general anxiety, hyperactivity and prosocial behaviour (See Supplementary Table S15-18 for descriptive statistics, including the sample size in extreme groups, MANOVA results and pairwise comparisons across timepoints).

**Figure 6.**
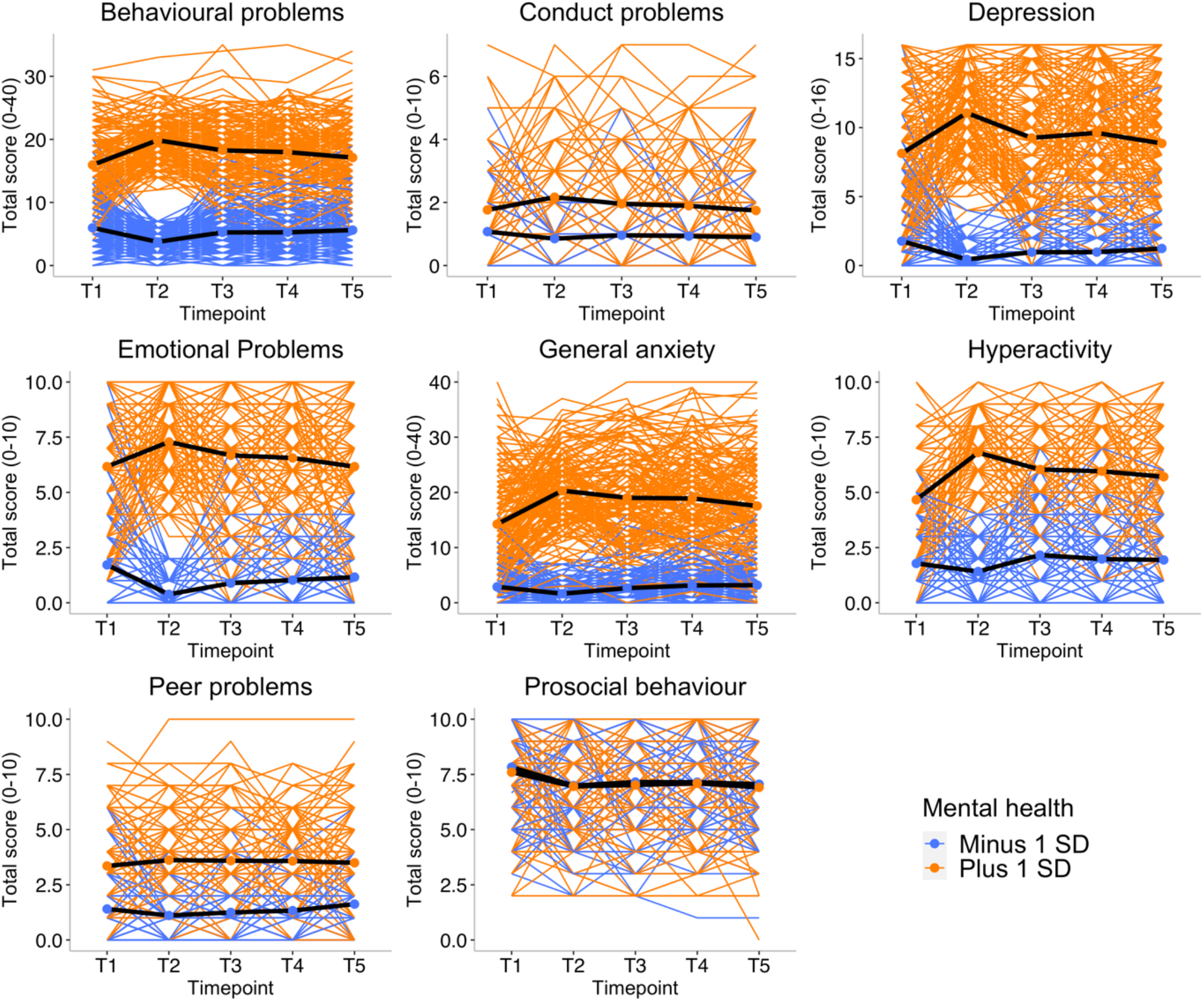
Patterns of individual variability across T1-T5 for all mental health measures separated by -/+ 1 SD on p-factor at T1 prior to the start of the pandemic. Individual trajectories are presented as coloured lines and average mean trajectory as a black line.

Polygenic scores at extremes did not show different trajectories for the low and high extremes groups (+/-1SD); these results are presented in Supplementary Figures S3-S8.

### Environmental extremes

We also examined the effect of environmental extremes on mental health outcomes during the COVID-19 pandemic and lockdown. For example, we compared low versus high (+/-one SD) family socioeconomic status (SES) obtained when the twins were infants. As expected, we found that on average individuals with lower SES reported worse mental health than individuals with higher SES, although the differences were small (partial eta squared=0.012; see MANOVA results in Supplementary Table S19). However, we did not see differential trajectories for these groups across the COVID-19 pandemic – that is, there was no moderation between time and SES in explaining mental health outcomes (See Supplementary Figure S9 and for MANOVA results Table S19).

We also investigated other possible environmental moderators, including parenthood, having access to green or garden space, having a family member experience job loss, and having extreme financial struggles such as worrying about paying for food. We found little evidence that these environmental factors had an effect on the response of these young adults to the COVID-19 pandemic (Supplementary Figures S10-S13).

In addition, we did not observe differences in mental health symptoms between individuals who had COVID-19 diagnoses or COVID-like symptoms at any point during the pandemic (26%) compared to individuals with no symptoms (see Supplementary Figure S14). Nor did differences emerge between individuals with versus without possible “long-COVID” symptoms, although the possible long-COVID sample was small (N=47; See Supplementary Figure S15).

Finally, we show that the sum score of major family events (infected by COVID-19, hospitalised, losing job or loss of family member) was only correlated weakly (r = 0.07 on average) with mental health symptoms. A composite score of worries correlated modestly with mental health symptoms (r=0.21 on average). See Supplementary Figure S16.

## Discussion

Despite the expectation of catastrophic effects on the COVID-19 pandemic on mental health, especially for young people, we find few longer-term consequences, phenotypically or aetiologically. The largest average negative effects – increased hyperactivity and decreased prosocial behaviour – were modest and emerged temporarily during the first month after lockdown (Figure 2). For prosocial behaviour, the change disappeared entirely four months after lockdown; for hyperactivity, the change diminished 7 and 11 months after lockdown. In addition to these modest negative effects, several average changes were in the positive direction, such as decreased conduct problems and decreased emotional problems.

Our latent profile analyses (Figure 3) supported these phenotypic conclusions at the level of individual differences, we did not identify any meaningful subgroups, with one possible exception: One small latent class of 35 individuals showed a sharp rise in self-harm which continued throughout the pandemic, despite the overall decline in self-harm before and after lockdown (Figure 2). These findings warrant caution, this self-reported measure of self-harm was reported on four-point scale and produced a highly skewed distribution. It is also possible that this set of 35 individuals was self-selecting to the study specifically because of their unusual self-harm results. These results warrant further investigation.

Part of the reason for the perception of the negative effect of COVID-19 on mental health is that reports often focus on statistical significance of mean differences rather than effect size. With the large sample size of the current study, almost any mean difference is statistically significant, despite their small effect sizes. These findings add to the growing evidence or smaller negative effects on mental health than previously expected^15–18^. Our results are also in line with reports that suggest that population levels symptoms of mental health increase immediately after lockdown, but then decreased again as the pandemic continues^14,16,66^.

An exception is that young adults in England and Wales who reported more mental health problems before the pandemic showed even more problems during the pandemic (Figure 6). This finding is well aligned with reports that pandemic is especially detrimental for individuals with existing psychological and psychiatric vulnerabilities^6,8,22,32^. However, we found that most of these effects were confined to one month after lockdown, and mental health generally returned to pre-pandemic levels as the COVID-19 crisis wore on.

Focusing on individual differences rather than average differences enabled several analyses. First, a simple comparison of phenotypic variance across time was illuminating. If individuals responded differently to the COVID-19 crisis, we would expect to see increased variance during the pandemic, but no increase was observed. Second, even in the absence of phenotypic changes in variance, it is possible that the underlying genetic and environmental causes of variance could shift because of the COVID-19 crisis, that is genetic effects explaining individual differences would be amplified or reduced, it is also possible that new (innovative) genetic effects could explain some variations in mental health symptoms. However, no systematic changes in heritability were observed (Figure 5a). The most striking genetic result was the average genetic correlation of 0.95 for mental health measures before and during the pandemic (Figure 5b), indicating that on average the genetic factors explaining individual differences in mental health did not change during the pandemic.

Similarly, environmental factors did not moderate the effect of the pandemic. The trajectories of mental health across the year of the COVID-19 crisis did not differ as a function of SES, financial struggles, or negative family events. Nor did we find any effect of reported COVID-19 symptoms on mental health.

These findings did not confirm our pre-registered hypotheses (https://osf.io/gzrk3) for changes in mental health symptoms; we did not find evidence for this in our sample on average. However, the group of young adults with pre-existing mental health vulnerabilities experienced increased mental health symptoms at the start of the pandemic, with their increased in their mental health problems subsiding as the pandemic persisted. We conclude that our results speak to the resilience of young people, whose lives were highly disrupted socially and economically by the crisis.

It should be noted that our results are limited to young adults in their twenties and will not necessarily generalise to other ages. Because this age group will be vital to the economy in the coming decades, it is good news for the economy and society if young people come through the pandemic relatively unscathed.

Although the sample is reasonably representative of its cohort, it consists of twins, and is more white and more educated than the rest of the UK general population. Attrition and selection bias might have influenced the results. It is well documented that participants in most studies tend to be slightly healthier and more educated than the general population, and therefore, the associations observed between study variables might be biased by collider effects^67^. This bias may be amplified with data collections happening during the pandemic^68^. Our sample also experienced attrition, and healthier, more educated participants took part in all data collection waves (see Table S20). However, the results here remained highly similar whether we used the full data (descriptive statistics), used imputation (LPA), or full-information-maximum-likelihood (FIML, LGC) statistical approaches, and our sample at T1, before the pandemic, is reasonably representative of the 1994-1996 birth cohort. More research is needed to assess the effects of the COVID-19 crisis on mental health in other populations and age groups.

We conclude that the mental health of young adults in England and Wales has been remarkably resilient to the effect of global pandemic and associated lockdown. Although mental health symptoms worsened immediately after the first lockdown, these effects subsided with time. The effects were stronger for young adults with pre-existing psychological and behavioural problems, however, their worsening symptoms also subsided as the pandemic evolved. These conclusions should not mean that the distress and worry many of our participants felt during this crisis should be dismissed nor that there could not be lasting effects of the crises for the most vulnerable ones. These results inform preventative interventions, indicating that the largest vulnerabilities of crises were felt at the start of the pandemic. The COVID-19 pandemic is ongoing, and the long-term impacts of the pandemic on the mental health of young adults are not known. Studying the psychological vulnerabilities of young adults should remain a research priority as young adulthood is a tipping point for life-long psychological problems.

## Data Availability

Data for this study came from the Twins Early Development Study (TEDS). Researchers can apply for access to the data: https://www.teds.ac.uk/researchers/teds-data-access-policy.

## Acknowledgements

We gratefully acknowledge the ongoing contribution of the participants in the Twins Early Development Study (TEDS) and their families. TEDS was supported by a program grant to RP from the UK Medical Research Council (MR/M021475/1 and previously G0901245), with additional support from the US National Institutes of Health (AG046938). RP is supported by a Medical Research Council Professorship award (G19/2). KR is supported by a Sir Henry Wellcome Postdoctoral Fellowship. SvS is supported by a Jacobs Foundation fellowship. The authors acknowledge use of the research computing facility at Kings College London, Rosalind (https://rosalind.kcl.ac.uk), which is delivered in partnership with the NIHR Biomedical Research Centre at South London & Maudsley and Guys & St. Thomas NHS Foundation Trusts, and part-funded by capital equipment grants from the Maudsley Charity (Grant Ref. 980) and Guys & St. Thomas Charity (TR130505).

This paper represents independent research part-funded by the National Institute for Health Research (NIHR) Maudsley Biomedical Research Centre at South London and Maudsley NHS Foundation Trust and King’s College London. The views expressed are those of the author(s) and not necessarily those of the NHS, the NIHR or the Department of Health and Social Care.

