## Supplementary Material for "The consequences of a year of the COVID-19 pandemic for the mental health of young adult twins in England and Wales"

Table S1. Descriptive statistics for one randomly selected twin per pair (a); for females only (b), and for males only (c); MANOVA results (d)

(a)

| | T1 | | | T2 | | | T3 | | | T4 | | | T5 | | |
|---|---|---|---|---|---|---|---|---|---|---|---|---|---|---|---|
| | **N** | **Mean (SD)** | **SE** | **N** | **Mean (SD)** | **SE** | **N** | **Mean (SD)** | **SE** | **N** | **Mean (SD)** | **SE** | **N** | **Mean (SD)** | **SE** |
| Emotional problems | 4771 | 3.63 (2.71) | 0.04 | 2437 | 3.16 (2.68) | 0.05 | 2006 | 3.40 (2.72) | 0.06 | 1830 | 3.54 (2.77) | 0.06 | 1940 | 3.5 (2.73) | 0.06 |
| Conduct problems | 4772 | 1.59 (1.31) | 0.02 | 2437 | 1.50 (1.20) | 0.02 | 2006 | 1.52 (1.20) | 0.03 | 1830 | 1.46 (1.14) | 0.03 | 1940 | 1.42 (1.13) | 0.03 |
| Hyperactivity | 4771 | 3.37 (2.21) | 0.03 | 2437 | 4.20 (2.26) | 0.05 | 2006 | 4.20 (2.28) | 0.05 | 1830 | 4.02 (2.24) | 0.05 | 1940 | 3.9 (2.3) | 0.05 |
| Peer problems | 4771 | 2.21 (1.82) | 0.03 | 2437 | 2.21 (1.65) | 0.03 | 2006 | 2.35 (1.70) | 0.04 | 1830 | 2.38 (1.68) | 0.04 | 1940 | 2.45 (1.73) | 0.04 |
| Prosocial behaviour | 4773 | 7.64 (1.89) | 0.03 | 2437 | 6.93 (1.95) | 0.04 | 2006 | 6.92 (1.99) | 0.04 | 1830 | 6.93 (1.96) | 0.05 | 1940 | 6.93 (1.96) | 0.04 |
| Behaviour problems | 4771 | 10.80 (5.87) | 0.08 | 2437 | 11.06 (5.62) | 0.11 | 2006 | 11.47 (5.74) | 0.13 | 1830 | 11.41 (5.79) | 0.14 | 1940 | 11.27 (5.76) | 0.13 |
| Depression | 4769 | 4.47 (4.13) | 0.06 | 2437 | 4.48 (3.97) | 0.08 | 2006 | 4.47 (4.00) | 0.09 | 1830 | 4.57 (4.22) | 0.1 | 1940 | 4.46 (4.09) | 0.09 |
| General anxiety | 4250 | 7.46 (7.50) | 0.12 | 2437 | 8.73 (7.63) | 0.15 | 2006 | 8.94 (7.77) | 0.17 | 1830 | 9.40 (8.02) | 0.19 | 1940 | 8.98 (7.76) | 0.18 |
| Self Harm | 4229 | 1.20 (0.67) | 0.01 | 2410 | 1.05 (0.28) | 0.01 | 1981 | 1.06 (0.31) | 0.01 | 1809 | 1.06 (0.33) | 0 | 1921 | 1.06 (0.34) | 0.01 |

(b) Females

| | **N** | **Mean (SD)** | **SE** | **N** | **Mean (SD)** | **SE** | **N** | **Mean (SD)** | **SE** | **N** | **Mean (SD)** | **SE** | **N** | **Mean (SD)** | **SE** |
|---|---|---|---|---|---|---|---|---|---|---|---|---|---|---|---|
| Emotional problems | 2970 | 4.14 (2.72) | 0.05 | 1665 | 3.68 (2.70) | 0.07 | 1395 | 3.89 (2.74) | 0.07 | 1279 | 4.01 (2.76) | 0.08 | 1374 | 3.93 (2.74) | 0.07 |
| Conduct problems | 2970 | 1.56 (1.31) | 0.02 | 1665 | 1.51 (1.19) | 0.03 | 1395 | 1.57 (1.25) | 0.03 | 1279 | 1.49 (1.19) | 0.03 | 1374 | 1.46 (1.18) | 0.03 |
| Hyperactivity | 2970 | 3.32 (2.21) | 0.04 | 1665 | 4.26 (2.27) | 0.06 | 1395 | 4.27 (2.29) | 0.06 | 1279 | 4.10 (2.27) | 0.06 | 1374 | 3.95 (2.33) | 0.06 |
| Peer problems | 2970 | 2.22 (1.87) | 0.03 | 1665 | 2.14 (1.67) | 0.04 | 1395 | 2.31 (1.72) | 0.05 | 1279 | 2.40 (1.72) | 0.05 | 1374 | 2.41 (1.75) | 0.05 |
| Prosocial behaviour | 2971 | 8.01 (1.71) | 0.03 | 1665 | 7.28 (1.82) | 0.04 | 1395 | 7.26 (1.88) | 0.05 | 1279 | 7.26 (1.85) | 0.05 | 1374 | 7.25 (1.83) | 0.05 |

| | | | | | | | | | | | | | | | |
|---|---|---|---|---|---|---|---|---|---|---|---|---|---|---|---|
| Behaviour problems | 2970 | 11.24 (6.02) | 0.11 | 1665 | 11.59 (5.70) | 0.14 | 1395 | 12.04 (5.88) | 0.16 | 1279 | 12.00 (5.90) | 0.16 | 1374 | 11.75 (5.92) | 0.16 |
| Depression | 2969 | 4.91 (4.33) | 0.08 | 1665 | 4.99 (4.11) | 0.1 | 1395 | 4.94 (4.14) | 0.11 | 1279 | 5.04 (4.37) | 0.12 | 1374 | 4.86 (4.23) | 0.11 |
| General anxiety | 2708 | 8.27 (7.72) | 0.15 | 1665 | 9.84 (7.84) | 0.19 | 1395 | 10.07 (8.00) | 0.21 | 1279 | 10.45 (8.29) | 0.23 | 1374 | 10 (8.02) | 0.22 |
| Self Harm | 2691 | 1.23 (0.72) | 0.01 | 1646 | 1.06 (0.32) | 0.01 | 1377 | 1.06 (0.32) | 0.01 | 1262 | 0.07 (0.37) | 0.01 | 1361 | 1.07 (0.37) | 0.01 |

(c) Males

| | T1 | | | T2 | | | T3 | | | T4 | | | T5 | | |
|---|---|---|---|---|---|---|---|---|---|---|---|---|---|---|---|
| | **N** | **Mean (SD)** | **SE** | **N** | **Mean (SD)** | **SE** | **N** | **Mean (SD)** | **SE** | **N** | **Mean (SD)** | **SE** | **N** | **Mean (SD)** | **SE** |
| Emotional problems | 1801 | 2.80 (2.49) | 0.06 | 772 | 2.03 (2.27) | 0.08 | 611 | 2.28 (2.32) | 0.09 | 551 | 2.46 (2.47) | 0.11 | 566 | 2.46 (2.39) | 0.1 |
| Conduct problems | 1802 | 1.63 (1.31) | 0.03 | 772 | 1.47 (1.22) | 0.04 | 611 | 1.40 (1.07) | 0.04 | 551 | 1.39 (1.02) | 0.04 | 566 | 1.34 (0.99) | 0.04 |
| Hyperactivity | 1801 | 3.45 (2.22) | 0.05 | 772 | 4.06 (2.25) | 0.08 | 611 | 4.04 (2.24) | 0.09 | 551 | 3.86 (2.18) | 0.09 | 566 | 3.77 (2.22) | 0.09 |
| Peer problems | 1801 | 2.19 (1.73) | 0.04 | 772 | 2.34 (1.60) | 0.06 | 611 | 2.46 (1.63) | 0.07 | 551 | 2.32 (1.58) | 0.07 | 566 | 2.55 (1.66) | 0.07 |
| Prosocial behaviour | 1802 | 7.02 (2.02) | 0.05 | 772 | 6.18 (1.99) | 0.07 | 611 | 6.15 (2.02) | 0.08 | 551 | 6.17 (1.99) | 0.08 | 566 | 6.15 (2.03) | 0.09 |
| Behaviour problems | 1801 | 10.08 (5.54) | 0.13 | 772 | 9.92 (5.25) | 0.19 | 611 | 10.19 (5.19) | 0.21 | 551 | 10.03 (5.29) | 0.23 | 566 | 10.11 (5.18) | 0.22 |
| Depression | 1800 | 3.75 (3.67) | 0.09 | 772 | 3.37 (3.41) | 0.12 | 611 | 3.39 (3.40) | 0.14 | 551 | 3.48 (3.62) | 0.15 | 566 | 3.49 (3.56) | 0.15 |
| General anxiety | 1542 | 6.04 (6.86) | 0.17 | 772 | 6.33 (6.53) | 0.24 | 611 | 6.36 (6.52) | 0.26 | 551 | 6.96 (6.76) | 0.29 | 566 | 6.49 (6.44) | 0.27 |
| Self Harm | 1538 | 1.14 (0.56) | 0.01 | 764 | 1.02 (0.18) | 0.01 | 604 | 1.04 (0.26) | 0.01 | 547 | 1.03 (0.23) | 0.01 | 560 | 1.04 (0.22) | 0.01 |

(d)

| | Time | | Sex | | Time * Sex | |
|---|---|---|---|---|---|---|
| | | | **F** | **Partial Eta squared** | **F** | **Partial Eta squared** |
| Emotional problems | 14.92** | 0.02 | 97.48** | 0.09 | 1.07 | 0 |
| Conduct problems | 2.28 | 0 | 2.56 | 0 | 1.24 | 0 |
| Hyperactivity | 53.43** | 0.05 | 5.71* | 0.01 | 1.96 | 0 |
| Peer problems | 11.67** | 0.01 | 0.25 | 0 | 0.27 | 0 |
| Prosocial behaviour | 69.62 | 0.07 | 96.94** | 0.09 | 0.41 | 0 |
| Behaviour problems | 5.03** | 0.01 | 29.35** | 0.03 | 2.06 | 0 |
| Depression | 0.2 | 0 | 38.00** | 0.04 | 1.07 | 0 |
| General anxiety | 10.32** | 1 | 0.40** | 0.06 | 2.88* | 0 |
| Self Harm | 26.75** | 0.03 | 1.2 | 0 | 1.13 | 0 |

Table S2. Descriptive statistics for CRISIS battery one randomly selected twin per pair

| **Suspected COVID-19 Infection** | T2 | T3 | T4 | T5 |
|---|---|---|---|---|
| yes, have positive test N (%) | 12 (0.5%) | 32 (1.6%) | 34 (1.8%) | 65 (3.2%) |
| yes, medical diagnosis but no test N (%) | 18 (0.7%) | 16 (0.8%) | 7 (0.4%) | 12 (0.6%) |
| yes, symptoms but no diagnosis N (%) | 368 (14.8%) | 208 (10.2%) | 178 (9.5%) | 108 (5.4%) |
| no symptoms or signs N (%) | 2095 (84%) | 1793 (87.5%) | 1655 (88.3%) | 1826 (90.8%) |

| **Covid Symptoms (% of the sample)** | | | | |
|---|---|---|---|---|
| **Fever** | T2 (only asked yes/no) | T3 | T4 | T5 |
| no | 92.9 | 90.5 | 91.3 | 92.4 |
| yes slightly | | 4.7 | 4.9 | 4.5 |
| yes moderately | | 2.5 | 2.1 | 1.7 |
| yes very | | 1.7 | 1.3 | 1.1 |
| yes extremely | | 0.6 | 0.5 | 0.3 |

| | | | | |
|---|---|---|---|---|
| **Cough** | T2 (only asked yes/no) | T3 | T4 | T5 |
| no | 85 | 80.9 | 77.3 | 82.6 |
| yes slightly | | 13.6 | 16.1 | 12.8 |
| yes moderately | | 3.2 | 3.8 | 2.8 |
| yes very | | 1.5 | 2.2 | 1.4 |
| yes extremely | | 0.8 | 0.5 | 0.4 |
| **Short breath** | T2 (only asked yes/no) | T3 | T4 | T5 |
| no | 92.1 | 85.5 | 87.1 | 87.2 |
| yes slightly | | 9.7 | 9.1 | 8.6 |
| yes moderately | | 3.2 | 2.3 | 2.8 |
| yes very | | 1.2 | 1.1 | 1 |
| yes extremely | | 0.5 | 0.4 | 0.4 |
| **Sore throat** | T2 (only asked yes/no) | T3 | T4 | T5 |
| no | 82.6 | 77 | 68.6 | 77.6 |
| yes slightly | | 15.8 | 22.1 | 16.3 |
| yes moderately | | 4.6 | 6 | 4.5 |
| yes very | | 1.7 | 2.7 | 1 |
| yes extremely | | 0.9 | 0.6 | 0.6 |
| **Fatigue** | T2 (only asked yes/no) | T3 | T4 | T5 |
| no | 82 | 64.7 | 62.9 | 60.1 |
| yes slightly | | 18.6 | 20.2 | 21.5 |
| yes moderately | | 9 | 9.6 | 10.8 |
| yes very | | 5.1 | 5.4 | 5.4 |

| | | | | |
|---|---|---|---|---|
| yes extremely | | 2.6 | 1.9 | 2.2 |

| **Taste or smell loss** | T2 (only asked yes/no) | T3 | T4 | T5 |
|---|---|---|---|---|
| no | 94.2 | 92.2 | 93.7 | 94.3 |
| yes slightly | | 2.6 | 2.8 | 2.1 |
| yes moderately | | 1.5 | 1 | 0.8 |
| yes very | | 1.4 | 1 | 1 |
| yes extremely | | 2.2 | 1.5 | 1.6 |

| **Eye infection** | T2 (item not used) | T3 | T4 | T5 |
|---|---|---|---|---|
| no | | 97.2 | 97.4 | 96.8 |
| yes slightly | | 2 | 1.8 | 2.1 |
| yes moderately | | 0.8 | 0.4 | 1 |
| yes very | | | 0.4 | |

| **Sleep problems** | T2 (item not used) | T3 (item not used) | T4 (item not used) | T5 |
|---|---|---|---|---|
| no | | | | 56.3 |
| yes slightly | | | | 23.4 |
| yes moderately | | | | 11.9 |
| yes very | | | | 5.5 |
| yes extremely | | | | 2.9 |

| **Memory problems** | T2 (item not used) | T3 (item not used) | T4 (item not used) | T5 |
|---|---|---|---|---|
| no | | | | 83.4 |
| yes slightly | | | | 10.6 |
| yes moderately | | | | 3.7 |
| yes very | | | | 1.6 |

| | | | | |
|---|---|---|---|---|
| yes extremely | | | | 0.7 |

| Concentration problems | T2 (item not used) | T3 (item not used) | T4 (item not used) | T5 |
|---|---|---|---|---|
| no | | | | 73.5 |
| yes slightly | | | | 16.7 |
| yes moderately | | | | 6 |
| yes very | | | | 2.4 |
| yes extremely | | | | 1.4 |

| Concentration problems | T2 (item not used) | T3 (item not used) | T4 (item not used) | T5 |
|---|---|---|---|---|
| no | | | | 73.5 |
| yes slightly | | | | 16.7 |
| yes moderately | | | | 6 |
| yes very | | | | 2.4 |
| yes extremely | | | | 1.4 |

| Muscle and joint pain | T2 (item not used) | T3 (item not used) | T4 (item not used) | T5 |
|---|---|---|---|---|
| no | | | | 73.5 |
| yes slightly | | | | 16.7 |
| yes moderately | | | | 6 |
| yes very | | | | 2.4 |
| yes extremely | | | | 1.4 |

| Diarrhoea | T2 (item not used) | T3 (item not used) | T4 (item not used) | T5 |
|---|---|---|---|---|
| no | | | | 88.9 |
| yes slightly | | | | 7.8 |
| yes moderately | | | | 2.1 |

| | | | | |
|---|---|---|---|---|
| yes very | | | | 0.7 |
| yes extremely | | | | 0.5 |
| **Stomach pains** | T2 (item not used) | T3 (item not used) | T4 (item not used) | T5 |
| no | | | | 82.2 |
| yes slightly | | | | 11.9 |
| yes moderately | | | | 4 |
| yes very | | | | 1.2 |
| yes extremely | | | | 0.6 |
| **Voice changes** | T2 (item not used) | T3 (item not used) | T4 (item not used) | T5 |
| no | | | | 96.3 |
| yes slightly | | | | 2.7 |
| yes moderately | | | | 0.3 |
| yes very | | | | 0.7 |
| yes extremely | | | | |
| **Hair loss** | T2 (item not used) | T3 (item not used) | T4 (item not used) | T5 |
| no | | | | 94.2 |
| yes slightly | | | | 4.2 |
| yes moderately | | | | 0.8 |
| yes very | | | | 0.8 |
| yes extremely | | | | |
| **Racing heart** | T2 (item not used) | T3 (item not used) | T4 (item not used) | T5 |
| no | | | | 88.6 |
| yes slightly | | | | 7.6 |

| | | | | |
|---|---|---|---|---|
| yes moderately | | | | 2.8 |
| yes very | | | | 0.8 |
| yes extremely | | | | 0.2 |
| **Sweating** | T2 (item not used) | T3 (item not used) | T4 (item not used) | T5 |
| no | | | | 91.1 |
| yes slightly | | | | 5.9 |
| yes moderately | | | | 1.8 |
| yes very | | | | 0.8 |
| yes extremely | | | | 0.3 |

| **How long symptoms lasted (only used in T5) (% of the sample)** | |
|---|---|
| no symptoms | 74.2 |
| still have symptoms | 1.1 |
| less than 1 week | 10.8 |
| 1 to 2 weeks | 7.1 |
| 2 to 4 weeks | 4.3 |
| 1 to 3 months | 1.3 |
| more than 3 months | 1 |

| **Vaccination (only used in T5) (% of the sample)** | |
|---|---|
| had vaccination | 24.6 |
| will be vaccinated soon | 2.6 |
| offered but not accepted | 1.1 |
| not yet offered but would accept | 66.7 |

| | |
|---|---|
| not yet offered but would not accept | 5 |

| **Access to garden (% of the sample)** | T2 | T3 | T4 | T5 |
|---|---|---|---|---|
| yes | 82.4 | 83.3 | 80.8 | 82.2 |

| **Number of people living with (% of the sample)** | T2 | T3 | T4 | T5 |
|---|---|---|---|---|
| 0 | 4.5 | 5.4 | 6.1 | 7.2 |
| 1 | 26.7 | 28.3 | 32.7 | 31.9 |
| 2 | 23.9 | 24.8 | 23.8 | 26 |
| 3 | 22 | 22.5 | 21.5 | 18.5 |
| 4 | 14.1 | 10.7 | 9.3 | 9.7 |
| 5 | 5.2 | 5 | 3.9 | 3.9 |
| more than 5 | 3.6 | 3.2 | 2.7 | 2.9 |

| **Covid Family effect (Mean (SD); range 0-7)** | T2 | T3 | T4 | T5 |
|---|---|---|---|---|
| | 0.97 (0.18) | 0.87 (1.21) | 0.95 (1.28) | 1.56 (0.75) |

Note: Caluclated as a sum score of family member fallen ill physically, family member hospitalised, put into isolation with or without symptoms, lost a job, reduced ability to ean money, passed away

| **Worries (Mean (SD); range 0-4)** | | | | |
|---|---|---|---|---|
| | T2 | T3 | T4 | T5 |
| Worries about COVID | 1.90 (0.75) | 1.67 (0.73) | 1.73 (.77) | 1.59 (0.75) |
| Mental Helath worries | 1.77 (1.2) | 1.66 (1.18) | 1.74 (1.19) | 1.71 (1.17) |
| Financial Worries | 1 (1.06) | 0.84 (0.98) | 0.87 (1.04) | 0.74 (1.0) |

Table S3. Longitudinal trends in mental health. Results of Latent Growth Curve model.

| **Emotional problems** | | | | | | | | | | | | |
|---|---|---|---|---|---|---|---|---|---|---|---|---|
| | χ2 | *p* | CFI | TLI | RMSA (95% CI) | SRMR | Intercept (SE) | Variance intercept | Slope | Variance slope | Quadratic | Variance quadratic |
| Model 1 | 294.23 | 0 | 0.952 | 0.958 | 0.048 (0.043-0.053) | 0.068 | 4.008 (0.042)** | 4.107 (0.112)** | | | | |
| Model 2 | 152.57 | 0 | 0.976 | 0.972 | 0.039 (0.033-0.044) | 0.037 | 4.111 (0.048)** | 3.619 (0.320)** | (-0.059) (0.01)** | 0.095 (0.03)* | | |
| Model 3 | 23.828 | 0 | 0.993 | 0.986 | 0.027 (0.020-0.035) | 0.018 | 4.142 (0.048)** | 4.175 (2.812)** | (-0.333) (0.067)** | 2.446 (1.321) | 0.088 (0.021)** | 0.249 (0.048)** |
| ANOVA Model 1-Model 2 | | | | | | | | | | | | |
| | Df | *AIC* | BIC | Chisq | Chisq diff | Df diff | Pr(>Chisq) | | | | | |
| Model 2 | 13 | 57164 | 57246 | 152.57 | | | | | | | | |
| Model 1 | 17 | 57297 | 57352 | 294.23 | 141.66 | 4 | < 2.2e-16 ** | | | | | |
| ANOVA Model 2-Model 3 | | | | | | | | | | | | |
| | Df | *AIC* | BIC | Chisq | Chisq diff | Df diff | Pr(>Chisq) | | | | | |
| Model 3 | 8 | 39863 | 39980 | 23.828 | | | | | | | | |
| Model 2 | 13 | 39913 | 39996 | 84.176 | 60.348 | 5 | 1.03e-11 ** | | | | | |

| **Conduct problems** |
|---|

| | χ2 | *p* | CFI | TLI | RMSA (95% CI) | SRMR | Intercept (SE) | Variance intercept | Slope | Variance slope | Quadratic | Variance quadratic |
|---|---|---|---|---|---|---|---|---|---|---|---|---|
| Model 1 | 129.616 | 0 | 0.949 | 0.955 | 0.030 (0.026-0.035) | 0.052 | 1.558 (0.018)** | 0.642 (0.023)** | | | | |
| Model 2 | 84.176 | 0 | 0.968 | 0.963 | 0.028 (0.022-0.033) | 0.043 | 1.579 (0.024)** | 0.471 (0.102) ** | (-0.011) (0.009) | 0.00 (0.013) | | |
| Model 3 | 23.828 | 0.002 | 0.993 | 0.986 | 0.017 (0.009-0.025) | 0.021 | (0.024) ** | 0.807 (0.947) | 0.079 (0.038)* | 0.695 (0.452) | (-0.029) (0.012)* | 0.059 (0.016)** |
| ANOVA Model 1-Model 2 | | | | | | | | | | | | |
| | Df | *AIC* | BIC | Chisq | Chisq diff | Df diff | Pr(>Chisq) | | | | | |
| Model 2 | 13 | *39913* | 39996 | 84.176 | | | | | | | | |
| Model 1 | 17 | 39951 | 40006 | 129.616 | 45.44 | 4 | 3.221e-09 ** | | | | | |
| ANOVA Model 2-Model 3 | | | | | | | | | | | | |
| | Df | *AIC* | BIC | Chisq | Chisq diff | Df diff | Pr(>Chisq) | | | | | |
| Model 3 | 8 | *39863* | 39980 | 23.828 | | | | | | | | |
| Model 2 | 13 | 39913 | 39996 | 84.176 | 60.348 | 5 | 1.03e-11 ** | | | | | |

| **Hyperactivity** | | | | | | | | | | | | |
|---|---|---|---|---|---|---|---|---|---|---|---|---|
| | χ2 | *p* | CFI | TLI | RMSA (95% CI) | SRMR | Intercept (SE) | Variance intercept | Slope | Variance slope | Quadratic | Variance quadratic |
| Model 1 | 788.21 | 0 | 0.792 | 0.817 | 0.080 (0.075-0.085) | 0.105 | 3.868 (0.036)** | 2.605 (0.078)** | | | | |
| Model 2 | 218.62 | 0 | 0.945 | 0.936 | 0.047 (0.042-0.053) | 0.048 | 3.4 (0.04)** | 1.591 (0.28)** | 0.269 (0.016)** | 0.013 (0.035) | | |

| | | | | | | | | | | | | |
|---|---|---|---|---|---|---|---|---|---|---|---|---|
| Model 3 | 14.765 | 0 | 0.998 | 0.997 | 0.011 (0.00-0.19) | 0.011 | 3.318 (0.040)** | (*0.251)(2.399) | 0.976 (0.063)** | 0.449 (1.148) | (-0.231) (0.020)** | 0.127 (0.044)* |
| ANOVA Model 1-Model 2 | | | | | | | | | | | | |
| | Df | *AIC* | BIC | Chisq | Chisq diff | Df diff | Pr(>Chisq) | | | | | |
| Model 2 | 13 | *54401* | 54483 | 218.62 | | | | | | | | |
| Model 1 | 17 | 54962 | 55017 | 788.21 | 569.59 | 4 | < 2.2e-16 ** | | | | | |
| ANOVA Model 2-Model 3 | | | | | | | | | | | | |
| | Df | *AIC* | BIC | Chisq | Chisq diff | Df diff | Pr(>Chisq) | | | | | |
| Model 3 | 8 | 54207 | 54324 | 14.765 | | | | | | | | |
| Model 2 | 13 | 54401 | 54483 | 2108.62 | 203.85 | 5 | < 2.2e-16 ** | | | | | |

| **Peer Problems** | | | | | | | | | | | | |
|---|---|---|---|---|---|---|---|---|---|---|---|---|
| | χ2 | *p* | CFI | TLI | RMSA (95% CI) | SRMR | Intercept (SE) | Variance intercept | Slope | Variance slope | Quadratic | Variance quadratic |
| Model 1 | 177.81 | 0 | 0.968 | 0.971 | 0.036 (0.032-0.041) | 0.046 | 2.296 (0.028)** | 1.772 (0.049)** | | | | |
| Model 2 | 81.795 | 0 | 0.986 | 0.984 | 0.027 (0.022-0.033) | 0.022 | 2.21 (0.033)** | 1.815 (0.146)** | 0.049 (0.011)** | 0.043 (0.017)* | | |
| Model 3 | 24.498 | 0.002 | 0.997 | 0.994 | 0.017 (0.010-0.025) | 0.014 | 2.235 (0.033)** | 3.046 (1.284)* | (-0.149) (0.043)** | 1.248 (0.605)* | 0.064 (0.013)** | 0.090 (0.02)** |
| ANOVA Model 1-Model 2 | | | | | | | | | | | | |
| | Df | *AIC* | BIC | Chisq | Chisq diff | Df diff | Pr(>Chisq) | | | | | |

| | | | | | | | |
|---|---|---|---|---|---|---|---|
| Model 2 | 13 | *43273* | 46355 | 81.795 | | | |
| Model 1 | 17 | 46361 | 46416 | 177.81 | 96.015 | 4 | < 2.2e-16 ** |
| ANOVA Model 2-Model 3 | | | | | | | |
| | Df | *AIC* | BIC | Chisq | Chisq diff | Df diff | Pr(>Chisq) |
| Model 3 | 3 | *46225* | 46342 | 24.498 | | | |
| Model 2 | 18 | 46273 | 46355 | 81.795 | 52.297 | 5 | 4.392e-11 ** |

| **Prosocial Behaviour** | | | | | | | | | | | |
|---|---|---|---|---|---|---|---|---|---|---|---|
| | χ2 | *p* | CFI | TLI | RMSA (95% CI) | SRMR | Intercept (SE) | Variance intercept | Slope | Variance slope | Quadratic | Variance quadratic |
| Model 1 | 900.506 | 0 | 0.8 | 0.823 | 0.085 (0.081-0.090) | 0.111 | 7.546 (0.030)** | 1.797 (0.054)** | | | | |
| Model 2 | 85.494 | 0 | 0.984 | 0.981 | 0.028 (0.022-0.034) | 0.032 | 7.986 (0.033)** | 1.753 (0.191)** | (-0.250) (0.013)** | 0.088 (0.024)** | | |
| Model 3 | 9.0897 | 0 | 1 | 1 | 0.004 (0.00-0.015) | 0.012 | 8.01 (0.033)** | 1.406 (1.737) | (-0.520) (0.052)** | 0.755 )0.827 | 0.089 (0.016)** | 0.086 (0.031)* |
| ANOVA Model 1-Model 2 | | | | | | | | | | | | |
| | Df | *AIC* | BIC | Chisq | Chisq diff | Df diff | Pr(>Chisq) | | | | | |
| Model 2 | 13 | *49697* | 49779 | 85.494 | | | | | | | | |
| Model 1 | 17 | 50504 | 50559 | 900.506 | 845.01 | 4 | < 2.2e-16 ** | | | | | |
| ANOVA Model 2-Model 3 | | | | | | | | | | | | |
| | Df | *AIC* | BIC | Chisq | Chisq diff | Df diff | Pr(>Chisq) | | | | | |

| | | | | | | | |
|---|---|---|---|---|---|---|---|
| Model 3 | 8 | *49630* | 49747 | 9.0897 | | | |
| Model 2 | 13 | 49697 | 49779 | 85.4937 | 76.404 | 5 | 4.737e-15 ** |

| **Behavoouràl Problems** | | | | | | | | | | | |
|---|---|---|---|---|---|---|---|---|---|---|---|
| | χ2 | *p* | CFI | TLI | RMSA (95% CI) | SRMR | Intercept (SE) | Variance intercept | Slope | Variance slope | Quadratic | Variance quadratic |
| Model 1 | 235.75 | 0 | 0.964 | 0.969 | 0.042 (0.038-0.047) | 0.061 | 11.754 (0.093)** | 21.20 (0.555)** | | | | |
| Model 2 | 100.8 | 0 | 0.986 | 0.984 | 0.031 (0.025-0.036) | 0.033 | 11.307 (0.105)** | 18.933 (1.410)** | 0.260 (0.035)** | 0.284 (0.167) | | |
| Model 3 | 30.608 | 0 | 0.996 | 0.993 | 0.02 (0.013-0.028) | 0.013 | 11.257 (0.106)** | 5.755 (11.808) | 0.603 (0.138)** | 3.508 (5.569) | (-0.114)(0.043)* | 0.938 (0.20)** |
| ANOVA Model 1-Model 2 | | | | | | | | | | | |
| | Df | *AIC* | BIC | Chisq | Chisq diff | Df diff | Pr(>Chisq) | | | | |
| Model 2 | 13 | *76334* | 76416 | 100.8 | | | | | | | |
| Model 1 | 17 | 76461 | 76516 | 235.75 | 134.95 | 4 | < 2.2e-16 ** | | | | |
| ANOVA Model 2-Model 3 | | | | | | | | | | | |
| | Df | *AIC* | BIC | Chisq | Chisq diff | Df diff | Pr(>Chisq) | | | | |
| Model 3 | 8 | *76273* | 73390 | 30.608 | | | | | | | |
| Model 2 | 13 | 76334 | 76416 | 100.799 | 70.192 | 5 | 9.349e-14 ** | | | | |

| **Depressive symptoms** |
|---|

| | χ2 | *p* | CFI | TLI | RMSA (95% CI) | SRMR | Intercept (SE) | Variance intercept | Slope | Variance slope | Quadratic | Variance quadratic |
|---|---|---|---|---|---|---|---|---|---|---|---|---|
| Model 1 | 159.492 | 0 | 0.974 | 0.977 | 0.034 (0.030-0.039) | 0.059 | 5.020 (0.065)** | 9.817 (0.265)** | | | | |
| Model 2 | 80.181 | 0 | 0.988 | 0.986 | 0.027 (0.021-0.033) | 0.033 | 4.945 (0.073)** | 7.806 (0.76)** | 0.044 (0.025) | 0.061 (0.092) | | |
| Model 3 | 26.005 | 0 | 0.997 | 0.994 | 0.018 (0.010-0.026) | 0.017 | 4.918 (0.074)** | 1.571 (6.488) | 0.259 (0.102)* | 1.816 (3.071) | (-0.071) (0.032)* | 0.475 (0.111)** |
| ANOVA Model 1-Model 2 | | | | | | | | | | | | |
| | Df | *AIC* | BIC | Chisq | Chisq diff | Df diff | Pr(>Chisq) | | | | | |
| Model 2 | 13 | *68022* | 68104 | 80.181 | | | | | | | | |
| Model 1 | 17 | 68093 | 68148 | 159.492 | 79.31 | 4 | 2.438e-16 ** | | | | | |
| ANOVA Model 2-Model 3 | | | | | | | | | | | | |
| | Df | *AIC* | BIC | Chisq | Chisq diff | Df diff | Pr(>Chisq) | | | | | |
| Model 3 | 8 | *67978* | 68095 | 26.005 | | | | | | | | |
| Model 2 | 13 | 68022 | 68104 | 80.181 | 54.177 | 5 | 1.928e-10 ** | | | | | |

| **General anxiety** | | | | | | | | | | | | |
|---|---|---|---|---|---|---|---|---|---|---|---|---|
| | χ2 | *p* | CFI | TLI | RMSA (95% CI) | SRMR | Intercept (SE) | Variance intercept | Slope | Variance slope | Quadratic | Variance quadratic |
| Model 1 | 472.228 | 0 | 0.926 | 0.934 | 0.061 (0.057-0.066) | 0.093 | 9.503 (0.127)** | 36.964 (0.991)** | | | | |
| Model 2 | 121.342 | 0 | 0.982 | 0.98 | 0.034 (0.029-0.040) | 0.035 | 8.403 (0.139)** | 30.439 (2.422)** | 0.595 (0.048)** | 0.906 (0.288)* | | |

| | | | | | | | | | | | | |
|---|---|---|---|---|---|---|---|---|---|---|---|---|
| Model 3 | 38.715 | 0 | 0.995 | 0.991 | 0.023 (0.016-0.031 | 0.016 | 8.287 )0.140)** | (-12.078) (21.197) | 1.374 (0.179)** | (-2.812) (9.952) | (-0.254) (0.055)** | 1.363 (0.652) ** |
| ANOVA Model 1-Model 2 | | | | | | | | | | | | |
| | Df | *AIC* | BIC | Chisq | Chisq diff | Df diff | Pr(>Chisq) | | | | | |
| Model 2 | 13 | *80199* | 80281 | 121.34 | | | | | | | | |
| Model 1 | 17 | 80542 | 80597 | 472.23 | 350.89 | 4 | < 2.2e-16 ** | | | | | |
| ANOVA Model 2-Model 3 | | | | | | | | | | | | |
| | Df | *AIC* | BIC | Chisq | Chisq diff | Df diff | Pr(>Chisq) | | | | | |
| Model 3 | 8 | *80126* | 80243 | 38.715 | | | | | | | | |
| Model 2 | 13 | 80199 | 80281 | 121.342 | 82.627 | 5 | 2.366e-16 ** | | | | | |

Note: ** p <0.001; * p<0.05

Table S4. Model-fit statistics for the Latent Profile Analyses

| Emotional problems | | | | | | | | | | | | | |
|---|---|---|---|---|---|---|---|---|---|---|---|---|---|
| Model | Classes | LogLik | AIC | BIC | SABIC | Entropy | prob_min | prob_max | n_min | n_max | BLRT_val | BLRT_p | delta_BIC |
| 1.00 | 2.00 | -29720.11 | 59472.21 | 59576.23 | 59525.39 | 0.87 | 0.95 | 0.97 | 0.41 | 0.59 | 10310.83 | 0.01 | 0.00 |
| 1.00 | 3.00 | -28244.35 | 56532.69 | 56675.72 | 56605.81 | 0.83 | 0.91 | 0.94 | 0.20 | 0.41 | 2951.41 | 0.01 | -2900.52 |
| 1.00 | 4.00 | -27774.63 | 55605.27 | 55787.30 | 55698.33 | 0.79 | 0.86 | 0.90 | 0.12 | 0.36 | 942.25 | 0.01 | -888.42 |
| 1.00 | 5.00 | -27637.47 | 55342.94 | 55563.98 | 55455.94 | 0.74 | 0.79 | 0.86 | 0.09 | 0.31 | 264.38 | 0.01 | -223.32 |

| | | | | | | | | | | | | | |
|---|---|---|---|---|---|---|---|---|---|---|---|---|---|
| 1.00 | 6.00 | -27581.40 | 55242.79 | 55502.84 | 55375.73 | 0.71 | 0.66 | 0.84 | 0.05 | 0.31 | 176.12 | 0.01 | -61.14 |
| 1.00 | 7.00 | -27513.32 | 55118.64 | 55417.70 | 55271.53 | 0.69 | 0.53 | 0.83 | 0.04 | 0.29 | 116.36 | 0.01 | -85.14 |
| Conduct problems | | | | | | | | | | | | | |
| Model | Classes | LogLik | AIC | BIC | SABIC | Entropy | prob_min | prob_max | n_min | n_max | BLRT_val | BLRT_p | delta_BIC |
| 1.00 | 2.00 | -32366.19 | 64764.39 | 64868.41 | 64817.57 | 0.80 | 0.88 | 0.97 | 0.27 | 0.73 | 4904.92 | 0.01 | 0.00 |
| 1.00 | 3.00 | -31795.25 | 63634.51 | 63777.54 | 63707.63 | 0.75 | 0.84 | 0.91 | 0.07 | 0.52 | 1142.83 | 0.01 | -1090.87 |
| 1.00 | 4.00 | -31565.37 | 63186.75 | 63368.79 | 63279.81 | 0.74 | 0.64 | 0.91 | 0.04 | 0.60 | 469.67 | 0.01 | -408.75 |
| 1.00 | 5.00 | -31480.27 | 63028.54 | 63249.59 | 63141.55 | 0.70 | 0.58 | 0.86 | 0.02 | 0.48 | 10.26 | 0.01 | -119.20 |
| 1.00 | 6.00 | -31432.73 | 62945.46 | 63205.52 | 63078.41 | 0.69 | 0.53 | 0.86 | 0.02 | 0.42 | 212.37 | 0.01 | -44.07 |
| 1.00 | 7.00 | -31410.15 | 62912.29 | 63211.36 | 63065.19 | 0.62 | 0.09 | 0.89 | 0.01 | 0.48 | 131.32 | 0.01 | 5.84 |
| Hyperactivity | | | | | | | | | | | | | |
| Model | Classes | LogLik | AIC | BIC | SABIC | Entropy | prob_min | prob_max | n_min | n_max | BLRT_val | BLRT_p | delta_BIC |
| 1.00 | 2.00 | -31274.83 | 62581.65 | 62685.68 | 62634.83 | 0.79 | 0.94 | 0.94 | 0.47 | 0.53 | 7119.33 | 0.01 | 0.00 |
| 1.00 | 3.00 | -30114.50 | 60273.00 | 60416.03 | 60346.12 | 0.79 | 0.88 | 0.91 | 0.21 | 0.52 | 2320.58 | 0.01 | -2269.65 |
| 1.00 | 4.00 | -29715.71 | 59487.41 | 59669.45 | 59580.47 | 0.76 | 0.84 | 0.87 | 0.09 | 0.40 | 792.52 | 0.01 | -746.58 |
| 1.00 | 5.00 | -29578.19 | 59224.37 | 59445.41 | 59337.37 | 0.73 | 0.79 | 0.83 | 0.06 | 0.36 | 279.26 | 0.01 | -224.03 |
| 1.00 | 6.00 | -29553.48 | 59186.96 | 59447.01 | 59319.90 | 0.65 | 0.58 | 0.84 | 0.05 | 0.29 | 62.40 | 0.01 | 1.59 |

| | | | | | | | | | | | | | |
|---|---|---|---|---|---|---|---|---|---|---|---|---|---|
| 1.00 | 7.00 | -29533.85 | 59159.70 | 59458.76 | 59312.59 | 0.64 | 0.18 | 0.84 | 0.03 | 0.30 | 10.31 | 0.03 | 11.75 |
| Peer problems | | | | | | | | | | | | | |
| Model | Classes | LogLik | AIC | BIC | SABIC | Entropy | prob_min | prob_max | n_min | n_max | BLRT_val | BLRT_p | delta_BIC |
| 1.00 | 2.00 | -30509.85 | 61051.69 | 61155.71 | 61104.87 | 0.81 | 0.94 | 0.95 | 0.44 | 0.56 | 8083.25 | 0.01 | 0.00 |
| 1.00 | 3.00 | -28930.15 | 57904.30 | 58047.33 | 57977.42 | 0.83 | 0.89 | 0.93 | 0.14 | 0.50 | 3159.32 | 0.01 | -3108.38 |
| 1.00 | 4.00 | -28464.88 | 56985.76 | 57167.80 | 57078.82 | 0.78 | 0.86 | 0.89 | 0.07 | 0.40 | 921.11 | 0.01 | -879.53 |
| 1.00 | 5.00 | -28252.00 | 56572.00 | 56793.04 | 56685.00 | 0.76 | 0.81 | 0.88 | 0.03 | 0.37 | 426.69 | 0.01 | -374.75 |
| 1.00 | 6.00 | -28220.24 | 56520.47 | 56780.52 | 56653.42 | 0.72 | 0.38 | 0.87 | 0.03 | 0.34 | 75.07 | 0.01 | -12.52 |
| 1.00 | 7.00 | -28096.82 | 56285.64 | 56584.70 | 56438.53 | 0.76 | 0.45 | 0.88 | 0.01 | 0.35 | 183.71 | 0.01 | -195.82 |
| Prosocial behaviour | | | | | | | | | | | | | |
| Model | Classes | LogLik | AIC | BIC | SABIC | Entropy | prob_min | prob_max | n_min | n_max | BLRT_val | BLRT_p | delta_BIC |
| 1.00 | 2.00 | -30898.87 | 61829.74 | 61933.76 | 61882.92 | 0.80 | 0.94 | 0.95 | 0.46 | 0.54 | 7615.25 | 0.01 | 0.00 |
| 1.00 | 3.00 | -29709.19 | 59462.39 | 59605.41 | 59535.50 | 0.80 | 0.89 | 0.91 | 0.19 | 0.48 | 2379.35 | 0.01 | -2328.35 |
| 1.00 | 4.00 | -29393.96 | 58843.93 | 59025.96 | 58936.99 | 0.75 | 0.84 | 0.87 | 0.09 | 0.38 | 626.65 | 0.01 | -579.45 |
| 1.00 | 5.00 | -29274.90 | 58617.79 | 58838.83 | 58730.79 | 0.72 | 0.77 | 0.83 | 0.04 | 0.35 | 242.04 | 0.01 | -187.13 |
| 1.00 | 6.00 | -29245.69 | 58571.37 | 58831.42 | 58704.32 | 0.66 | 0.56 | 0.84 | 0.04 | 0.33 | 59.91 | 0.01 | -7.41 |
| 1.00 | 7.00 | -29195.67 | 58483.33 | 58782.39 | 58636.22 | 0.66 | 0.48 | 0.82 | 0.02 | 0.27 | 101.72 | 0.01 | -49.03 |

| | | | | | | | | | | | | | |
|---|---|---|---|---|---|---|---|---|---|---|---|---|---|
| Behaviour problems | | | | | | | | | | | | | |
| Model | Classes | LogLik | AIC | BIC | SABIC | Entropy | prob_min | prob_max | n_min | n_max | BLRT_val | BLRT_p | delta_BIC |
| 1.00 | 2.00 | -29852.03 | 59736.07 | 59840.09 | 59789.25 | 0.85 | 0.95 | 0.96 | 0.42 | 0.58 | 10060.56 | 0.01 | 0.00 |
| 1.00 | 3.00 | -27991.21 | 56026.41 | 56169.44 | 56099.53 | 0.84 | 0.92 | 0.93 | 0.19 | 0.47 | 3721.77 | 0.01 | -3670.65 |
| 1.00 | 4.00 | -27090.46 | 54236.93 | 54418.96 | 54329.99 | 0.84 | 0.89 | 0.91 | 0.09 | 0.41 | 1801.00 | 0.01 | -1750.48 |
| 1.00 | 5.00 | -26707.53 | 53483.06 | 53704.11 | 53596.07 | 0.82 | 0.87 | 0.90 | 0.05 | 0.34 | 765.55 | 0.01 | -714.86 |
| 1.00 | 6.00 | -26556.13 | 53192.26 | 53452.31 | 53325.20 | 0.79 | 0.80 | 0.89 | 0.04 | 0.30 | 70.73 | 0.01 | -251.80 |
| 1.00 | 7.00 | -26510.99 | 53113.98 | 53413.04 | 53266.87 | 0.75 | 0.49 | 0.90 | 0.04 | 0.31 | 319.10 | 0.01 | -39.27 |
| Depression | | | | | | | | | | | | | |
| Model | Classes | LogLik | AIC | BIC | SABIC | Entropy | prob_min | prob_max | n_min | n_max | BLRT_val | BLRT_p | delta_BIC |
| 1.00 | 2.00 | -29903.06 | 59838.11 | 59942.13 | 59891.29 | 0.87 | 0.94 | 0.97 | 0.35 | 0.65 | 9725.03 | 0.01 | 0.00 |
| 1.00 | 3.00 | -28407.49 | 56858.99 | 57002.02 | 56932.11 | 0.83 | 0.91 | 0.94 | 0.15 | 0.43 | 2990.86 | 0.01 | -2940.11 |
| 1.00 | 4.00 | -27864.25 | 55784.50 | 55966.54 | 55877.57 | 0.79 | 0.87 | 0.90 | 0.08 | 0.39 | 1086.52 | 0.01 | -1035.48 |
| 1.00 | 5.00 | -27796.95 | 55661.90 | 55882.95 | 55774.91 | 0.78 | 0.52 | 0.91 | 0.04 | 0.38 | 568.55 | 0.01 | -83.59 |
| 1.00 | 6.00 | -27457.50 | 54994.99 | 55255.05 | 55127.95 | 0.78 | 0.80 | 0.87 | 0.02 | 0.31 | 245.61 | 0.01 | -627.89 |
| 1.00 | 7.00 | -27397.94 | 54887.89 | 55186.95 | 55040.78 | 0.78 | 0.53 | 0.88 | 0.02 | 0.31 | 183.34 | 0.01 | -68.10 |
| General Anxiety | | | | | | | | | | | | | |

| | | | | | | | | | | | | | |
|---|---|---|---|---|---|---|---|---|---|---|---|---|---|
| Model | Classes | LogLik | AIC | BIC | SABIC | Entropy | prob_min | prob_max | n_min | n_max | BLRT_val | BLRT_p | delta_BIC |
| 1.00 | 2.00 | -32081.53 | 64195.06 | 64300.15 | 64249.31 | 0.88 | 0.95 | 0.98 | 0.33 | 0.67 | 11149.16 | 0.01 | 0.00 |
| 1.00 | 3.00 | -30216.58 | 60477.16 | 60621.67 | 60551.76 | 0.85 | 0.91 | 0.95 | 0.13 | 0.47 | 3730.17 | 0.01 | -3678.48 |
| 1.00 | 4.00 | -29334.99 | 58725.97 | 58909.89 | 58820.92 | 0.82 | 0.85 | 0.93 | 0.09 | 0.46 | 1760.82 | 0.01 | -1711.78 |
| 1.00 | 5.00 | -28818.15 | 57704.30 | 57927.63 | 57819.59 | 0.85 | 0.87 | 0.92 | 0.03 | 0.38 | 1037.98 | 0.01 | -982.26 |
| 1.00 | 6.00 | -28498.78 | 57077.56 | 57340.30 | 57213.20 | 0.82 | 0.85 | 0.91 | 0.02 | 0.35 | 642.34 | 0.01 | -587.33 |
| 1.00 | 7.00 | -28432.51 | 56957.02 | 57259.16 | 57112.99 | 0.79 | 0.63 | 0.91 | 0.02 | 0.36 | 127.39 | 0.01 | -81.14 |
| Self Harm | | | | | | | | | | | | | |
| Model | Classes | LogLik | AIC | BIC | SABIC | Entropy | prob_min | prob_max | n_min | n_max | BLRT_val | BLRT_p | delta_BIC |
| 1.00 | 2.00 | -34102.42 | 68236.84 | 68341.92 | 68291.07 | 0.99 | 0.97 | 1.00 | 0.02 | 0.98 | 5701.83 | 0.01 | 0.00 |
| 1.00 | 3.00 | -33432.73 | 66909.45 | 67053.93 | 66984.02 | 0.91 | 0.75 | 0.99 | 0.02 | 0.90 | 1339.99 | 0.01 | -1287.99 |
| 1.00 | 4.00 | -31839.66 | 63735.31 | 63919.19 | 63830.21 | 0.92 | 0.83 | 1.00 | 0.01 | 0.82 | 3185.82 | 0.01 | -3134.74 |
| 1.00 | 5.00 | -31839.00 | 63745.99 | 63969.28 | 63861.23 | 0.66 | 0.00 | 1.00 | 0.00 | 0.82 | 0.38 | 0.01 | 50.09 |
| 1.00 | 6.00 | -31839.18 | 63758.35 | 64021.04 | 63893.93 | 0.47 | 0.00 | 1.00 | 0.00 | 0.55 | 0.58 | 0.01 | 51.76 |
| 1.00 | 7.00 | -31267.50 | 62627.01 | 62929.10 | 62782.92 | 0.52 | 0.00 | 1.00 | 0.00 | 0.75 | 2.68 | 0.01 | -1091.94 |

Table S5. Phenotypic correlations between T1 and T5

| **Emotional problems** | | 95% CI | | **Conduct problems** | | 95% CI | | **Hyperactivity** | | 95% CI | | **Peer problems** | | 95% CI | |
|---|---|---|---|---|---|---|---|---|---|---|---|---|---|---|---|
| Time | r | lower | upper | Time | r | lower | upper | Time | r | lower | upper | Time | r | lower | upper |
| T1-T2 | 0.56 | 0.53 | 0.58 | T1-T2 | 0.31 | 0.28 | 0.35 | T1-T2 | 0.44 | 0.4 | 0.47 | T1-T2 | 0.57 | 0.54 | 0.6 |
| T1-T3 | 0.52 | 0.49 | 0.56 | T1-T3 | 0.36 | 0.32 | 0.4 | T1-T3 | 0.45 | 0.41 | 0.48 | T1-T3 | 0.54 | 0.51 | 0.58 |
| T1-T4 | 0.53 | 0.49 | 0.56 | T1-T4 | 0.3 | 0.26 | 0.34 | T1-T4 | 0.46 | 0.42 | 0.5 | T1-T4 | 0.53 | 0.49 | 0.56 |
| T1-T5 | 0.52 | 0.49 | 0.55 | T1-T5 | 0.34 | 0.3 | 0.38 | T1-T5 | 0.45 | 0.41 | 0.48 | T1-T5 | 0.53 | 0.5 | 0.57 |
| T2-T3 | 0.72 | 0.69 | 0.74 | T2-T3 | 0.55 | 0.52 | 0.58 | T2-T3 | 0.64 | 0.61 | 0.67 | T2-T3 | 0.72 | 0.7 | 0.75 |
| T2-T4 | 0.68 | 0.66 | 0.71 | T2-T4 | 0.46 | 0.42 | 0.5 | T2-T4 | 0.6 | 0.57 | 0.64 | T2-T4 | 0.66 | 0.63 | 0.69 |
| T2-T5 | 0.62 | 0.59 | 0.65 | T2-T5 | 0.41 | 0.37 | 0.45 | T2-T5 | 0.56 | 0.52 | 0.59 | T2-T5 | 0.65 | 0.62 | 0.68 |
| T3-T4 | 0.69 | 0.66 | 0.72 | T3-T4 | 0.51 | 0.47 | 0.55 | T3-T4 | 0.65 | 0.61 | 0.68 | T3-T4 | 0.68 | 0.66 | 0.71 |
| T3-T5 | 0.67 | 0.63 | 0.69 | T3-T5 | 0.45 | 0.41 | 0.49 | T3-T5 | 0.6 | 0.56 | 0.63 | T3-T5 | 0.69 | 0.66 | 0.71 |
| T4-T5 | 0.7 | 0.67 | 0.72 | T4-T5 | 0.48 | 0.43 | 0.52 | T4-T5 | 0.64 | 0.61 | 0.67 | T4-T5 | 0.68 | 0.65 | 0.71 |

| **Prosocial Behaviour** | | 95% CI | | **Behaviour problems** | | 95% CI | | **Depression** | | 95% CI | | **General Anxiety** | | 95% CI | |
|---|---|---|---|---|---|---|---|---|---|---|---|---|---|---|---|
| Time | r | lower | upper | Time | r | lower | upper | Time | r | lower | upper | Time | r | lower | upper |
| T1-T2 | 0.47 | 0.43 | 0.5 | T1-T2 | 0.6 | 0.58 | 0.63 | T1-T2 | 0.56 | 0.53 | 0.58 | T1-T2 | 0.57 | 0.54 | 0.6 |
| T1-T3 | 0.47 | 0.44 | 0.51 | T1-T3 | 0.59 | 0.56 | 0.62 | T1-T3 | 0.54 | 0.5 | 0.57 | T1-T3 | 0.56 | 0.53 | 0.59 |
| T1-T4 | 0.42 | 0.38 | 0.46 | T1-T4 | 0.6 | 0.57 | 0.63 | T1-T4 | 0.56 | 0.53 | 0.6 | T1-T4 | 0.54 | 0.51 | 0.58 |
| T1-T5 | 0.41 | 0.37 | 0.45 | T1-T5 | 0.58 | 0.55 | 0.61 | T1-T5 | 0.55 | 0.51 | 0.58 | T1-T5 | 0.55 | 0.52 | 0.58 |
| T2-T3 | 0.64 | 0.61 | 0.66 | T2-T3 | 0.77 | 0.75 | 0.79 | T2-T3 | 0.73 | 0.7 | 0.75 | T2-T3 | 0.75 | 0.73 | 0.77 |
| T2-T4 | 0.59 | 0.55 | 0.62 | T2-T4 | 0.73 | 0.7 | 0.75 | T2-T4 | 0.68 | 0.65 | 0.71 | T2-T4 | 0.71 | 0.69 | 0.74 |
| T2-T5 | 0.56 | 0.52 | 0.59 | T2-T5 | 0.67 | 0.64 | 0.7 | T2-T5 | 0.63 | 0.6 | 0.66 | T2-T5 | 0.7 | 0.67 | 0.72 |
| T3-T4 | 0.62 | 0.59 | 0.65 | T3-T4 | 0.75 | 0.72 | 0.77 | T3-T4 | 0.71 | 0.68 | 0.74 | T3-T4 | 0.71 | 0.69 | 0.74 |
| T3-T5 | 0.59 | 0.56 | 0.63 | T3-T5 | 0.71 | 0.68 | 0.74 | T3-T5 | 0.66 | 0.63 | 0.69 | T3-T5 | 0.72 | 0.69 | 0.74 |
| T4-T5 | 0.6 | 0.57 | 0.64 | T4-T5 | 0.74 | 0.72 | 0.77 | T4-T5 | 0.71 | 0.68 | 0.74 | T4-T5 | 0.76 | 0.74 | 0.78 |

Table S6. Univariate twin model-fitting results

| Emotional problems | h2 | 95% CI | | c2 | 95% CI | | e2 | 95% CI | |
|---|---|---|---|---|---|---|---|---|---|
| Time | | lower | upper | | lower | upper | | lower | upper |
| T1 | 0.31 | 0.19 | 0.38 | 0.04 | 0.00 | 0.12 | 0.66 | 0.62 | 0.70 |
| T2 | 0.40 | 0.27 | 0.46 | 0.00 | 0.00 | 0.10 | 0.60 | 0.54 | 0.65 |
| T3 | 0.23 | 0.02 | 0.40 | 0.10 | 0.00 | 0.26 | 0.67 | 0.60 | 0.74 |
| T4 | 0.39 | 0.30 | 0.45 | 0.00 | 0.00 | 0.05 | 0.61 | 0.55 | 0.68 |
| T5 | 0.40 | 0.20 | 0.47 | 0.01 | 0.00 | 0.17 | 0.59 | 0.53 | 0.66 |

| Conduct Problems | h2 | 95% CI | | c2 | 95% CI | | e2 | 95% CI | |
|---|---|---|---|---|---|---|---|---|---|
| Time | | lower | upper | | lower | upper | | lower | upper |
| T1 | 0.27 | 0.16 | 0.31 | 0.00 | 0.00 | 0.08 | 0.73 | 0.69 | 0.78 |
| T2 | 0.00 | 0.00 | 0.17 | 0.19 | 0.06 | 0.24 | 0.81 | 0.74 | 0.86 |
| T3 | 0.23 | 0.00 | 0.34 | 0.04 | 0.00 | 0.22 | 0.73 | 0.66 | 0.81 |
| T4 | 0.24 | 0.02 | 0.32 | 0.00 | 0.00 | 0.17 | 0.76 | 0.68 | 0.85 |
| T5 | 0.19 | 0.00 | 0.28 | 0.01 | 0.00 | 0.19 | 0.80 | 0.72 | 0.89 |

| Hyperactivity | h2 | 95% CI | | c2 | 95% CI | | e2 | 95% CI | |
|---|---|---|---|---|---|---|---|---|---|
| Time | | lower | upper | | lower | upper | | lower | upper |
| T1 | 0.34 | 0.25 | 0.38 | 0.00 | 0.00 | 0.07 | 0.66 | 0.62 | 0.70 |
| T2 | 0.31 | 0.23 | 0.37 | 0.00 | 0.00 | 0.05 | 0.69 | 0.63 | 0.75 |
| T3 | 0.29 | 0.08 | 0.39 | 0.03 | 0.00 | 0.20 | 0.68 | 0.61 | 0.76 |
| T4 | 0.35 | 0.15 | 0.41 | 0.00 | 0.00 | 0.16 | 0.65 | 0.59 | 0.73 |
| T5 | 0.32 | 0.16 | 0.39 | 0.00 | 0.00 | 0.12 | 0.68 | 0.61 | 0.75 |

| Peer problems | h2 | 95% CI | | c2 | 95% CI | | e2 | 95% CI | |
|---|---|---|---|---|---|---|---|---|---|
| Time | | lower | upper | | lower | upper | | lower | upper |
| T1 | 0.39 | 0.29 | 0.43 | 0.00 | 0.00 | 0.00 | 0.61 | 0.57 | 0.65 |
| T2 | 0.44 | 0.26 | 0.50 | 0.01 | 0.00 | 0.15 | 0.55 | 0.50 | 0.61 |
| T3 | 0.25 | 0.06 | 0.44 | 0.15 | 0.00 | 0.30 | 0.60 | 0.54 | 0.67 |
| T4 | 0.35 | 0.15 | 0.47 | 0.06 | 0.00 | 0.22 | 0.59 | 0.53 | 0.66 |

| | | | | | | | | | |
|---|---|---|---|---|---|---|---|---|---|
| T5 | 0.44 | 0.25 | 0.50 | 0.00 | 0.00 | 0.15 | 0.56 | 0.50 | 0.63 |

| Prosocial Behaviour | h2 | 95% CI | | c2 | 95% CI | | e2 | 95% CI | |
|---|---|---|---|---|---|---|---|---|---|
| Time | | lower | upper | | lower | upper | | lower | upper |
| T1 | 0.27 | 0.21 | 0.31 | 0.00 | 0.00 | 0.04 | 0.73 | 0.69 | 0.78 |
| T2 | 0.36 | 0.24 | 0.42 | 0.00 | 0.00 | 0.09 | 0.64 | 0.58 | 0.70 |
| T3 | 0.35 | 0.20 | 0.41 | 0.00 | 0.00 | 0.12 | 0.65 | 0.59 | 0.72 |
| T4 | 0.30 | 0.12 | 0.37 | 0.00 | 0.00 | 0.14 | 0.70 | 0.63 | 0.78 |
| T5 | 0.30 | 0.17 | 0.37 | 0.00 | 0.00 | 0.00 | 0.70 | 0.63 | 0.77 |

| Behavioural Problems | h2 | 95% CI | | c2 | 95% CI | | e2 | 95% CI | |
|---|---|---|---|---|---|---|---|---|---|
| Time | | lower | upper | | lower | upper | | lower | upper |
| T1 | 0.37 | 0.26 | 0.47 | 0.07 | 0.00 | 0.15 | 0.57 | 0.53 | 0.61 |
| T2 | 0.48 | 0.31 | 0.53 | 0.00 | 0.00 | 0.14 | 0.52 | 0.47 | 0.58 |
| T3 | 0.25 | 0.06 | 0.45 | 0.16 | 0.00 | 0.32 | 0.59 | 0.52 | 0.66 |
| T4 | 0.47 | 0.36 | 0.53 | 0.00 | 0.00 | 0.00 | 0.53 | 0.47 | 0.59 |
| T5 | 0.40 | 0.21 | 0.49 | 0.03 | 0.00 | 0.18 | 0.57 | 0.51 | 0.64 |

| Depression | h2 | 95% CI | | c2 | 95% CI | | e2 | 95% CI | |
|---|---|---|---|---|---|---|---|---|---|
| Time | | lower | upper | | lower | upper | | lower | upper |
| T1 | 0.31 | 0.19 | 0.36 | 0.01 | 0.00 | 0.10 | 0.68 | 0.64 | 0.73 |
| T2 | 0.37 | 0.23 | 0.43 | 0.00 | 0.00 | 0.11 | 0.63 | 0.57 | 0.69 |
| T3 | 0.35 | 0.19 | 0.41 | 0.00 | 0.00 | 0.13 | 0.65 | 0.59 | 0.72 |
| T4 | 0.38 | 0.25 | 0.45 | 0.00 | 0.00 | 0.10 | 0.62 | 0.55 | 0.68 |
| T5 | 0.33 | 0.12 | 0.44 | 0.05 | 0.00 | 0.21 | 0.62 | 0.56 | 0.70 |

| General Anxiety | h2 | 95% CI | | c2 | 95% CI | | e2 | 95% CI | |
|---|---|---|---|---|---|---|---|---|---|
| Time | | lower | upper | | lower | upper | | lower | upper |
| T1 | 0.33 | 0.28 | 0.38 | 0.00 | 0.00 | 0.00 | 0.67 | 0.62 | 0.71 |
| T2 | 0.43 | 0.33 | 0.49 | 0.00 | 0.00 | 0.00 | 0.57 | 0.51 | 0.62 |

| | | | | | | | | | |
|---|---|---|---|---|---|---|---|---|---|
| T3 | 0.25 | 0.03 | 0.37 | 0.05 | 0.00 | 0.23 | 0.70 | 0.63 | 0.77 |
| T4 | 0.39 | 0.24 | 0.45 | 0.00 | 0.00 | 0.11 | 0.61 | 0.55 | 0.68 |
| T5 | 0.44 | 0.24 | 0.51 | 0.01 | 0.00 | 0.17 | 0.55 | 0.49 | 0.62 |

Table S7. Genetic (rA), shared environmental (rC) and non-shared environmental correlations between (rE) T1 and T5

| **Emotional problems** | | 95% CI | | **Conduct problems** | | 95% CI | | **Hyperactivity** | | 95% CI | | **Peer problems** | | 95% CI | |
|---|---|---|---|---|---|---|---|---|---|---|---|---|---|---|---|
| Time | rA | lower | upper | Time | rA | lower | upper | Time | rA | lower | upper | Time | rA | lower | upper |
| T1-T2 | 0.86 | 0.75 | 0.97 | T1-T2 | 0.78 | 0.45 | 1.00 | T1-T2 | 0.84 | 0.69 | 1.00 | T1-T2 | 0.94 | 0.87 | 1.00 |
| T1-T3 | 0.95 | 0.83 | 1.00 | T1-T3 | 0.79 | 0.54 | 1.00 | T1-T3 | 0.86 | 0.69 | 1.00 | T1-T3 | 0.92 | 0.83 | 1.00 |
| T1-T4 | 0.96 | 0.88 | 1.00 | T1-T4 | 0.83 | 0.63 | 1.00 | T1-T4 | 0.84 | 0.68 | 1.00 | T1-T4 | 0.95 | 0.87 | 1.00 |
| T1-T5 | 0.88 | 0.75 | 0.99 | T1-T5 | 0.83 | 0.58 | 1.00 | T1-T5 | 0.78 | 0.62 | 1.00 | T1-T5 | 0.92 | 0.82 | 1.00 |
| T2-T3 | 0.97 | 0.84 | 1.00 | T2-T3 | 0.93 | 0.59 | 1.00 | T2-T3 | 1.00 | 0.93 | 1.00 | T2-T3 | 1.00 | 0.93 | 1.00 |
| T2-T4 | 0.97 | 0.89 | 1.00 | T2-T4 | 0.99 | 0.72 | 1.00 | T2-T4 | 0.94 | 0.83 | 1.00 | T2-T4 | 1.00 | 0.93 | 1.00 |
| T2-T5 | 1.00 | 0.87 | 1.00 | T2-T5 | 1.00 | 0.76 | 1.00 | T2-T5 | 0.98 | 0.90 | 1.00 | T2-T5 | 1.00 | 0.94 | 1.00 |
| T3-T4 | 1.00 | 0.92 | 1.00 | T3-T4 | 0.90 | 0.61 | 1.00 | T3-T4 | 0.96 | 0.82 | 1.00 | T3-T4 | 1.00 | 0.91 | 1.00 |
| T3-T5 | 0.98 | 0.85 | 1.00 | T3-T5 | 0.94 | 0.68 | 1.00 | T3-T5 | 0.97 | 0.82 | 1.00 | T3-T5 | 1.00 | 0.96 | 1.00 |
| T4-T5 | 0.98 | 0.92 | 1.00 | T4-T5 | 0.99 | 0.75 | 1.00 | T4-T5 | 0.87 | 0.73 | 1.00 | T4-T5 | 0.99 | 0.89 | 1.00 |
| **Prosocial Behaviour** | | 95% CI | | **Behaviour problems** | | 95% CI | | **Depression** | | 95% CI | | **General Anxiety** | | 95% CI | |
| Time | rA | lower | upper | Time | rA | lower | upper | Time | rA | lower | upper | Time | rA | lower | upper |
| T1-T2 | 0.97 | 0.83 | 1.00 | T1-T2 | 0.84 | 0.76 | 0.95 | T1-T2 | 0.97 | 0.90 | 1.00 | T1-T2 | 0.81 | 0.88 | 1.00 |
| T1-T3 | 1.00 | 0.85 | 1.00 | T1-T3 | 0.89 | 0.80 | 0.99 | T1-T3 | 0.92 | 0.82 | 1.00 | T1-T3 | 0.86 | 0.94 | 1.00 |
| T1-T4 | 0.99 | 0.87 | 1.00 | T1-T4 | 0.92 | 0.84 | 0.99 | T1-T4 | 0.93 | 0.84 | 1.00 | T1-T4 | 0.83 | 0.92 | 1.00 |
| T1-T5 | 0.99 | 0.87 | 1.00 | T1-T5 | 0.79 | 0.69 | 0.91 | T1-T5 | 0.93 | 0.83 | 1.00 | T1-T5 | 0.80 | 0.88 | 1.00 |
| T2-T3 | 0.97 | 0.86 | 1.00 | T2-T3 | 0.99 | 0.95 | 1.00 | T2-T3 | 0.99 | 0.94 | 1.00 | T2-T3 | 0.92 | 0.97 | 1.00 |

| | | | | | | | | | | | | | | | |
|---|---|---|---|---|---|---|---|---|---|---|---|---|---|---|---|
| T2-T4 | 0.98 | 0.83 | 1.00 | T2-T4 | 0.97 | 0.92 | 1.00 | T2-T4 | 0.99 | 0.93 | 1.00 | T2-T4 | 0.90 | 0.96 | 1.00 |
| T2-T5 | 0.94 | 0.78 | 1.00 | T2-T5 | 0.99 | 0.93 | 1.00 | T2-T5 | 0.99 | 0.93 | 1.00 | T2-T5 | 0.95 | 1.00 | 1.00 |
| T3-T4 | 0.98 | 0.88 | 1.00 | T3-T4 | 0.97 | 0.91 | 1.00 | T3-T4 | 0.97 | 0.91 | 1.00 | T3-T4 | 0.92 | 1.00 | 1.00 |
| T3-T5 | 0.99 | 0.88 | 1.00 | T3-T5 | 0.96 | 0.87 | 1.00 | T3-T5 | 0.99 | 0.90 | 1.00 | T3-T5 | 0.89 | 0.98 | 1.00 |
| T4-T5 | 0.98 | 0.90 | 1.00 | T4-T5 | 0.96 | 0.88 | 1.00 | T4-T5 | 1.00 | 0.93 | 1.00 | T4-T5 | 0.91 | 0.96 | 1.00 |
| **Emotional problems** | | 95% CI | | **Conduct problems** | | 95% CI | | **Hyperactivity** | | 95% CI | | **Peer problems** | | 95% CI | |
| Time | rC | lower | upper | Time | rC | lower | upper | Time | rC | lower | upper | Time | rC | lower | upper |
| T1-T2 | 0.95 | -1.00 | 1.00 | T1-T2 | 1.00 | -1.00 | NA | T1-T2 | -0.44 | -1.00 | 1.00 | T1-T2 | 0.99 | -1.00 | 1.00 |
| T1-T3 | 0.94 | -1.00 | 1.00 | T1-T3 | 0.89 | -1.00 | NA | T1-T3 | -0.31 | -1.00 | 1.00 | T1-T3 | 0.96 | -1.00 | 1.00 |
| T1-T4 | 0.94 | -1.00 | 1.00 | T1-T4 | 0.54 | -1.00 | 1.00 | T1-T4 | 0.69 | -1.00 | 1.00 | T1-T4 | -0.63 | -1.00 | 1.00 |
| T1-T5 | 0.79 | -1.00 | 1.00 | T1-T5 | 0.77 | -1.00 | 1.00 | T1-T5 | 0.54 | -1.00 | 1.00 | T1-T5 | 0.98 | -1.00 | 1.00 |
| T2-T3 | 0.81 | -1.00 | 1.00 | T2-T3 | 0.92 | -1.00 | NA | T2-T3 | 0.16 | -1.00 | 1.00 | T2-T3 | 0.89 | -1.00 | 1.00 |
| T2-T4 | 0.78 | -1.00 | 1.00 | T2-T4 | 0.47 | -1.00 | 1.00 | T2-T4 | 0.34 | -1.00 | 1.00 | T2-T4 | -0.75 | -1.00 | 1.00 |
| T2-T5 | 0.57 | -1.00 | 1.00 | T2-T5 | 0.82 | -1.00 | 1.00 | T2-T5 | 0.31 | -1.00 | 1.00 | T2-T5 | 0.94 | -1.00 | 1.00 |
| T3-T4 | 0.97 | -1.00 | 1.00 | T3-T4 | 0.08 | -1.00 | 1.00 | T3-T4 | -0.17 | -1.00 | 1.00 | T3-T4 | -0.38 | -1.00 | 1.00 |
| T3-T5 | 0.81 | -1.00 | 1.00 | T3-T5 | 0.98 | -1.00 | 1.00 | T3-T5 | -0.70 | -1.00 | 1.00 | T3-T5 | 0.99 | 0.27 | 1.00 |
| T4-T5 | 0.92 | -1.00 | 1.00 | T4-T5 | -0.12 | -1.00 | 1.00 | T4-T5 | 0.80 | -1.00 | 1.00 | T4-T5 | -0.48 | -1.00 | 1.00 |
| **Prosocial Behaviour** | | 95% CI | | **Behaviour problems** | | 95% CI | | **Depression** | | 95% CI | | **General Anxiety** | | 95% CI | |
| Time | rC | lower | upper | Time | rC | lower | upper | Time | rC | lower | upper | Time | rC | lower | upper |
| T1-T2 | -0.75 | -1.00 | 1.00 | T1-T2 | 1.00 | -1.00 | 1.00 | T1-T2 | 1.00 | -1.00 | 1.00 | T1-T2 | 1.00 | -1.00 | 1.00 |
| T1-T3 | -0.88 | -1.00 | 1.00 | T1-T3 | 0.92 | 0.47 | 1.00 | T1-T3 | 1.00 | -1.00 | 1.00 | T1-T3 | 1.00 | -1.00 | 1.00 |
| T1-T4 | -0.03 | -1.00 | 1.00 | T1-T4 | 0.97 | -1.00 | 1.00 | T1-T4 | 1.00 | -1.00 | 1.00 | T1-T4 | -0.99 | -1.00 | 1.00 |

| | | | | | | | | | | | | | | | |
|---|---|---|---|---|---|---|---|---|---|---|---|---|---|---|---|
| T1-T5 | -0.17 | -1.00 | 1.00 | T1-T5 | 1.00 | -1.00 | 1.00 | T1-T5 | 1.00 | -1.00 | 1.00 | T1-T5 | -1.00 | -1.00 | 1.00 |
| T2-T3 | 0.34 | -1.00 | 1.00 | T2-T3 | 0.94 | -1.00 | 1.00 | T2-T3 | 1.00 | -1.00 | 1.00 | T2-T3 | 1.00 | -1.00 | 1.00 |
| T2-T4 | -0.64 | -1.00 | 1.00 | T2-T4 | 0.98 | -1.00 | 1.00 | T2-T4 | 1.00 | -1.00 | 1.00 | T2-T4 | -0.99 | -1.00 | 1.00 |
| T2-T5 | -0.52 | -1.00 | 1.00 | T2-T5 | 0.99 | 0.72 | 1.00 | T2-T5 | 1.00 | -1.00 | 1.00 | T2-T5 | -0.99 | -1.00 | 1.00 |
| T3-T4 | 0.51 | -1.00 | 1.00 | T3-T4 | 0.99 | -1.00 | 1.00 | T3-T4 | 1.00 | -1.00 | 1.00 | T3-T4 | -0.99 | -1.00 | 1.00 |
| T3-T5 | 0.63 | -1.00 | 1.00 | T3-T5 | 0.89 | -1.00 | 1.00 | T3-T5 | 1.00 | -1.00 | 1.00 | T3-T5 | -1.00 | -1.00 | 1.00 |
| T4-T5 | 0.99 | -1.00 | 1.00 | T4-T5 | 0.94 | -1.00 | 1.00 | T4-T5 | 1.00 | -1.00 | 1.00 | T4-T5 | 1.00 | -1.00 | 1.00 |
| **Emotional problems** | | 95% CI | | **Conduct problems** | | 95% CI | | **Hyperactivity** | | 95% CI | | **Peer problems** | | 95% CI | |
| Time | rE | lower | upper | Time | rE | lower | upper | Time | rE | lower | upper | Time | rE | lower | upper |
| T1-T2 | 0.34 | 0.29 | 0.39 | T1-T2 | 0.22 | 0.16 | 0.27 | T1-T2 | 0.26 | 0.21 | 0.31 | T1-T2 | 0.30 | 0.25 | 0.35 |
| T1-T3 | 0.30 | 0.24 | 0.35 | T1-T3 | 0.22 | 0.16 | 0.28 | T1-T3 | 0.28 | 0.22 | 0.33 | T1-T3 | 0.25 | 0.20 | 0.31 |
| T1-T4 | 0.32 | 0.27 | 0.36 | T1-T4 | 0.20 | 0.14 | 0.26 | T1-T4 | 0.29 | 0.23 | 0.35 | T1-T4 | 0.26 | 0.21 | 0.32 |
| T1-T5 | 0.32 | 0.26 | 0.37 | T1-T5 | 0.20 | 0.14 | 0.26 | T1-T5 | 0.30 | 0.25 | 0.36 | T1-T5 | 0.25 | 0.20 | 0.31 |
| T2-T3 | 0.57 | 0.53 | 0.62 | T2-T3 | 0.49 | 0.44 | 0.54 | T2-T3 | 0.50 | 0.46 | 0.54 | T2-T3 | 0.48 | 0.44 | 0.52 |
| T2-T4 | 0.48 | 0.43 | 0.53 | T2-T4 | 0.42 | 0.36 | 0.47 | T2-T4 | 0.43 | 0.38 | 0.48 | T2-T4 | 0.39 | 0.34 | 0.44 |
| T2-T5 | 0.40 | 0.34 | 0.45 | T2-T5 | 0.35 | 0.30 | 0.41 | T2-T5 | 0.37 | 0.32 | 0.42 | T2-T5 | 0.36 | 0.31 | 0.41 |
| T3-T4 | 0.54 | 0.49 | 0.58 | T3-T4 | 0.47 | 0.41 | 0.52 | T3-T4 | 0.52 | 0.48 | 0.57 | T3-T4 | 0.49 | 0.44 | 0.53 |
| T3-T5 | 0.48 | 0.43 | 0.53 | T3-T5 | 0.37 | 0.32 | 0.43 | T3-T5 | 0.47 | 0.41 | 0.51 | T3-T5 | 0.39 | 0.34 | 0.43 |
| T4-T5 | 0.52 | 0.48 | 0.57 | T4-T5 | 0.43 | 0.38 | 0.48 | T4-T5 | 0.52 | 0.47 | 0.57 | T4-T5 | 0.49 | 0.45 | 0.54 |
| **Prosocial Behaviour** | | 95% CI | | **Behaviour problems** | | 95% CI | | **Depression** | | 95% CI | | **General Anxiety** | | 95% CI | |
| Time | rE | lower | upper | Time | rE | lower | upper | Time | rE | lower | upper | Time | rE | lower | upper |
| T1-T2 | 0.25 | 0.21 | 0.30 | T1-T2 | 0.38 | 0.33 | 0.42 | T1-T2 | 0.31 | 0.26 | 0.35 | T1-T2 | 0.38 | 0.33 | 0.43 |
| T1-T3 | 0.25 | 0.20 | 0.30 | T1-T3 | 0.35 | 0.30 | 0.40 | T1-T3 | 0.34 | 0.29 | 0.39 | T1-T3 | 0.40 | 0.35 | 0.45 |

| | | | | | | | | | | | | | | | |
|---|---|---|---|---|---|---|---|---|---|---|---|---|---|---|---|
| T1-T4 | 0.22 | 0.17 | 0.27 | T1-T4 | 0.35 | 0.30 | 0.40 | T1-T4 | 0.35 | 0.30 | 0.40 | T1-T4 | 0.38 | 0.33 | 0.44 |
| T1-T5 | 0.21 | 0.16 | 0.26 | T1-T5 | 0.39 | 0.34 | 0.44 | T1-T5 | 0.30 | 0.25 | 0.35 | T1-T5 | 0.39 | 0.33 | 0.44 |
| T2-T3 | 0.47 | 0.42 | 0.51 | T2-T3 | 0.58 | 0.54 | 0.62 | T2-T3 | 0.56 | 0.52 | 0.60 | T2-T3 | 0.62 | 0.58 | 0.65 |
| T2-T4 | 0.42 | 0.37 | 0.47 | T2-T4 | 0.49 | 0.44 | 0.54 | T2-T4 | 0.45 | 0.40 | 0.49 | T2-T4 | 0.52 | 0.47 | 0.57 |
| T2-T5 | 0.40 | 0.34 | 0.45 | T2-T5 | 0.43 | 0.38 | 0.48 | T2-T5 | 0.41 | 0.37 | 0.46 | T2-T5 | 0.46 | 0.41 | 0.50 |
| T3-T4 | 0.46 | 0.41 | 0.51 | T3-T4 | 0.58 | 0.53 | 0.62 | T3-T4 | 0.55 | 0.50 | 0.60 | T3-T4 | 0.60 | 0.56 | 0.64 |
| T3-T5 | 0.40 | 0.35 | 0.45 | T3-T5 | 0.54 | 0.49 | 0.58 | T3-T5 | 0.49 | 0.44 | 0.53 | T3-T5 | 0.57 | 0.53 | 0.62 |
| T4-T5 | 0.43 | 0.38 | 0.48 | T4-T5 | 0.61 | 0.57 | 0.66 | T4-T5 | 0.55 | 0.50 | 0.59 | T4-T5 | 0.62 | 0.58 | 0.66 |

Table S8. Results of multivariate Cholesky analyses presenting standardised squared path estimates

| **Emotional problems** | | 95% CI | | | **Emotional problems** | | 95% CI | | | **Emotional problems** | | 95% CI | |
|---|---|---|---|---|---|---|---|---|---|---|---|---|---|
| | estimate | lower | upper | | | estimate | lower | upper | | | estimate | lower | upper |
| CholACE.sta2[1,1] | 0.29 | 0.20 | 0.37 | | CholACE.stc2[1,1] | 0.05 | 0.00 | 0.12 | | CholACE.ste2[1,1] | 0.66 | 0.61 | 0.71 |
| CholACE.sta2[2,1] | 0.25 | 0.15 | 0.37 | | CholACE.stc2[2,1] | 0.04 | 0.00 | 0.13 | | CholACE.ste2[2,1] | 0.07 | 0.05 | 0.10 |
| CholACE.sta2[3,1] | 0.25 | 0.15 | 0.38 | | CholACE.stc2[3,1] | 0.07 | 0.00 | 0.17 | | CholACE.ste2[3,1] | 0.06 | 0.04 | 0.08 |
| CholACE.sta2[4,1] | 0.33 | 0.23 | 0.41 | | CholACE.stc2[4,1] | 0.02 | 0.00 | 0.08 | | CholACE.ste2[4,1] | 0.06 | 0.04 | 0.09 |
| CholACE.sta2[5,1] | 0.27 | 0.15 | 0.40 | | CholACE.stc2[5,1] | 0.03 | 0.00 | 0.13 | | CholACE.ste2[5,1] | 0.06 | 0.04 | 0.08 |
| CholACE.sta2[2,2] | 0.09 | 0.02 | 0.15 | | CholACE.stc2[2,2] | 0.00 | 0.00 | 0.05 | | CholACE.ste2[2,2] | 0.53 | 0.48 | 0.59 |
| CholACE.sta2[3,2] | 0.03 | 0.00 | 0.08 | | CholACE.stc2[3,2] | 0.01 | 0.00 | 0.05 | | CholACE.ste2[3,2] | 0.16 | 0.13 | 0.20 |
| CholACE.sta2[4,2] | 0.03 | 0.00 | 0.08 | | CholACE.stc2[4,2] | 0.00 | 0.00 | 0.03 | | CholACE.ste2[4,2] | 0.10 | 0.07 | 0.13 |
| CholACE.sta2[5,2] | 0.08 | 0.00 | 0.15 | | CholACE.stc2[5,2] | 0.01 | 0.00 | 0.08 | | CholACE.ste2[5,2] | 0.06 | 0.04 | 0.08 |
| CholACE.sta2[3,3] | 0.00 | 0.00 | 0.06 | | CholACE.stc2[3,3] | 0.00 | 0.00 | 0.05 | | CholACE.ste2[3,3] | 0.42 | 0.37 | 0.46 |
| CholACE.sta2[4,3] | 0.00 | 0.00 | 0.04 | | CholACE.stc2[4,3] | 0.00 | 0.00 | 0.03 | | CholACE.ste2[4,3] | 0.06 | 0.04 | 0.08 |
| CholACE.sta2[5,3] | 0.00 | 0.00 | 0.08 | | CholACE.stc2[5,3] | 0.00 | 0.00 | 0.07 | | CholACE.ste2[5,3] | 0.05 | 0.03 | 0.07 |
| CholACE.sta2[4,4] | 0.00 | 0.00 | 0.03 | | CholACE.stc2[4,4] | 0.00 | 0.00 | 0.02 | | CholACE.ste2[4,4] | 0.40 | 0.36 | 0.44 |
| CholACE.sta2[5,4] | 0.00 | 0.00 | 0.07 | | CholACE.stc2[5,4] | 0.00 | 0.00 | 0.06 | | CholACE.ste2[5,4] | 0.04 | 0.03 | 0.06 |
| CholACE.sta2[5,5] | 0.00 | 0.00 | 0.05 | | CholACE.stc2[5,5] | 0.00 | 0.00 | 0.04 | | CholACE.ste2[5,5] | 0.39 | 0.35 | 0.44 |

| Conduct problems | | 95% CI | | | Conduct problems | | 95% CI | | | Conduct problems | | 95% CI | |
|---|---|---|---|---|---|---|---|---|---|---|---|---|---|
| | estimate | lower | upper | | | estimate | lower | upper | | | estimate | lower | upper |
| CholACE.sta2[1,1] | 0.26 | 0.15 | 0.31 | | CholACE.stc2[1,1] | 0.01 | 0.00 | 0.10 | | CholACE.ste2[1,1] | 0.72 | 0.67 | 0.77 |
| CholACE.sta2[2,1] | 0.08 | 0.02 | 0.21 | | CholACE.stc2[2,1] | 0.08 | 0.00 | 0.17 | | CholACE.ste2[2,1] | 0.04 | 0.02 | 0.06 |
| CholACE.sta2[3,1] | 0.13 | 0.03 | 0.25 | | CholACE.stc2[3,1] | 0.02 | 0.00 | 0.15 | | CholACE.ste2[3,1] | 0.04 | 0.02 | 0.06 |
| CholACE.sta2[4,1] | 0.16 | 0.06 | 0.28 | | CholACE.stc2[4,1] | 0.00 | 0.00 | 0.11 | | CholACE.ste2[4,1] | 0.03 | 0.01 | 0.05 |
| CholACE.sta2[5,1] | 0.12 | 0.04 | 0.24 | | CholACE.stc2[5,1] | 0.02 | 0.00 | 0.13 | | CholACE.ste2[5,1] | 0.03 | 0.02 | 0.05 |
| CholACE.sta2[2,2] | 0.05 | 0.00 | 0.14 | | CholACE.stc2[2,2] | 0.00 | 0.00 | 0.13 | | CholACE.ste2[2,2] | 0.72 | 0.66 | 0.79 |
| CholACE.sta2[3,2] | 0.05 | 0.00 | 0.14 | | CholACE.stc2[3,2] | 0.00 | 0.00 | 0.11 | | CholACE.ste2[3,2] | 0.15 | 0.12 | 0.20 |
| CholACE.sta2[4,2] | 0.07 | 0.00 | 0.14 | | CholACE.stc2[4,2] | 0.00 | 0.00 | 0.09 | | CholACE.ste2[4,2] | 0.11 | 0.08 | 0.15 |
| CholACE.sta2[5,2] | 0.05 | 0.00 | 0.13 | | CholACE.stc2[5,2] | 0.01 | 0.00 | 0.10 | | CholACE.ste2[5,2] | 0.08 | 0.05 | 0.11 |
| CholACE.sta2[3,3] | 0.02 | 0.00 | 0.09 | | CholACE.stc2[3,3] | 0.00 | 0.00 | 0.07 | | CholACE.ste2[3,3] | 0.56 | 0.50 | 0.62 |
| CholACE.sta2[4,3] | 0.00 | 0.00 | 0.08 | | CholACE.stc2[4,3] | 0.00 | 0.00 | 0.07 | | CholACE.ste2[4,3] | 0.06 | 0.04 | 0.09 |
| CholACE.sta2[5,3] | 0.00 | 0.00 | 0.05 | | CholACE.stc2[5,3] | 0.00 | 0.00 | 0.06 | | CholACE.ste2[5,3] | 0.03 | 0.02 | 0.06 |
| CholACE.sta2[4,4] | 0.00 | 0.00 | 0.08 | | CholACE.stc2[4,4] | 0.00 | 0.00 | 0.06 | | CholACE.ste2[4,4] | 0.55 | 0.48 | 0.61 |
| CholACE.sta2[5,4] | 0.00 | 0.00 | 0.05 | | CholACE.stc2[5,4] | 0.00 | 0.00 | 0.05 | | CholACE.ste2[5,4] | 0.05 | 0.03 | 0.07 |
| CholACE.sta2[5,5] | 0.00 | 0.00 | 0.00 | | CholACE.stc2[5,5] | 0.00 | 0.00 | 0.05 | | CholACE.ste2[5,5] | 0.58 | 0.52 | 0.64 |
| **Hyperactivity** | | 95% CI | | | **Hyperactivity** | | 95% CI | | | **Hyperactivity** | | 95% CI | |
| | estimate | lower | upper | | | estimate | lower | upper | | | estimate | lower | upper |
| CholACE.sta2[1,1] | 0.22 | 0.31 | 0.37 | | CholACE.stc2[1,1] | 0.03 | 0.00 | 0.10 | | CholACE.ste2[1,1] | 0.66 | 0.61 | 0.71 |
| CholACE.sta2[2,1] | 0.13 | 0.22 | 0.33 | | CholACE.stc2[2,1] | 0.00 | 0.00 | 0.05 | | CholACE.ste2[2,1] | 0.05 | 0.03 | 0.07 |
| CholACE.sta2[3,1] | 0.13 | 0.23 | 0.34 | | CholACE.stc2[3,1] | 0.00 | 0.00 | 0.08 | | CholACE.ste2[3,1] | 0.05 | 0.03 | 0.08 |
| CholACE.sta2[4,1] | 0.11 | 0.21 | 0.36 | | CholACE.stc2[4,1] | 0.02 | 0.00 | 0.14 | | CholACE.ste2[4,1] | 0.06 | 0.03 | 0.08 |
| CholACE.sta2[5,1] | 0.09 | 0.17 | 0.30 | | CholACE.stc2[5,1] | 0.01 | 0.00 | 0.09 | | CholACE.ste2[5,1] | 0.06 | 0.04 | 0.09 |
| CholACE.sta2[2,2] | 0.00 | 0.09 | 0.17 | | CholACE.stc2[2,2] | 0.00 | 0.00 | 0.06 | | CholACE.ste2[2,2] | 0.64 | 0.58 | 0.70 |
| CholACE.sta2[3,2] | 0.00 | 0.08 | 0.17 | | CholACE.stc2[3,2] | 0.00 | 0.00 | 0.08 | | CholACE.ste2[3,2] | 0.13 | 0.11 | 0.17 |
| CholACE.sta2[4,2] | 0.00 | 0.06 | 0.16 | | CholACE.stc2[4,2] | 0.02 | 0.00 | 0.12 | | CholACE.ste2[4,2] | 0.09 | 0.06 | 0.12 |
| CholACE.sta2[5,2] | 0.00 | 0.10 | 0.18 | | CholACE.stc2[5,2] | 0.01 | 0.00 | 0.09 | | CholACE.ste2[5,2] | 0.06 | 0.04 | 0.09 |
| CholACE.sta2[3,3] | 0.00 | 0.00 | 0.04 | | CholACE.stc2[3,3] | 0.01 | 0.00 | 0.04 | | CholACE.ste2[3,3] | 0.49 | 0.45 | 0.54 |

| | | | | | | | | | | | | | |
|---|---|---|---|---|---|---|---|---|---|---|---|---|---|
| CholACE.sta2[4,3] | 0.00 | 0.03 | 0.09 | | CholACE.stc2[4,3] | 0.00 | 0.00 | 0.07 | | CholACE.ste2[4,3] | 0.07 | 0.05 | 0.09 |
| CholACE.sta2[5,3] | 0.00 | 0.01 | 0.06 | | CholACE.stc2[5,3] | 0.01 | 0.00 | 0.05 | | CholACE.ste2[5,3] | 0.06 | 0.04 | 0.08 |
| CholACE.sta2[4,4] | 0.00 | 0.00 | 0.09 | | CholACE.stc2[4,4] | 0.00 | 0.00 | 0.07 | | CholACE.ste2[4,4] | 0.44 | 0.39 | 0.49 |
| CholACE.sta2[5,4] | 0.00 | 0.00 | 0.06 | | CholACE.stc2[5,4] | 0.00 | 0.00 | 0.04 | | CholACE.ste2[5,4] | 0.05 | 0.03 | 0.07 |
| CholACE.sta2[5,5] | 0.00 | 0.00 | 0.06 | | CholACE.stc2[5,5] | 0.00 | 0.00 | 0.04 | | CholACE.ste2[5,5] | 0.46 | 0.41 | 0.52 |
| **Peer problems** | | 95% CI | | | **Peer problems** | | 95% CI | | | **Peer problems** | | 95% CI | |
| | estimate | lower | upper | | | estimate | lower | upper | | | estimate | lower | upper |
| CholACE.sta2[1,1] | 0.37 | 0.28 | 0.42 | | CholACE.stc2[1,1] | 0.02 | 0.00 | 0.09 | | CholACE.ste2[1,1] | 0.61 | 0.57 | 0.66 |
| CholACE.sta2[2,1] | 0.36 | 0.26 | 0.43 | | CholACE.stc2[2,1] | 0.01 | 0.00 | 0.10 | | CholACE.ste2[2,1] | 0.05 | 0.04 | 0.07 |
| CholACE.sta2[3,1] | 0.31 | 0.21 | 0.41 | | CholACE.stc2[3,1] | 0.06 | 0.00 | 0.15 | | CholACE.ste2[3,1] | 0.04 | 0.02 | 0.06 |
| CholACE.sta2[4,1] | 0.38 | 0.27 | 0.46 | | CholACE.stc2[4,1] | 0.00 | 0.00 | 0.07 | | CholACE.ste2[4,1] | 0.04 | 0.02 | 0.06 |
| CholACE.sta2[5,1] | 0.28 | 0.19 | 0.38 | | CholACE.stc2[5,1] | 0.05 | 0.00 | 0.13 | | CholACE.ste2[5,1] | 0.04 | 0.02 | 0.06 |
| CholACE.sta2[2,2] | 0.05 | 0.00 | 0.10 | | CholACE.stc2[2,2] | 0.00 | 0.00 | 0.04 | | CholACE.ste2[2,2] | 0.53 | 0.48 | 0.58 |
| CholACE.sta2[3,2] | 0.05 | 0.00 | 0.12 | | CholACE.stc2[3,2] | 0.01 | 0.00 | 0.07 | | CholACE.ste2[3,2] | 0.10 | 0.08 | 0.13 |
| CholACE.sta2[4,2] | 0.04 | 0.00 | 0.11 | | CholACE.stc2[4,2] | 0.00 | 0.00 | 0.06 | | CholACE.ste2[4,2] | 0.06 | 0.04 | 0.09 |
| CholACE.sta2[5,2] | 0.05 | 0.00 | 0.12 | | CholACE.stc2[5,2] | 0.00 | 0.00 | 0.06 | | CholACE.ste2[5,2] | 0.05 | 0.04 | 0.08 |
| CholACE.sta2[3,3] | 0.00 | 0.00 | 0.05 | | CholACE.stc2[3,3] | 0.00 | 0.00 | 0.06 | | CholACE.ste2[3,3] | 0.45 | 0.40 | 0.49 |
| CholACE.sta2[4,3] | 0.00 | 0.00 | 0.06 | | CholACE.stc2[4,3] | 0.00 | 0.00 | 0.05 | | CholACE.ste2[4,3] | 0.06 | 0.04 | 0.08 |
| CholACE.sta2[5,3] | 0.00 | 0.00 | 0.04 | | CholACE.stc2[5,3] | 0.00 | 0.00 | 0.04 | | CholACE.ste2[5,3] | 0.03 | 0.02 | 0.05 |
| CholACE.sta2[4,4] | 0.00 | 0.00 | 0.05 | | CholACE.stc2[4,4] | 0.00 | 0.00 | 0.04 | | CholACE.ste2[4,4] | 0.41 | 0.37 | 0.46 |
| CholACE.sta2[5,4] | 0.00 | 0.00 | 0.02 | | CholACE.stc2[5,4] | 0.00 | 0.00 | 0.01 | | CholACE.ste2[5,4] | 0.06 | 0.04 | 0.07 |
| CholACE.sta2[5,5] | 0.00 | 0.00 | 0.02 | | CholACE.stc2[5,5] | 0.00 | 0.00 | 0.01 | | CholACE.ste2[5,5] | 0.43 | 0.39 | 0.47 |
| **Prosocial Behaviour** | | 95% CI | | | **Prosocial Behaviour** | | 95% CI | | | **Prosocial Behaviour** | | 95% CI | |
| | estimate | lower | upper | | | estimate | lower | upper | | | estimate | lower | upper |
| CholACE.sta2[1,1] | 0.26 | 0.22 | 0.30 | | CholACE.stc2[1,1] | 0.00 | 0.00 | 0.03 | | CholACE.ste2[1,1] | 0.73 | 0.68 | 0.78 |
| CholACE.sta2[2,1] | 0.31 | 0.22 | 0.38 | | CholACE.stc2[2,1] | 0.01 | 0.00 | 0.08 | | CholACE.ste2[2,1] | 0.04 | 0.03 | 0.06 |
| CholACE.sta2[3,1] | 0.32 | 0.23 | 0.39 | | CholACE.stc2[3,1] | 0.01 | 0.00 | 0.08 | | CholACE.ste2[3,1] | 0.04 | 0.03 | 0.06 |
| CholACE.sta2[4,1] | 0.31 | 0.23 | 0.36 | | CholACE.stc2[4,1] | 0.00 | 0.00 | 0.06 | | CholACE.ste2[4,1] | 0.03 | 0.02 | 0.05 |

| | | | | | | | | | | | | | |
|---|---|---|---|---|---|---|---|---|---|---|---|---|---|
| CholACE.sta2[5,1] | 0.31 | 0.23 | 0.37 | | CholACE.stc2[5,1] | 0.00 | 0.00 | 0.06 | | CholACE.ste2[5,1] | 0.03 | 0.02 | 0.05 |
| CholACE.sta2[2,2] | 0.02 | 0.00 | 0.11 | | CholACE.stc2[2,2] | 0.01 | 0.00 | 0.08 | | CholACE.ste2[2,2] | 0.62 | 0.56 | 0.68 |
| CholACE.sta2[3,2] | 0.00 | 0.00 | 0.10 | | CholACE.stc2[3,2] | 0.00 | 0.00 | 0.07 | | CholACE.ste2[3,2] | 0.11 | 0.09 | 0.15 |
| CholACE.sta2[4,2] | 0.00 | 0.00 | 0.08 | | CholACE.stc2[4,2] | 0.01 | 0.00 | 0.06 | | CholACE.ste2[4,2] | 0.10 | 0.07 | 0.13 |
| CholACE.sta2[5,2] | 0.00 | 0.00 | 0.08 | | CholACE.stc2[5,2] | 0.01 | 0.00 | 0.05 | | CholACE.ste2[5,2] | 0.09 | 0.06 | 0.12 |
| CholACE.sta2[3,3] | 0.00 | 0.00 | 0.07 | | CholACE.stc2[3,3] | 0.00 | 0.00 | 0.05 | | CholACE.ste2[3,3] | 0.50 | 0.45 | 0.55 |
| CholACE.sta2[4,3] | 0.00 | 0.00 | 0.07 | | CholACE.stc2[4,3] | 0.00 | 0.00 | 0.05 | | CholACE.ste2[4,3] | 0.05 | 0.04 | 0.08 |
| CholACE.sta2[5,3] | 0.00 | 0.00 | 0.08 | | CholACE.stc2[5,3] | 0.00 | 0.00 | 0.05 | | CholACE.ste2[5,3] | 0.04 | 0.02 | 0.06 |
| CholACE.sta2[4,4] | 0.00 | 0.00 | 0.05 | | CholACE.stc2[4,4] | 0.00 | 0.00 | 0.04 | | CholACE.ste2[4,4] | 0.50 | 0.45 | 0.55 |
| CholACE.sta2[5,4] | 0.00 | 0.00 | 0.05 | | CholACE.stc2[5,4] | 0.00 | 0.00 | 0.03 | | CholACE.ste2[5,4] | 0.03 | 0.02 | 0.05 |
| CholACE.sta2[5,5] | 0.00 | 0.00 | 0.04 | | CholACE.stc2[5,5] | 0.00 | 0.00 | 0.02 | | CholACE.ste2[5,5] | 0.49 | 0.44 | 0.54 |
| **Behaviour problems** | | 95% CI | | | **Behaviour problems** | | 95% CI | | | **Behaviour problems** | | 95% CI | |
| | estimate | lower | upper | | | estimate | lower | upper | | | estimate | lower | upper |
| CholACE.sta2[1,1] | 0.36 | 0.26 | 0.45 | | CholACE.stc2[1,1] | 0.08 | 0.01 | 0.15 | | CholACE.ste2[1,1] | 0.56 | 0.52 | 0.61 |
| CholACE.sta2[2,1] | 0.30 | 0.20 | 0.41 | | CholACE.stc2[2,1] | 0.04 | 0.00 | 0.11 | | CholACE.ste2[2,1] | 0.08 | 0.05 | 0.10 |
| CholACE.sta2[3,1] | 0.30 | 0.19 | 0.42 | | CholACE.stc2[3,1] | 0.04 | 0.00 | 0.14 | | CholACE.ste2[3,1] | 0.07 | 0.05 | 0.10 |
| CholACE.sta2[4,1] | 0.38 | 0.26 | 0.47 | | CholACE.stc2[4,1] | 0.02 | 0.00 | 0.09 | | CholACE.ste2[4,1] | 0.07 | 0.05 | 0.09 |
| CholACE.sta2[5,1] | 0.20 | 0.12 | 0.31 | | CholACE.stc2[5,1] | 0.07 | 0.00 | 0.15 | | CholACE.ste2[5,1] | 0.09 | 0.07 | 0.12 |
| CholACE.sta2[2,2] | 0.12 | 0.04 | 0.17 | | CholACE.stc2[2,2] | 0.00 | 0.00 | 0.05 | | CholACE.ste2[2,2] | 0.45 | 0.41 | 0.50 |
| CholACE.sta2[3,2] | 0.07 | 0.01 | 0.12 | | CholACE.stc2[3,2] | 0.01 | 0.00 | 0.06 | | CholACE.ste2[3,2] | 0.13 | 0.11 | 0.16 |
| CholACE.sta2[4,2] | 0.05 | 0.01 | 0.12 | | CholACE.stc2[4,2] | 0.00 | 0.00 | 0.04 | | CholACE.ste2[4,2] | 0.08 | 0.06 | 0.10 |
| CholACE.sta2[5,2] | 0.12 | 0.05 | 0.17 | | CholACE.stc2[5,2] | 0.00 | 0.00 | 0.05 | | CholACE.ste2[5,2] | 0.06 | 0.04 | 0.08 |
| CholACE.sta2[3,3] | 0.00 | 0.00 | 0.03 | | CholACE.stc2[3,3] | 0.00 | 0.00 | 0.03 | | CholACE.ste2[3,3] | 0.36 | 0.33 | 0.40 |
| CholACE.sta2[4,3] | 0.01 | 0.00 | 0.05 | | CholACE.stc2[4,3] | 0.00 | 0.00 | 0.04 | | CholACE.ste2[4,3] | 0.06 | 0.04 | 0.08 |
| CholACE.sta2[5,3] | 0.01 | 0.00 | 0.05 | | CholACE.stc2[5,3] | 0.00 | 0.00 | 0.03 | | CholACE.ste2[5,3] | 0.06 | 0.04 | 0.08 |
| CholACE.sta2[4,4] | 0.00 | 0.00 | 0.05 | | CholACE.stc2[4,4] | 0.00 | 0.00 | 0.03 | | CholACE.ste2[4,4] | 0.32 | 0.28 | 0.36 |
| CholACE.sta2[5,4] | 0.00 | 0.00 | 0.05 | | CholACE.stc2[5,4] | 0.00 | 0.00 | 0.03 | | CholACE.ste2[5,4] | 0.06 | 0.04 | 0.08 |
| CholACE.sta2[5,5] | 0.00 | 0.00 | 0.05 | | CholACE.stc2[5,5] | 0.00 | 0.00 | 0.03 | | CholACE.ste2[5,5] | 0.33 | 0.29 | 0.37 |

| **Depression** | | 95% CI | | | **Depression** | | 95% CI | | | **Depression** | | 95% CI | |
|---|---|---|---|---|---|---|---|---|---|---|---|---|---|
| | estimate | lower | upper | | | estimate | lower | upper | | | estimate | lower | upper |
| CholACE.sta2[1,1] | 0.30 | 0.21 | 0.36 | | CholACE.stc2[1,1] | 0.02 | 0.00 | 0.09 | | CholACE.ste2[1,1] | 0.68 | 0.63 | 0.72 |
| CholACE.sta2[2,1] | 0.35 | 0.26 | 0.42 | | CholACE.stc2[2,1] | 0.01 | 0.00 | 0.07 | | CholACE.ste2[2,1] | 0.06 | 0.04 | 0.08 |
| CholACE.sta2[3,1] | 0.29 | 0.19 | 0.38 | | CholACE.stc2[3,1] | 0.00 | 0.00 | 0.08 | | CholACE.ste2[3,1] | 0.08 | 0.05 | 0.10 |
| CholACE.sta2[4,1] | 0.34 | 0.23 | 0.43 | | CholACE.stc2[4,1] | 0.00 | 0.00 | 0.08 | | CholACE.ste2[4,1] | 0.08 | 0.05 | 0.10 |
| CholACE.sta2[5,1] | 0.29 | 0.18 | 0.39 | | CholACE.stc2[5,1] | 0.03 | 0.00 | 0.12 | | CholACE.ste2[5,1] | 0.06 | 0.04 | 0.08 |
| CholACE.sta2[2,2] | 0.03 | 0.00 | 0.07 | | CholACE.stc2[2,2] | 0.00 | 0.00 | 0.03 | | CholACE.ste2[2,2] | 0.55 | 0.51 | 0.60 |
| CholACE.sta2[3,2] | 0.05 | 0.00 | 0.11 | | CholACE.stc2[3,2] | 0.00 | 0.00 | 0.05 | | CholACE.ste2[3,2] | 0.15 | 0.12 | 0.18 |
| CholACE.sta2[4,2] | 0.04 | 0.00 | 0.11 | | CholACE.stc2[4,2] | 0.00 | 0.00 | 0.06 | | CholACE.ste2[4,2] | 0.08 | 0.06 | 0.10 |
| CholACE.sta2[5,2] | 0.05 | 0.00 | 0.11 | | CholACE.stc2[5,2] | 0.00 | 0.00 | 0.06 | | CholACE.ste2[5,2] | 0.07 | 0.05 | 0.10 |
| CholACE.sta2[3,3] | 0.00 | 0.00 | 0.04 | | CholACE.stc2[3,3] | 0.00 | 0.00 | 0.02 | | CholACE.ste2[3,3] | 0.43 | 0.38 | 0.47 |
| CholACE.sta2[4,3] | 0.01 | 0.00 | 0.05 | | CholACE.stc2[4,3] | 0.00 | 0.00 | 0.03 | | CholACE.ste2[4,3] | 0.06 | 0.04 | 0.08 |
| CholACE.sta2[5,3] | 0.00 | 0.00 | 0.05 | | CholACE.stc2[5,3] | 0.00 | 0.00 | 0.04 | | CholACE.ste2[5,3] | 0.05 | 0.03 | 0.07 |
| CholACE.sta2[4,4] | 0.00 | 0.00 | 0.05 | | CholACE.stc2[4,4] | 0.00 | 0.00 | 0.03 | | CholACE.ste2[4,4] | 0.39 | 0.35 | 0.44 |
| CholACE.sta2[5,4] | 0.00 | 0.00 | 0.05 | | CholACE.stc2[5,4] | 0.00 | 0.00 | 0.04 | | CholACE.ste2[5,4] | 0.05 | 0.04 | 0.07 |
| CholACE.sta2[5,5] | 0.00 | 0.00 | 0.04 | | CholACE.stc2[5,5] | 0.00 | 0.00 | 0.03 | | CholACE.ste2[5,5] | 0.40 | 0.36 | 0.44 |
| **General Anxiety** | | 95% CI | | | **General Anxiety** | | 95% CI | | | **General Anxiety** | | 95% CI | |
| | estimate | lower | upper | | | estimate | lower | upper | | | estimate | lower | upper |
| CholACE.sta2[1,1] | 0.33 | 0.26 | 0.37 | | CholACE.stc2[1,1] | 0.00 | 0.00 | 0.04 | | CholACE.ste2[1,1] | 0.67 | 0.62 | 0.72 |
| CholACE.sta2[2,1] | 0.33 | 0.24 | 0.42 | | CholACE.stc2[2,1] | 0.00 | 0.00 | 0.09 | | CholACE.ste2[2,1] | 0.08 | 0.06 | 0.11 |
| CholACE.sta2[3,1] | 0.30 | 0.20 | 0.38 | | CholACE.stc2[3,1] | 0.01 | 0.00 | 0.10 | | CholACE.ste2[3,1] | 0.10 | 0.08 | 0.13 |
| CholACE.sta2[4,1] | 0.33 | 0.22 | 0.41 | | CholACE.stc2[4,1] | 0.00 | 0.00 | 0.10 | | CholACE.ste2[4,1] | 0.09 | 0.06 | 0.12 |
| CholACE.sta2[5,1] | 0.33 | 0.24 | 0.43 | | CholACE.stc2[5,1] | 0.00 | 0.00 | 0.10 | | CholACE.ste2[5,1] | 0.08 | 0.06 | 0.11 |
| CholACE.sta2[2,2] | 0.09 | 0.00 | 0.15 | | CholACE.stc2[2,2] | 0.00 | 0.00 | 0.07 | | CholACE.ste2[2,2] | 0.48 | 0.43 | 0.53 |
| CholACE.sta2[3,2] | 0.03 | 0.00 | 0.08 | | CholACE.stc2[3,2] | 0.00 | 0.00 | 0.06 | | CholACE.ste2[3,2] | 0.16 | 0.13 | 0.19 |
| CholACE.sta2[4,2] | 0.04 | 0.00 | 0.11 | | CholACE.stc2[4,2] | 0.00 | 0.00 | 0.07 | | CholACE.ste2[4,2] | 0.10 | 0.07 | 0.13 |
| CholACE.sta2[5,2] | 0.09 | 0.00 | 0.15 | | CholACE.stc2[5,2] | 0.00 | 0.00 | 0.08 | | CholACE.ste2[5,2] | 0.06 | 0.04 | 0.09 |
| CholACE.sta2[3,3] | 0.01 | 0.00 | 0.04 | | CholACE.stc2[3,3] | 0.00 | 0.00 | 0.03 | | CholACE.ste2[3,3] | 0.37 | 0.33 | 0.41 |
| CholACE.sta2[4,3] | 0.02 | 0.00 | 0.06 | | CholACE.stc2[4,3] | 0.00 | 0.00 | 0.05 | | CholACE.ste2[4,3] | 0.06 | 0.04 | 0.08 |

| | | | | | | | | | | | | | |
|---|---|---|---|---|---|---|---|---|---|---|---|---|---|
| CholACE.sta2[5,3] | 0.00 | 0.00 | 0.04 | | CholACE.stc2[5,3] | 0.00 | 0.00 | 0.03 | | CholACE.ste2[5,3] | 0.06 | 0.04 | 0.08 |
| CholACE.sta2[4,4] | 0.00 | 0.00 | 0.05 | | CholACE.stc2[4,4] | 0.00 | 0.00 | 0.04 | | CholACE.ste2[4,4] | 0.34 | 0.31 | 0.38 |
| CholACE.sta2[5,4] | 0.00 | 0.00 | 0.04 | | CholACE.stc2[5,4] | 0.00 | 0.00 | 0.03 | | CholACE.ste2[5,4] | 0.05 | 0.03 | 0.07 |
| CholACE.sta2[5,5] | 0.00 | 0.00 | 0.04 | | CholACE.stc2[5,5] | 0.00 | 0.00 | 0.02 | | CholACE.ste2[5,5] | 0.30 | 0.27 | 0.34 |

Table S9. Polygenic score predictions. Educational Attainment GWAS (Lee et al. 2018)

| **T1** | | | | | | |
|---|---|---|---|---|---|---|
| Phenotype | BETA | SE | F | p | R2 | N |
| Emotional problems | -0.077 | 0.016 | 22.600 | 0.000 | 0.006 | 3750 |
| Conduct Problems | -0.101 | 0.016 | 38.350 | 0.000 | 0.010 | 3750 |
| Hyperactivity | -0.045 | 0.016 | 7.559 | 0.006 | 0.002 | 3749 |
| Peer problems | -0.133 | 0.016 | 67.650 | 0.000 | 0.017 | 3750 |
| Prosocial Behaviour | 0.020 | 0.016 | 1.478 | 0.224 | 0.000 | 3750 |
| Behaviour problems | -0.115 | 0.016 | 50.410 | 0.000 | 0.013 | 3750 |
| Depression | -0.066 | 0.016 | 16.160 | 0.000 | 0.004 | 3744 |
| General Anxiety | -0.007 | 0.017 | 0.178 | 0.673 | 0.000 | 3352 |
| **T2** | | | | | | |
| Phenotype | BETA | SE | F | p | R2 | N |
| Emotional problems | -0.020 | 0.023 | 0.750 | 0.387 | 0.000 | 1957 |
| Conduct Problems | -0.040 | 0.023 | 2.952 | 0.086 | 0.001 | 1957 |
| Hyperactivity | 0.017 | 0.023 | 0.557 | 0.456 | 0.000 | 1957 |
| Peer problems | -0.121 | 0.023 | 27.610 | 0.000 | 0.013 | 1957 |
| Prosocial Behaviour | -0.021 | 0.023 | 0.781 | 0.377 | 0.000 | 1957 |
| Behaviour problems | -0.047 | 0.023 | 4.107 | 0.043 | 0.002 | 1957 |

| | | | | | | |
|---|---|---|---|---|---|---|
| Depression | -0.013 | 0.023 | 0.313 | 0.576 | 0.000 | 1957 |
| General Anxiety | -0.006 | 0.023 | 0.065 | 0.799 | 0.000 | 1957 |
| **T3** | | | | | | |
| Phenotype | BETA | SE | F | p | R2 | N |
| Emotional problems | -0.048 | 0.025 | 3.751 | 0.053 | 0.002 | 1660 |
| Conduct Problems | -0.052 | 0.025 | 4.366 | 0.037 | 0.002 | 1660 |
| Hyperactivity | 0.001 | 0.025 | 0.001 | 0.973 | -0.001 | 1660 |
| Peer problems | -0.130 | 0.025 | 27.330 | 0.000 | 0.016 | 1660 |
| Prosocial Behaviour | -0.011 | 0.025 | 0.201 | 0.654 | 0.000 | 1660 |
| Behaviour problems | -0.072 | 0.025 | 8.356 | 0.004 | 0.004 | 1660 |
| Depression | -0.039 | 0.025 | 2.457 | 0.117 | 0.001 | 1660 |
| General Anxiety | 0.002 | 0.025 | 0.007 | 0.931 | -0.001 | 1660 |
| **T4** | | | | | | |
| Phenotype | BETA | SE | F | p | R2 | N |
| Emotional problems | -0.036 | 0.026 | 1.954 | 0.162 | 0.001 | 1516 |
| Conduct Problems | -0.032 | 0.026 | 1.531 | 0.216 | 0.000 | 1516 |
| Hyperactivity | 0.011 | 0.026 | 0.194 | 0.659 | -0.001 | 1516 |
| Peer problems | -0.124 | 0.026 | 23.070 | 0.000 | 0.014 | 1516 |
| Prosocial Behaviour | -0.006 | 0.026 | 0.060 | 0.807 | -0.001 | 1516 |
| Behaviour problems | -0.055 | 0.026 | 4.565 | 0.033 | 0.002 | 1516 |
| Depression | -0.034 | 0.026 | 1.718 | 0.190 | 0.000 | 1516 |
| General Anxiety | -0.009 | 0.026 | 0.119 | 0.730 | -0.001 | 1516 |
| **T5** | | | | | | |

| Phenotype | BETA | SE | F | p | R2 | N |
|---|---|---|---|---|---|---|
| Emotional problems | -0.060 | 0.025 | 5.625 | 0.018 | 0.003 | 1570 |
| Conduct Problems | -0.055 | 0.025 | 4.845 | 0.028 | 0.002 | 1570 |
| Hyperactivity | -0.039 | 0.025 | 2.422 | 0.120 | 0.001 | 1570 |
| Peer problems | -0.147 | 0.025 | 35.040 | 0.000 | 0.021 | 1570 |
| Prosocial Behaviour | -0.005 | 0.025 | 0.047 | 0.829 | -0.001 | 1570 |
| Behaviour problems | -0.098 | 0.025 | 15.260 | 0.000 | 0.009 | 1570 |
| Depression | -0.053 | 0.025 | 4.454 | 0.035 | 0.002 | 1570 |
| General Anxiety | -0.046 | 0.025 | 3.415 | 0.065 | 0.002 | 1570 |

Table S10. Polygenic score predictions. Genomic p factor

| **T1** | | | | | | |
|---|---|---|---|---|---|---|
| Phenotype | BETA | SE | F | p | R2 | N |
| Emotional problems | 0.146 | 0.016 | 80.450 | < 2.2e-16 | 0.021 | 3750 |
| Conduct Problems | 0.075 | 0.016 | 20.920 | 0.000 | 0.005 | 3750 |
| Hyperactivity | 0.079 | 0.016 | 23.260 | 0.000 | 0.006 | 3749 |
| Peer problems | 0.095 | 0.016 | 33.890 | 0.000 | 0.009 | 3750 |
| Prosocial Behaviour | 0.004 | 0.016 | 0.061 | 0.805 | 0.000 | 3750 |
| Behaviour problems | 0.141 | 0.016 | 74.920 | < 2.2e-16 | 0.019 | 3750 |
| Depression | 0.134 | 0.016 | 67.650 | 0.000 | 0.017 | 3744 |
| General Anxiety | 0.117 | 0.017 | 45.850 | 0.000 | 0.013 | 3352 |
| **T2** | | | | | | |
| Phenotype | BETA | SE | F | p | R2 | N |

| | | | | | | |
|---|---|---|---|---|---|---|
| Emotional problems | 0.095 | 0.023 | 17.470 | 0.000 | 0.008 | 1957 |
| Conduct Problems | 0.023 | 0.023 | 1.037 | 0.309 | 0.000 | 1957 |
| Hyperactivity | 0.090 | 0.023 | 15.370 | 0.000 | 0.007 | 1957 |
| Peer problems | 0.068 | 0.023 | 8.892 | 0.003 | 0.004 | 1957 |
| Prosocial Behaviour | 0.027 | 0.023 | 1.411 | 0.235 | 0.000 | 1957 |
| Behaviour problems | 0.106 | 0.023 | 21.740 | 0.000 | 0.010 | 1957 |
| Depression | 0.076 | 0.023 | 11.170 | 0.001 | 0.005 | 1957 |
| General Anxiety | 0.102 | 0.023 | 19.780 | 0.000 | 0.010 | 1957 |
| **T3** | | | | | | |
| Phenotype | BETA | SE | F | p | R2 | N |
| Emotional problems | 0.123 | 0.025 | 24.420 | 0.000 | 0.014 | 1660 |
| Conduct Problems | 0.054 | 0.025 | 4.647 | 0.031 | 0.002 | 1660 |
| Hyperactivity | 0.057 | 0.025 | 5.112 | 0.024 | 0.002 | 1660 |
| Peer problems | 0.043 | 0.025 | 2.917 | 0.088 | 0.001 | 1660 |
| Prosocial Behaviour | 0.011 | 0.025 | 0.192 | 0.661 | 0.000 | 1660 |
| Behaviour problems | 0.104 | 0.025 | 17.430 | 0.000 | 0.010 | 1660 |
| Depression | 0.098 | 0.025 | 15.530 | 0.000 | 0.009 | 1660 |
| General Anxiety | 0.130 | 0.025 | 27.470 | 0.000 | 0.016 | 1660 |
| **T4** | | | | | | |
| Phenotype | BETA | SE | F | p | R2 | N |
| Emotional problems | 0.091 | 0.026 | 11.950 | 0.001 | 0.007 | 1516 |
| Conduct Problems | 0.038 | 0.026 | 2.051 | 0.152 | 0.001 | 1516 |
| Hyperactivity | 0.048 | 0.026 | 3.310 | 0.069 | 0.002 | 1516 |
| Peer problems | 0.025 | 0.026 | 0.907 | 0.341 | 0.000 | 1516 |

| | | | | | | |
|---|---|---|---|---|---|---|
| Prosocial Behaviour | 0.028 | 0.026 | 1.126 | 0.289 | 0.000 | 1516 |
| Behaviour problems | 0.076 | 0.026 | 8.173 | 0.004 | 0.005 | 1516 |
| Depression | 0.090 | 0.026 | 11.570 | 0.001 | 0.007 | 1516 |
| General Anxiety | 0.099 | 0.026 | 14.100 | 0.000 | 0.009 | 1516 |
| **T5** | | | | | | |
| Phenotype | BETA | SE | F | p | R2 | N |
| Emotional problems | 0.109 | 0.025 | 18.400 | 0.000 | 0.011 | 1570 |
| Conduct Problems | 0.050 | 0.026 | 3.888 | 0.049 | 0.002 | 1570 |
| Hyperactivity | 0.018 | 0.026 | 0.513 | 0.474 | 0.000 | 1570 |
| Peer problems | 0.041 | 0.026 | 2.603 | 0.107 | 0.001 | 1570 |
| Prosocial Behaviour | 0.015 | 0.026 | 0.338 | 0.561 | 0.000 | 1570 |
| Behaviour problems | 0.080 | 0.026 | 9.698 | 0.002 | 0.006 | 1570 |
| Depression | 0.086 | 0.026 | 11.290 | 0.001 | 0.007 | 1570 |
| General Anxiety | 0.093 | 0.026 | 13.370 | 0.000 | 0.008 | 1570 |

Table S11. Polygenic score predictions. Cross-disorder GWAS (Lee et al. 2019)

| **T1** | | | | | | |
|---|---|---|---|---|---|---|
| Phenotype | BETA | SE | F | p | R2 | N |
| Emotional problems | 0.074 | 0.016 | 20.490 | 0.000 | 0.005 | 3750 |
| Conduct Problems | 0.044 | 0.016 | 7.245 | 0.007 | 0.002 | 3750 |
| Hyperactivity | 0.041 | 0.016 | 6.343 | 0.012 | 0.001 | 3750 |
| Peer problems | 0.029 | 0.016 | 3.061 | 0.080 | 0.001 | 3750 |
| Prosocial Behaviour | 0.005 | 0.016 | 0.110 | 0.741 | 0.000 | 3750 |

| | | | | | | |
|---|---|---|---|---|---|---|
| Behaviour problems | 0.068 | 0.016 | 16.930 | 0.000 | 0.004 | 3750 |
| Depression | 0.074 | 0.016 | 20.360 | 0.000 | 0.005 | 3744 |
| General Anxiety | 0.071 | 0.017 | 16.810 | 0.000 | 0.005 | 3352 |
| **T2** | | | | | | |
| Phenotype | BETA | SE | F | p | R2 | N |
| Emotional problems | 0.035 | 0.023 | 2.297 | 0.130 | 0.001 | 1957 |
| Conduct Problems | -0.012 | 0.023 | 0.298 | 0.586 | 0.000 | 1957 |
| Hyperactivity | 0.009 | 0.023 | 0.140 | 0.709 | 0.000 | 1957 |
| Peer problems | 0.009 | 0.023 | 0.159 | 0.690 | 0.000 | 1957 |
| Prosocial Behaviour | 0.015 | 0.023 | 0.439 | 0.508 | 0.000 | 1957 |
| Behaviour problems | 0.020 | 0.023 | 0.751 | 0.386 | 0.000 | 1957 |
| Depression | 0.022 | 0.023 | 0.914 | 0.339 | 0.000 | 1957 |
| General Anxiety | 0.039 | 0.023 | 2.905 | 0.088 | 0.001 | 1957 |
| **T3** | | | | | | |
| Phenotype | BETA | SE | F | p | R2 | N |
| Emotional problems | 0.046 | 0.025 | 3.533 | 0.060 | 0.002 | 1660 |
| Conduct Problems | 0.009 | 0.025 | 0.133 | 0.715 | -0.001 | 1660 |
| Hyperactivity | 0.024 | 0.025 | 0.914 | 0.339 | 0.000 | 1660 |
| Peer problems | 0.001 | 0.025 | 0.001 | 0.972 | -0.001 | 1660 |
| Prosocial Behaviour | 0.030 | 0.025 | 1.449 | 0.229 | 0.000 | 1660 |
| Behaviour problems | 0.033 | 0.025 | 1.814 | 0.178 | 0.000 | 1660 |
| Depression | 0.038 | 0.025 | 2.412 | 0.121 | 0.001 | 1660 |
| General Anxiety | 0.079 | 0.025 | 10.380 | 0.001 | 0.006 | 1660 |

| **T4** | | | | | | |
|---|---|---|---|---|---|---|
| Phenotype | BETA | SE | F | p | R2 | N |
| Emotional problems | 0.025 | 0.026 | 0.925 | 0.336 | 0.000 | 1516 |
| Conduct Problems | -0.027 | 0.026 | 1.112 | 0.292 | 0.000 | 1516 |
| Hyperactivity | -0.004 | 0.026 | 0.023 | 0.879 | -0.001 | 1516 |
| Peer problems | -0.014 | 0.026 | 0.283 | 0.595 | 0.000 | 1516 |
| Prosocial Behaviour | 0.025 | 0.026 | 0.884 | 0.347 | 0.000 | 1516 |
| Behaviour problems | 0.001 | 0.026 | 0.001 | 0.979 | -0.001 | 1516 |
| Depression | 0.026 | 0.026 | 1.006 | 0.316 | 0.000 | 1516 |
| General Anxiety | 0.014 | 0.026 | 0.272 | 0.602 | 0.000 | 1516 |
| **T5** | | | | | | |
| Phenotype | BETA | SE | F | p | R2 | N |
| Emotional problems | 0.048 | 0.025 | 3.749 | 0.053 | 0.002 | 1570 |
| Conduct Problems | 0.029 | 0.025 | 1.348 | 0.246 | 0.000 | 1570 |
| Hyperactivity | 0.024 | 0.025 | 0.957 | 0.328 | 0.000 | 1570 |
| Peer problems | 0.015 | 0.025 | 0.369 | 0.544 | 0.000 | 1570 |
| Prosocial Behaviour | 0.022 | 0.025 | 0.783 | 0.377 | 0.000 | 1570 |
| Behaviour problems | 0.042 | 0.025 | 2.831 | 0.093 | 0.001 | 1570 |
| Depression | 0.030 | 0.025 | 1.467 | 0.226 | 0.000 | 1570 |
| General Anxiety | 0.042 | 0.025 | 2.887 | 0.089 | 0.001 | 1570 |

Table S12. Polygenic score predictions. Anxiety disorders GWAS (Purves et al. 2019)

| **T1** | | | | | | |
|---|---|---|---|---|---|---|
| Phenotype | BETA | SE | F | p | R2 | N |

| | | | | | | |
|---|---|---|---|---|---|---|
| Emotional problems | -0.084 | 0.016 | 26.240 | 0.000 | 0.007 | 3750 |
| Conduct Problems | -0.023 | 0.016 | 2.016 | 0.156 | 0.000 | 3750 |
| Hyperactivity | -0.048 | 0.016 | 8.661 | 0.003 | 0.002 | 3750 |
| Peer problems | 0.059 | 0.016 | 12.980 | 0.000 | 0.003 | 3750 |
| Prosocial Behaviour | 0.003 | 0.016 | 0.034 | 0.854 | 0.000 | 3750 |
| Behaviour problems | -0.079 | 0.016 | 23.100 | 0.000 | 0.006 | 3750 |
| Depression | -0.085 | 0.016 | 26.730 | 0.000 | 0.007 | 3744 |
| General Anxiety | -0.064 | 0.017 | 13.300 | 0.000 | 0.004 | 3352 |
| **T2** | | | | | | |
| Phenotype | BETA | SE | F | p | R2 | N |
| Emotional problems | -0.054 | 0.023 | 5.847 | 0.016 | 0.002 | 1957 |
| Conduct Problems | -0.027 | 0.023 | 1.402 | 0.237 | 0.000 | 1957 |
| Hyperactivity | -0.062 | 0.023 | 7.577 | 0.006 | 0.003 | 1957 |
| Peer problems | -0.024 | 0.023 | 1.129 | 0.288 | 0.000 | 1957 |
| Prosocial Behaviour | -0.015 | 0.023 | 0.427 | 0.514 | 0.000 | 1957 |
| Behaviour problems | -0.063 | 0.023 | 7.946 | 0.005 | 0.004 | 1957 |
| Depression | -0.041 | 0.023 | 3.266 | 0.071 | 0.001 | 1957 |
| General Anxiety | -0.068 | 0.023 | 9.255 | 0.002 | 0.004 | 1957 |
| **T3** | | | | | | |
| Phenotype | BETA | SE | F | p | R2 | N |
| Emotional problems | -0.059 | 0.025 | 5.448 | 0.020 | 0.003 | 1660 |
| Conduct Problems | -0.011 | 0.026 | 0.189 | 0.664 | 0.000 | 1660 |
| Hyperactivity | -0.030 | 0.025 | 1.355 | 0.245 | 0.000 | 1660 |
| Peer problems | -0.021 | 0.026 | 0.658 | 0.418 | 0.000 | 1660 |

| | | | | | | |
|---|---|---|---|---|---|---|
| Prosocial Behaviour | -0.019 | 0.026 | 0.535 | 0.465 | 0.000 | 1660 |
| Behaviour problems | -0.048 | 0.025 | 3.575 | 0.059 | 0.002 | 1660 |
| Depression | -0.031 | 0.025 | 1.498 | 0.221 | 0.000 | 1660 |
| General Anxiety | -0.079 | 0.025 | 9.649 | 0.002 | 0.005 | 1660 |
| **T4** | | | | | | |
| Phenotype | BETA | SE | F | p | R2 | N |
| Emotional problems | -0.024 | 0.026 | 0.823 | 0.364 | 0.000 | 1516 |
| Conduct Problems | -0.051 | 0.026 | 3.874 | 0.049 | 0.002 | 1516 |
| Hyperactivity | 0.002 | 0.026 | 0.008 | 0.930 | -0.001 | 1516 |
| Peer problems | 0.023 | 0.026 | 0.804 | 0.370 | 0.000 | 1516 |
| Prosocial Behaviour | -0.023 | 0.026 | 0.802 | 0.371 | 0.000 | 1516 |
| Behaviour problems | -0.013 | 0.026 | 0.254 | 0.615 | 0.000 | |
| Depression | -0.037 | 0.026 | 2.044 | 0.153 | 0.001 | |
| General Anxiety | -0.053 | 0.026 | 4.194 | 0.041 | 0.002 | 1516 |
| **T5** | | | | | | |
| Phenotype | BETA | SE | F | p | R2 | N |
| Emotional problems | -0.065 | 0.025 | 6.560 | 0.011 | 0.004 | 1570 |
| Conduct Problems | -0.036 | 0.025 | 2.040 | 0.154 | 0.001 | 1570 |
| Hyperactivity | 0.009 | 0.025 | 0.138 | 0.710 | -0.001 | 1570 |
| Peer problems | -0.004 | 0.025 | 0.026 | 0.872 | -0.001 | 1570 |
| Prosocial Behaviour | -0.019 | 0.025 | 0.590 | 0.443 | 0.000 | 1570 |
| Behaviour problems | -0.034 | 0.025 | 1.841 | 0.175 | 0.001 | 1570 |
| Depression | -0.036 | 0.025 | 1.968 | 0.161 | 0.001 | 1570 |

| | | | | | | |
|---|---|---|---|---|---|---|
| General Anxiety | -0.060 | 0.025 | 5.727 | 0.017 | 0.003 | 1570 |

Table S13. Polygenic score predictions. Major depression GWAS (Wray et al 2018)

| **T1** | | | | | | |
|---|---|---|---|---|---|---|
| Phenotype | BETA | SE | F | p | R2 | N |
| Emotional problems | 0.089 | 0.016 | 30.160 | 0.000 | 0.008 | 3750 |
| Conduct Problems | 0.045 | 0.016 | 7.647 | 0.006 | 0.002 | 3750 |
| Hyperactivity | 0.045 | 0.016 | 7.600 | 0.006 | 0.002 | 3750 |
| Peer problems | 0.068 | 0.016 | 17.390 | 0.000 | 0.004 | 3750 |
| Prosocial Behaviour | -0.003 | 0.016 | 0.030 | 0.862 | 0.000 | 3750 |
| Behaviour problems | 0.088 | 0.016 | 29.110 | 0.000 | 0.007 | 3750 |
| Depression | 0.078 | 0.016 | 22.790 | 0.000 | 0.006 | 3744 |
| General Anxiety | 0.062 | 0.017 | 13.100 | 0.000 | 0.004 | 3352 |
| **T2** | | | | | | |
| Phenotype | BETA | SE | F | p | R2 | N |
| Emotional problems | 0.057 | 0.023 | 6.237 | 0.013 | 0.003 | 1957 |
| Conduct Problems | -0.010 | 0.023 | 0.174 | 0.676 | 0.000 | 1957 |
| Hyperactivity | 0.057 | 0.023 | 6.372 | 0.012 | 0.003 | 1957 |
| Peer problems | 0.063 | 0.023 | 7.679 | 0.006 | 0.003 | 1957 |
| Prosocial Behaviour | 0.052 | 0.023 | 5.307 | 0.021 | 0.002 | 1957 |
| Behaviour problems | 0.067 | 0.023 | 8.600 | 0.003 | 0.004 | 1957 |
| Depression | 0.056 | 0.023 | 6.102 | 0.014 | 0.003 | 1957 |
| General Anxiety | 0.053 | 0.023 | 5.503 | 0.019 | 0.002 | 1957 |

| **T3** | | | | | | |
|---|---|---|---|---|---|---|
| Phenotype | BETA | SE | F | p | R2 | N |
| Emotional problems | 0.085 | 0.024 | 12.200 | 0.000 | 0.007 | 1660 |
| Conduct Problems | 0.053 | 0.024 | 4.743 | 0.030 | 0.002 | 1660 |
| Hyperactivity | 0.041 | 0.024 | 2.760 | 0.097 | 0.001 | 1660 |
| Peer problems | 0.031 | 0.024 | 1.569 | 0.211 | 0.000 | 1660 |
| Prosocial Behaviour | 0.024 | 0.024 | 0.965 | 0.326 | 0.000 | 1660 |
| Behaviour problems | 0.076 | 0.024 | 9.761 | 0.002 | 0.005 | 1660 |
| Depression | 0.066 | 0.024 | 7.363 | 0.007 | 0.004 | 1660 |
| General Anxiety | 0.079 | 0.024 | 10.520 | 0.001 | 0.006 | 1660 |
| **T4** | | | | | | |
| Phenotype | BETA | SE | F | p | R2 | N |
| Emotional problems | 0.073 | 0.026 | 8.026 | 0.005 | 0.005 | 1516 |
| Conduct Problems | 0.005 | 0.026 | 0.032 | 0.858 | -0.001 | 1516 |
| Hyperactivity | 0.052 | 0.026 | 4.128 | 0.042 | 0.002 | 1516 |
| Peer problems | 0.013 | 0.026 | 0.234 | 0.629 | -0.001 | 1516 |
| Prosocial Behaviour | 0.047 | 0.026 | 3.247 | 0.072 | 0.001 | 1516 |
| Behaviour problems | 0.059 | 0.026 | 5.138 | 0.024 | 0.003 | 1516 |
| Depression | 0.064 | 0.026 | 6.098 | 0.014 | 0.014 | 1516 |
| General Anxiety | 0.050 | 0.026 | 3.739 | 0.053 | 0.002 | 1516 |
| **T5** | | | | | | |
| Phenotype | BETA | SE | F | p | R2 | N |
| Emotional problems | 0.081 | 0.025 | 10.380 | 0.001 | 0.006 | 1570 |
| Conduct Problems | -0.004 | 0.025 | 0.026 | 0.873 | -0.001 | 1570 |

| | | | | | | |
|---|---|---|---|---|---|---|
| Hyperactivity | 0.004 | 0.025 | 0.024 | 0.878 | -0.001 | 1570 |
| Peer problems | 0.018 | 0.025 | 0.527 | 0.468 | 0.000 | 1570 |
| Prosocial Behaviour | 0.024 | 0.025 | 0.947 | 0.331 | 0.000 | 1570 |
| Behaviour problems | 0.043 | 0.025 | 2.919 | 0.088 | 0.001 | 1570 |
| Depression | 0.055 | 0.025 | 4.812 | 0.028 | 0.002 | 1570 |
| General Anxiety | 0.055 | 0.025 | 4.767 | 0.029 | 0.002 | 1570 |

Table S14. Polygenic score predictions. Risk tolerance and risky behaviours GWAS (Linner et al 2019)

| **T1** | | | | | | |
|---|---|---|---|---|---|---|
| Phenotype | BETA | SE | F | p | R2 | N |
| Emotional problems | -0.025 | 0.016 | 2.284 | 0.131 | 0.000 | 3750 |
| Conduct Problems | 0.069 | 0.016 | 17.600 | 0.000 | 0.004 | 3750 |
| Hyperactivity | 0.059 | 0.016 | 12.880 | 0.000 | 0.003 | 3750 |
| Peer problems | 0.068 | 0.016 | 17.390 | 0.000 | 0.004 | 3750 |
| Prosocial Behaviour | 0.012 | 0.017 | 0.501 | 0.479 | 0.000 | 3750 |
| Behaviour problems | 0.020 | 0.016 | 1.537 | 0.215 | 0.000 | 3750 |
| Depression | -0.001 | 0.017 | 0.005 | 0.944 | 0.000 | 3744 |
| General Anxiety | 0.006 | 0.017 | 0.137 | 0.712 | 0.000 | 3352 |
| **T2** | | | | | | |
| Phenotype | BETA | SE | F | p | R2 | N |
| Emotional problems | 0.003 | 0.023 | 0.015 | 0.902 | -0.001 | 1957 |
| Conduct Problems | 0.045 | 0.023 | 3.947 | 0.047 | 0.002 | 1957 |
| Hyperactivity | 0.019 | 0.023 | 0.679 | 0.410 | 0.000 | 1957 |
| Peer problems | -0.008 | 0.023 | 0.135 | 0.713 | 0.000 | 1957 |

| | | | | | | |
|---|---|---|---|---|---|---|
| Prosocial Behaviour | 0.018 | 0.023 | 0.620 | 0.431 | 0.000 | 1957 |
| Behaviour problems | 0.016 | 0.023 | 0.494 | 0.482 | 0.000 | 1957 |
| Depression | 0.015 | 0.023 | 0.431 | 0.511 | 0.000 | 1957 |
| General Anxiety | 0.012 | 0.023 | 0.273 | 0.601 | 0.000 | 1957 |
| **T3** | | | | | | |
| Phenotype | BETA | SE | F | p | R2 | N |
| Emotional problems | -0.005 | 0.025 | 0.037 | 0.848 | -0.001 | 1660 |
| Conduct Problems | 0.046 | 0.025 | 3.412 | 0.065 | 0.001 | 1660 |
| Hyperactivity | 0.023 | 0.025 | 0.860 | 0.354 | 0.000 | 1660 |
| Peer problems | 0.002 | 0.025 | 0.009 | 0.926 | -0.001 | 1660 |
| Prosocial Behaviour | -0.029 | 0.025 | 1.322 | 0.250 | 0.000 | 1660 |
| Behaviour problems | 0.017 | 0.025 | 0.468 | 0.494 | 0.000 | 1660 |
| Depression | 0.022 | 0.025 | 0.742 | 0.389 | 0.000 | 1660 |
| General Anxiety | 0.013 | 0.025 | 0.252 | 0.616 | 0.000 | 1660 |
| **T4** | | | | | | |
| Phenotype | BETA | SE | F | p | R2 | N |
| Emotional problems | -0.035 | 0.026 | 1.921 | 0.166 | 0.001 | 1516 |
| Conduct Problems | 0.014 | 0.026 | 0.296 | 0.586 | 0.000 | 1516 |
| Hyperactivity | 0.001 | 0.026 | 0.002 | 0.961 | -0.001 | 1516 |
| Peer problems | -0.042 | 0.026 | 2.757 | 0.097 | 0.001 | 1516 |
| Prosocial Behaviour | 0.039 | 0.026 | 2.379 | 0.123 | 0.001 | 1516 |
| Behaviour problems | -0.026 | 0.026 | 1.022 | 0.312 | 0.000 | 1516 |
| Depression | -0.004 | 0.026 | 0.023 | 0.881 | -0.001 | 1516 |

| | | | | | | |
|---|---|---|---|---|---|---|
| General Anxiety | -0.019 | 0.026 | 0.560 | 0.454 | 0.000 | 1516 |
| **T5** | | | | | | |
| Phenotype | BETA | SE | F | p | R2 | N |
| Emotional problems | 0.012 | 0.025 | 0.238 | 0.626 | 0.000 | 1570 |
| Conduct Problems | 0.076 | 0.025 | 9.065 | 0.003 | 0.005 | 1570 |
| Hyperactivity | 0.039 | 0.025 | 2.344 | 0.126 | 0.001 | 1570 |
| Peer problems | -0.035 | 0.025 | 1.920 | 0.166 | 0.001 | 1570 |
| Prosocial Behaviour | -0.006 | 0.025 | 0.058 | 0.810 | -0.001 | 1570 |
| Behaviour problems | 0.000 | 0.025 | 1.034 | 0.309 | 0.000 | 1570 |
| Depression | 0.037 | 0.025 | 2.143 | 0.143 | 0.001 | 1570 |
| General Anxiety | -0.003 | 0.025 | 0.011 | 0.011 | -0.001 | 1570 |

Tables S15. Descriptive statistics for individuals with pre-existing mental health problems (+1SD on phenotypic p factor) and for those scoring low on mental health symptoms (+1SD on phenotypic p factor)

| (-1SD) on p phenotypic p | | | | | | | (+1SD) on p phenotypic p | | | | |
|---|---|---|---|---|---|---|---|---|---|---|---|
| **T1** | | | | | | | **T1** | | | | |
| | n | mean | sd | se | | | | n | mean | sd | se |
| SDQ Emotional problems | 329 | 1.49 | 1.74 | 0.1 | | | SDQ Emotional problems | 376 | 6.15 | 2.54 | 0.13 |
| SDQ Conduct | 329 | 1.14 | 0.99 | 0.05 | | | SDQ Conduct | 376 | 1.89 | 1.44 | 0.07 |
| SDQ Hyperactivity | 329 | 1.89 | 1.58 | 0.09 | | | SDQ Hyperactivity | 376 | 4.72 | 2.27 | 0.12 |
| SDQ Peer problems | 329 | 1.38 | 1.41 | 0.08 | | | SDQ Peer problems | 376 | 3.23 | 2.02 | 0.1 |
| SDQ Prosocial | 329 | 7.8 | 1.91 | 0.11 | | | SDQ Prosocial | 376 | 7.7 | 1.97 | 0.1 |
| SDQ Behaviour problems | 329 | 5.9 | 3.65 | 0.2 | | | SDQ Behaviour problems | 376 | 15.99 | 5.84 | 0.3 |
| Depression | 328 | 1.53 | 2.38 | 0.13 | | | Depression | 376 | 8.3 | 4.32 | 0.22 |

| | | | | | | | | | | | |
|---|---|---|---|---|---|---|---|---|---|---|---|
| General Anxiety | 307 | 2.86 | 3.85 | 0.22 | | | General Anxiety | 363 | 14.35 | 9.01 | 0.47 |
| Self Harm | 307 | 1.03 | 0.18 | 0.01 | | | Self Harm | 358 | 1.6 | 1.18 | 0.06 |
| T2 | | | | | | | T2 | | | | |
| | n | mean | sd | se | | | | n | mean | sd | se |
| SDQ Emotional problems | 368 | 0.35 | 0.59 | 0.03 | | | SDQ Emotional problems | 429 | 7.21 | 1.69 | 0.08 |
| SDQ Conduct | 368 | 0.86 | 0.64 | 0.03 | | | SDQ Conduct | 429 | 2.4 | 1.57 | 0.08 |
| SDQ Hyperactivity | 368 | 1.57 | 1.16 | 0.06 | | | SDQ Hyperactivity | 429 | 6.6 | 1.72 | 0.08 |
| SDQ Peer problems | 368 | 1.22 | 1.21 | 0.06 | | | SDQ Peer problems | 429 | 3.42 | 1.79 | 0.09 |
| SDQ Prosocial | 368 | 7.02 | 2.01 | 0.1 | | | SDQ Prosocial | 429 | 7.01 | 1.87 | 0.09 |
| SDQ Behaviour problems | 368 | 4 | 1.79 | 0.09 | | | SDQ Behaviour problems | 429 | 19.64 | 3.66 | 0.18 |
| Depression | 368 | 0.42 | 0.74 | 0.04 | | | Depression | 429 | 10.79 | 2.96 | 0.14 |
| General Anxiety | 368 | 1.74 | 1.76 | 0.09 | | | General Anxiety | 429 | 20.88 | 6.58 | 0.32 |
| Self Harm | 365 | 1 | 0.05 | 0 | | | Self Harm | 421 | 1.21 | 0.58 | 0.03 |
| T3 | | | | | | | T3 | | | | |
| | n | mean | sd | se | | | | n | mean | sd | se |
| SDQ Emotional problems | 244 | 0.98 | 1.35 | 0.09 | | | SDQ Emotional problems | 281 | 6.72 | 2.25 | 0.13 |
| SDQ Conduct | 244 | 1 | 0.82 | 0.05 | | | SDQ Conduct | 281 | 2.06 | 1.47 | 0.09 |
| SDQ Hyperactivity | 244 | 2.15 | 1.56 | 0.1 | | | SDQ Hyperactivity | 281 | 6.18 | 1.92 | 0.11 |
| SDQ Peer problems | 244 | 1.37 | 1.32 | 0.08 | | | SDQ Peer problems | 281 | 3.55 | 1.83 | 0.11 |
| SDQ Prosocial | 244 | 7.15 | 2.03 | 0.13 | | | SDQ Prosocial | 281 | 7.04 | 1.92 | 0.11 |
| SDQ Behaviour problems | 244 | 5.5 | 3.14 | 0.2 | | | SDQ Behaviour problems | 281 | 18.51 | 4.85 | 0.29 |
| Depression | 244 | 0.96 | 1.35 | 0.09 | | | Depression | 281 | 9.33 | 3.98 | 0.24 |
| General Anxiety | 244 | 2.7 | 2.86 | 0.18 | | | General Anxiety | 281 | 18.83 | 8.3 | 0.49 |
| Self Harm | 243 | 1 | 0 | 0 | | | Self Harm | 271 | 1.23 | 0.59 | 0.04 |
| T4 | | | | | | | T4 | | | | |
| | n | mean | sd | se | | | | n | mean | sd | se |
| SDQ Emotional problems | 227 | 1.01 | 1.29 | 0.09 | | | SDQ Emotional problems | 265 | 6.71 | 2.34 | 0.14 |

| | | | | | | | | | | | |
|---|---|---|---|---|---|---|---|---|---|---|---|
| SDQ Conduct | 227 | 0.99 | 0.78 | 0.05 | | | SDQ Conduct | 265 | 2.03 | 1.42 | 0.09 |
| SDQ Hyperactivity | 227 | 2 | 1.6 | 0.11 | | | SDQ Hyperactivity | 265 | 5.99 | 1.96 | 0.12 |
| SDQ Peer problems | 227 | 1.42 | 1.32 | 0.09 | | | SDQ Peer problems | 265 | 3.56 | 1.83 | 0.11 |
| SDQ Prosocial | 227 | 7.16 | 1.84 | 0.12 | | | SDQ Prosocial | 265 | 7.16 | 2.01 | 0.12 |
| SDQ Behaviour problems | 227 | 5.41 | 2.92 | 0.19 | | | SDQ Behaviour problems | 265 | 18.29 | 5.13 | 0.32 |
| Depression | 227 | 1.02 | 1.65 | 0.11 | | | Depression | 265 | 9.64 | 4.14 | 0.25 |
| General Anxiety | 227 | 3.05 | 3.56 | 0.24 | | | General Anxiety | 265 | 19.13 | 8.79 | 0.54 |
| Self Harm | 225 | 1 | 0 | 0 | | | Self Harm | 256 | 1.27 | 0.72 | 0.05 |
| **T5** | | | | | | | T5 | | | | |
| | n | mean | sd | se | | | | n | mean | sd | se |
| SDQ Emotional problems | 224 | 1.18 | 1.45 | 0.1 | | | SDQ Emotional problems | 272 | 6.33 | 2.46 | 0.15 |
| SDQ Conduct | 224 | 0.95 | 0.77 | 0.05 | | | SDQ Conduct | 272 | 1.88 | 1.46 | 0.09 |
| SDQ Hyperactivity | 224 | 2.06 | 1.67 | 0.11 | | | SDQ Hyperactivity | 272 | 5.67 | 2.2 | 0.13 |
| SDQ Peer problems | 224 | 1.67 | 1.46 | 0.1 | | | SDQ Peer problems | 272 | 3.47 | 1.98 | 0.12 |
| SDQ Prosocial | 224 | 7 | 2.02 | 0.13 | | | SDQ Prosocial | 272 | 7 | 1.93 | 0.12 |
| SDQ Behaviour problems | 224 | 5.86 | 3.45 | 0.23 | | | SDQ Behaviour problems | 272 | 17.35 | 5.7 | 0.35 |
| Depression | 224 | 1.21 | 1.81 | 0.12 | | | Depression | 272 | 8.75 | 4.24 | 0.26 |
| General Anxiety | 224 | 3.21 | 3.25 | 0.22 | | | General Anxiety | 272 | 17.78 | 8.67 | 0.53 |
| Self Harm | 223 | 1 | 0 | 0 | | | Self Harm | 267 | 1.27 | 0.7 | 0.04 |

Tables S16. MANOVA results for individuals with pre-existing mental health problems compared to the rest of the sample

| | **Time** | | | **Mental health** | | | **Time * Mental health** | | |
|---|---|---|---|---|---|---|---|---|---|
| | **F** | **p** | **Partial Eta squared** | **F** | **p** | **Partial Eta squared** | **F** | **p** | **Partial Eta squared** |
| SDQ Emotional problems | 3.068 | 0.016 | 0.003 | 607.48 | 0.001 | 0.39 | 30.483 | 0.001 | 0.031 |
| SDQ Conduct | 5.6 | 0.001 | 0.006 | 95.667 | 0.001 | 0.091 | 6.098 | 0.001 | 0.006 |
| SDQ Hyperactivity | 76.726 | 0.001 | 0.075 | 316.048 | 0.001 | 0.249 | 14.252 | 0.001 | 0.015 |
| SDQ Peer problems | 5.302 | 0.001 | 0.006 | 161.75 | 0.001 | 0.145 | 2.84 | 0.026 | 0.003 |

| | | | | | | | | | |
|---|---|---|---|---|---|---|---|---|---|
| SDQ Prosocial | 38.322 | 0.001 | 0.039 | 0.451 | 0.502 | 0 | 1.585 | 0.172 | 0.002 |
| SDQ Behaviour problems | 28.878 | 0.001 | 29 | 632.876 | 0.001 | 0.399 | 30.348 | 0.001 | 0.031 |
| Depression | 20.709 | 0.001 | 0.021 | 684.068 | 0.001 | 0.418 | 37.524 | 0.001 | 0.038 |
| General Anxiety | 45.146 | 0.001 | 0.045 | 710.344 | 0.001 | 0.427 | 37.142 | 0.001 | 0.038 |

Tables S17. Pairwise comparisons between T1-T5 for individuals with pre-existing mental health problems (+1SD on phenotypic p factor).

| Emotional problems | | | | | | |
|---|---|---|---|---|---|---|
| (I) time | (J) time | Mean Difference (I-J) | Std. Error | Sig.b | 95% Confidence Interval for Differenceb | |
| | | | | | Lower Bound | Upper Bound |
| 1 | 2 | -1.118* | 0.20 | 0.00 | -1.70 | -0.54 |
| | 3 | -0.51 | 0.22 | 0.19 | -1.12 | 0.10 |
| | 4 | -0.39 | 0.22 | 0.80 | -1.02 | 0.24 |
| | 5 | 0.00 | 0.24 | 1.00 | -0.68 | 0.68 |
| 2 | 1 | 1.118* | 0.20 | 0.00 | 0.54 | 1.70 |
| | 3 | .609* | 0.18 | 0.01 | 0.11 | 1.11 |
| | 4 | .727* | 0.18 | 0.00 | 0.21 | 1.25 |
| | 5 | 1.118* | 0.20 | 0.00 | 0.55 | 1.69 |
| 3 | 1 | 0.51 | 0.22 | 0.19 | -0.10 | 1.12 |
| | 2 | -.609* | 0.18 | 0.01 | -1.11 | -0.11 |
| | 4 | 0.12 | 0.16 | 1.00 | -0.34 | 0.57 |
| | 5 | 0.51 | 0.18 | 0.05 | 0.00 | 1.02 |
| 4 | 1 | 0.39 | 0.22 | 0.80 | -0.24 | 1.02 |
| | 2 | -.727* | 0.18 | 0.00 | -1.25 | -0.21 |
| | 3 | -0.12 | 0.16 | 1.00 | -0.57 | 0.34 |
| | 5 | 0.39 | 0.18 | 0.31 | -0.12 | 0.90 |

| | | | | | | |
|---|---|---|---|---|---|---|
| 5 | 1 | 0.00 | 0.24 | 1.00 | -0.68 | 0.68 |
| | 2 | -1.118* | 0.20 | 0.00 | -1.69 | -0.55 |
| | 3 | -0.51 | 0.18 | 0.05 | -1.02 | 0.00 |
| | 4 | -0.39 | 0.18 | 0.31 | -0.90 | 0.12 |
| Conduct Problems | | | | | | |
| (I) time | (J) time | Mean Difference (I-J) | Std. Error | Sig.b | 95% Confidence Interval for Differenceb | |
| | | | | | Lower Bound | Upper Bound |
| 1 | 2 | -.404* | 0.13 | 0.03 | -0.79 | -0.02 |
| | 3 | -0.19 | 0.13 | 1.00 | -0.56 | 0.17 |
| | 4 | -0.13 | 0.13 | 1.00 | -0.50 | 0.24 |
| | 5 | 0.02 | 0.12 | 1.00 | -0.32 | 0.36 |
| 2 | 1 | .404* | 0.13 | 0.03 | 0.02 | 0.79 |
| | 3 | 0.21 | 0.10 | 0.45 | -0.09 | 0.51 |
| | 4 | 0.27 | 0.12 | 0.24 | -0.07 | 0.62 |
| | 5 | .422* | 0.12 | 0.01 | 0.09 | 0.76 |
| 3 | 1 | 0.19 | 0.13 | 1.00 | -0.17 | 0.56 |
| | 2 | -0.21 | 0.10 | 0.45 | -0.51 | 0.09 |
| | 4 | 0.06 | 0.12 | 1.00 | -0.27 | 0.39 |
| | 5 | 0.21 | 0.11 | 0.63 | -0.11 | 0.53 |
| 4 | 1 | 0.13 | 0.13 | 1.00 | -0.24 | 0.50 |
| | 2 | -0.27 | 0.12 | 0.24 | -0.62 | 0.07 |
| | 3 | -0.06 | 0.12 | 1.00 | -0.39 | 0.27 |
| | 5 | 0.15 | 0.12 | 1.00 | -0.18 | 0.48 |
| 5 | 1 | -0.02 | 0.12 | 1.00 | -0.36 | 0.32 |
| | 2 | -.422* | 0.12 | 0.01 | -0.76 | -0.09 |

| | | | | | | |
|---|---|---|---|---|---|---|
| | 3 | -0.21 | 0.11 | 0.63 | -0.53 | 0.11 |
| | 4 | -0.15 | 0.12 | 1.00 | -0.48 | 0.18 |
| Hyperactivity | | | | | | |
| (I) time | (J) time | Mean Difference (I-J) | Std. Error | Sig.b | 95% Confidence Interval for Differenceb | |
| | | | | | Lower Bound | Upper Bound |
| 1 | 2 | -2.137* | 0.18 | 0.00 | -2.64 | -1.63 |
| | 3 | -1.373* | 0.18 | 0.00 | -1.89 | -0.85 |
| | 4 | -1.292* | 0.19 | 0.00 | -1.83 | -0.76 |
| | 5 | -1.043* | 0.21 | 0.00 | -1.64 | -0.45 |
| 2 | 1 | 2.137* | 0.18 | 0.00 | 1.63 | 2.64 |
| | 3 | .764* | 0.15 | 0.00 | 0.34 | 1.19 |
| | 4 | .845* | 0.15 | 0.00 | 0.42 | 1.27 |
| | 5 | 1.093* | 0.18 | 0.00 | 0.58 | 1.61 |
| 3 | 1 | 1.373* | 0.18 | 0.00 | 0.85 | 1.89 |
| | 2 | -.764* | 0.15 | 0.00 | -1.19 | -0.34 |
| | 4 | 0.08 | 0.14 | 1.00 | -0.32 | 0.49 |
| | 5 | 0.33 | 0.17 | 0.52 | -0.15 | 0.81 |
| 4 | 1 | 1.292* | 0.19 | 0.00 | 0.76 | 1.83 |
| | 2 | -.845* | 0.15 | 0.00 | -1.27 | -0.42 |
| | 3 | -0.08 | 0.14 | 1.00 | -0.49 | 0.32 |
| | 5 | 0.25 | 0.17 | 1.00 | -0.23 | 0.72 |
| 5 | 1 | 1.043* | 0.21 | 0.00 | 0.45 | 1.64 |
| | 2 | -1.093* | 0.18 | 0.00 | -1.61 | -0.58 |
| | 3 | -0.33 | 0.17 | 0.52 | -0.81 | 0.15 |
| | 4 | -0.25 | 0.17 | 1.00 | -0.72 | 0.23 |

| | | | | | | |
|---|---|---|---|---|---|---|
| Peer problems | | | | | | |
| (I) time | (J) time | Mean Difference (I-J) | Std. Error | Sig.a | 95% Confidence Interval for Differencea | |
| | | | | | Lower Bound | Upper Bound |
| 1 | 2 | -0.26 | 0.14 | 0.73 | -0.67 | 0.15 |
| | 3 | -0.24 | 0.14 | 1.00 | -0.64 | 0.17 |
| | 4 | -0.22 | 0.15 | 1.00 | -0.66 | 0.21 |
| | 5 | -0.14 | 0.15 | 1.00 | -0.58 | 0.29 |
| 2 | 1 | 0.26 | 0.14 | 0.73 | -0.15 | 0.67 |
| | 3 | 0.03 | 0.12 | 1.00 | -0.31 | 0.36 |
| | 4 | 0.04 | 0.13 | 1.00 | -0.33 | 0.40 |
| | 5 | 0.12 | 0.14 | 1.00 | -0.29 | 0.53 |
| 3 | 1 | 0.24 | 0.14 | 1.00 | -0.17 | 0.64 |
| | 2 | -0.03 | 0.12 | 1.00 | -0.36 | 0.31 |
| | 4 | 0.01 | 0.14 | 1.00 | -0.38 | 0.40 |
| | 5 | 0.09 | 0.14 | 1.00 | -0.30 | 0.48 |
| 4 | 1 | 0.22 | 0.15 | 1.00 | -0.21 | 0.66 |
| | 2 | -0.04 | 0.13 | 1.00 | -0.40 | 0.33 |
| | 3 | -0.01 | 0.14 | 1.00 | -0.40 | 0.38 |
| | 5 | 0.08 | 0.14 | 1.00 | -0.31 | 0.47 |
| 5 | 1 | 0.14 | 0.15 | 1.00 | -0.29 | 0.58 |
| | 2 | -0.12 | 0.14 | 1.00 | -0.53 | 0.29 |
| | 3 | -0.09 | 0.14 | 1.00 | -0.48 | 0.30 |
| | 4 | -0.08 | 0.14 | 1.00 | -0.47 | 0.31 |

| Prosocial Behaviour | | | | | | |
|---|---|---|---|---|---|---|
| (I) time | (J) time | Mean Difference (I-J) | Std. Error | Sig.b | 95% Confidence Interval for Differenceb | |
| | | | | | Lower Bound | Upper Bound |
| 1 | 2 | .640* | 0.16 | 0.00 | 0.18 | 1.10 |
| | 3 | .578* | 0.17 | 0.01 | 0.10 | 1.05 |
| | 4 | .497* | 0.17 | 0.04 | 0.02 | 0.98 |
| | 5 | .683* | 0.17 | 0.00 | 0.20 | 1.17 |
| 2 | 1 | -.640* | 0.16 | 0.00 | -1.10 | -0.18 |
| | 3 | -0.06 | 0.15 | 1.00 | -0.47 | 0.35 |
| | 4 | -0.14 | 0.14 | 1.00 | -0.55 | 0.26 |
| | 5 | 0.04 | 0.16 | 1.00 | -0.40 | 0.49 |
| 3 | 1 | -.578* | 0.17 | 0.01 | -1.05 | -0.10 |
| | 2 | 0.06 | 0.15 | 1.00 | -0.35 | 0.47 |
| | 4 | -0.08 | 0.13 | 1.00 | -0.44 | 0.28 |
| | 5 | 0.11 | 0.15 | 1.00 | -0.33 | 0.54 |
| 4 | 1 | -.497* | 0.17 | 0.04 | -0.98 | -0.02 |
| | 2 | 0.14 | 0.14 | 1.00 | -0.26 | 0.55 |
| | 3 | 0.08 | 0.13 | 1.00 | -0.28 | 0.44 |
| | 5 | 0.19 | 0.14 | 1.00 | -0.22 | 0.60 |
| 5 | 1 | -.683* | 0.17 | 0.00 | -1.17 | -0.20 |
| | 2 | -0.04 | 0.16 | 1.00 | -0.49 | 0.40 |
| | 3 | -0.11 | 0.15 | 1.00 | -0.54 | 0.33 |
| | 4 | -0.19 | 0.14 | 1.00 | -0.60 | 0.22 |

| Behaviour problems | | | | | | |
|---|---|---|---|---|---|---|
| (I) time | (J) time | Mean Difference (I-J) | Std. Error | Sig.b | 95% Confidence Interval for Differenceb | |
| | | | | | Lower Bound | Upper Bound |
| 1 | 2 | -3.919* | 0.42 | 0.00 | -5.12 | -2.72 |
| | 3 | -2.311* | 0.43 | 0.00 | -3.52 | -1.10 |
| | 4 | -2.037* | 0.46 | 0.00 | -3.36 | -0.72 |
| | 5 | -1.17 | 0.51 | 0.22 | -2.61 | 0.27 |
| 2 | 1 | 3.919* | 0.42 | 0.00 | 2.72 | 5.12 |
| | 3 | 1.609* | 0.33 | 0.00 | 0.67 | 2.55 |
| | 4 | 1.882* | 0.37 | 0.00 | 0.83 | 2.93 |
| | 5 | 2.752* | 0.43 | 0.00 | 1.53 | 3.97 |
| 3 | 1 | 2.311* | 0.43 | 0.00 | 1.10 | 3.52 |
| | 2 | -1.609* | 0.33 | 0.00 | -2.55 | -0.67 |
| | 4 | 0.27 | 0.35 | 1.00 | -0.73 | 1.28 |
| | 5 | 1.14 | 0.41 | 0.06 | -0.02 | 2.30 |
| 4 | 1 | 2.037* | 0.46 | 0.00 | 0.72 | 3.36 |
| | 2 | -1.882* | 0.37 | 0.00 | -2.93 | -0.83 |
| | 3 | -0.27 | 0.35 | 1.00 | -1.28 | 0.73 |
| | 5 | 0.87 | 0.41 | 0.36 | -0.30 | 2.04 |
| 5 | 1 | 1.17 | 0.51 | 0.22 | -0.27 | 2.61 |
| | 2 | -2.752* | 0.43 | 0.00 | -3.97 | -1.53 |
| | 3 | -1.14 | 0.41 | 0.06 | -2.30 | 0.02 |
| | 4 | -0.87 | 0.411 | 0.359 | -2.039 | 0.3 |

| | | | | | | |
|---|---|---|---|---|---|---|
| Depression | | | | | | |
| (I) time | (J) time | Mean Difference (I-J) | Std. Error | Sig.b | 95% Confidence Interval for Differenceb | |
| | | | | | Lower Bound | Upper Bound |
| 1 | 2 | -2.938* | 0.34 | 0.00 | -3.92 | -1.96 |
| | 3 | -1.130* | 0.37 | 0.03 | -2.19 | -0.07 |
| | 4 | -1.491* | 0.37 | 0.00 | -2.53 | -0.45 |
| | 5 | -0.73 | 0.40 | 0.67 | -1.85 | 0.40 |
| 2 | 1 | 2.938* | 0.34 | 0.00 | 1.96 | 3.92 |
| | 3 | 1.807* | 0.31 | 0.00 | 0.93 | 2.69 |
| | 4 | 1.447* | 0.33 | 0.00 | 0.51 | 2.38 |
| | 5 | 2.211* | 0.33 | 0.00 | 1.28 | 3.15 |
| 3 | 1 | 1.130* | 0.37 | 0.03 | 0.07 | 2.19 |
| | 2 | -1.807* | 0.31 | 0.00 | -2.69 | -0.93 |
| | 4 | -0.36 | 0.31 | 1.00 | -1.23 | 0.51 |
| | 5 | 0.40 | 0.32 | 1.00 | -0.51 | 1.31 |
| 4 | 1 | 1.491* | 0.37 | 0.00 | 0.45 | 2.53 |
| | 2 | -1.447* | 0.33 | 0.00 | -2.38 | -0.51 |
| | 3 | 0.36 | 0.31 | 1.00 | -0.51 | 1.23 |
| | 5 | 0.76 | 0.29 | 0.10 | -0.07 | 1.60 |
| 5 | 1 | 0.73 | 0.40 | 0.67 | -0.40 | 1.85 |
| | 2 | -2.211* | 0.33 | 0.00 | -3.15 | -1.28 |
| | 3 | -0.40 | 0.32 | 1.00 | -1.31 | 0.51 |

| | | | | | | |
|---|---|---|---|---|---|---|
| | 4 | -0.76 | 0.29 | 0.10 | -1.60 | 0.07 |
| General Anxiety | | | | | | |
| (I) time | (J) time | Mean Difference (I-J) | Std. Error | Sig.b | 95% Confidence Interval for Differenceb | |
| | | | | | Lower Bound | Upper Bound |
| 1 | 2 | -6.067* | 0.72 | 0.00 | -8.12 | -4.02 |
| | 3 | -4.779* | 0.77 | 0.00 | -6.97 | -2.59 |
| | 4 | -4.669* | 0.76 | 0.00 | -6.83 | -2.51 |
| | 5 | -3.294* | 0.79 | 0.00 | -5.56 | -1.03 |
| 2 | 1 | 6.067* | 0.72 | 0.00 | 4.02 | 8.12 |
| | 3 | 1.29 | 0.56 | 0.22 | -0.29 | 2.87 |
| | 4 | 1.40 | 0.63 | 0.29 | -0.40 | 3.20 |
| | 5 | 2.773* | 0.66 | 0.00 | 0.90 | 4.65 |
| 3 | 1 | 4.779* | 0.77 | 0.00 | 2.59 | 6.97 |
| | 2 | -1.29 | 0.56 | 0.22 | -2.87 | 0.29 |
| | 4 | 0.11 | 0.60 | 1.00 | -1.59 | 1.81 |
| | 5 | 1.49 | 0.61 | 0.15 | -0.24 | 3.21 |
| 4 | 1 | 4.669* | 0.76 | 0.00 | 2.51 | 6.83 |
| | 2 | -1.40 | 0.63 | 0.29 | -3.20 | 0.40 |
| | 3 | -0.11 | 0.60 | 1.00 | -1.81 | 1.59 |
| | 5 | 1.37 | 0.60 | 0.23 | -0.33 | 3.08 |
| 5 | 1 | 3.294* | 0.79 | 0.00 | 1.03 | 5.56 |
| | 2 | -2.773* | 0.66 | 0.00 | -4.65 | -0.90 |

| | | | | | | |
|---|---|---|---|---|---|---|
| | 3 | -1.49 | 0.61 | 0.15 | -3.21 | 0.24 |
| | 4 | -1.37 | 0.60 | 0.23 | -3.08 | 0.33 |
| Based on estimated marginal means | | | | | | |
| * The mean difference is significant at the .05 level. | | | | | | |
| b Adjustment for multiple comparisons: Bonferroni. | | | | | | |

Tables S18. Descriptive statistics for individuals in low socioeconomic group (-1SD SES) and for those in higher socioeconomic status group (+1SD SES)

| **(-1SD) on SES** | | | | | | | **(+1SD) on SES** | | | | |
|---|---|---|---|---|---|---|---|---|---|---|---|
| **T1** | | | | | | | **T1** | | | | |
| | n | mean | sd | se | | | | n | mean | sd | se |
| SDQ Emotional problems | 506 | 4.04 | 2.76 | 0.12 | | | SDQ Emotional problems | 1142 | 3.27 | 2.56 | 0.08 |
| SDQ Conduct | 506 | 1.87 | 1.54 | 0.07 | | | SDQ Conduct | 1142 | 1.42 | 1.11 | 0.03 |
| SDQ Hyperactivity | 506 | 3.67 | 2.26 | 0.1 | | | SDQ Hyperactivity | 1142 | 3.14 | 2.14 | 0.06 |
| SDQ Peer problems | 506 | 2.78 | 1.86 | 0.08 | | | SDQ Peer problems | 1142 | 1.67 | 1.67 | 0.05 |
| SDQ Prosocial | 506 | 7.68 | 1.88 | 0.08 | | | SDQ Prosocial | 1142 | 7.63 | 1.84 | 0.05 |
| SDQ Behaviour problems | 506 | 12.35 | 6.11 | 0.27 | | | SDQ Behaviour problems | 1142 | 9.5 | 5.34 | 0.16 |
| Depression | 504 | 5.04 | 4.36 | 0.19 | | | Depression | 1142 | 3.97 | 3.78 | 0.11 |
| General Anxiety | 422 | 8.05 | 8.6 | 0.42 | | | General Anxiety | 1036 | 7.03 | 6.55 | 0.2 |
| Self Harm | 415 | 0.25 | 0.74 | 0.04 | | | Self Harm | 1034 | 1.17 | 0.58 | 0.02 |
| **T2** | | | | | | | **T2** | | | | |
| | n | mean | sd | se | | | | n | mean | sd | se |
| SDQ Emotional problems | 226 | 3.64 | 2.91 | 0.19 | | | SDQ Emotional problems | 628 | 2.78 | 2.42 | 0.1 |
| SDQ Conduct | 226 | 1.73 | 1.31 | 0.09 | | | SDQ Conduct | 628 | 1.37 | 1.13 | 0.05 |
| SDQ Hyperactivity | 226 | 4.34 | 2.36 | 0.16 | | | SDQ Hyperactivity | 628 | 4.21 | 2.24 | 0.09 |
| SDQ Peer problems | 226 | 2.62 | 1.9 | 0.13 | | | SDQ Peer problems | 628 | 1.75 | 1.45 | 0.06 |
| SDQ Prosocial | 226 | 7.01 | 1.87 | 0.12 | | | SDQ Prosocial | 628 | 6.8 | 1.95 | 0.08 |
| SDQ Behaviour problems | 226 | 12.32 | 6.11 | 0.41 | | | SDQ Behaviour problems | 628 | 10.1 | 5.11 | 0.2 |
| Depression | 226 | 5.09 | 4.14 | 0.28 | | | Depression | 628 | 4.01 | 3.73 | 0.15 |
| General Anxiety | 226 | 9.46 | 8.39 | 0.56 | | | General Anxiety | 628 | 7.92 | 6.27 | 0.25 |

| | | | | | | | | | | | |
|---|---|---|---|---|---|---|---|---|---|---|---|
| Self Harm | 225 | 1.08 | 0.4 | 0.03 | | | Self Harm | 622 | 1.04 | 0.26 | 0.01 |
| **T3** | | | | | | | T3 | | | | |
| | n | mean | sd | se | | | | n | mean | sd | se |
| SDQ Emotional problems | 199 | 3.87 | 2.86 | 0.2 | | | SDQ Emotional problems | 527 | 3.02 | 2.56 | 0.11 |
| SDQ Conduct | 199 | 1.88 | 1.41 | 0.1 | | | SDQ Conduct | 527 | 1.39 | 1.13 | 0.05 |
| SDQ Hyperactivity | 199 | 4.5 | 2.45 | 0.17 | | | SDQ Hyperactivity | 527 | 4.12 | 2.24 | 0.1 |
| SDQ Peer problems | 199 | 3.02 | 1.78 | 0.13 | | | SDQ Peer problems | 527 | 1.87 | 1.47 | 0.06 |
| SDQ Prosocial | 199 | 7.01 | 1.96 | 0.14 | | | SDQ Prosocial | 527 | 6.81 | 2.02 | 0.09 |
| SDQ Behaviour problems | 199 | 13.27 | 6.22 | 0.44 | | | SDQ Behaviour problems | 527 | 10.4 | 5.28 | 0.23 |
| Depression | 199 | 5.32 | 4.19 | 0.3 | | | Depression | 527 | 3.98 | 3.77 | 0.16 |
| General Anxiety | 199 | 10.06 | 8.85 | 0.63 | | | General Anxiety | 527 | 8 | 6.65 | 0.29 |
| Self Harm | 194 | 1.08 | 0.36 | 0.03 | | | Self Harm | 522 | 1.06 | 0.32 | 0.01 |
| **T4** | | | | | | | **T4** | | | | |
| | n | mean | sd | se | | | | n | mean | sd | se |
| SDQ Emotional problems | 165 | 3.95 | 2.77 | 0.22 | | | SDQ Emotional problems | 496 | 3.1 | 2.64 | 0.12 |
| SDQ Conduct | 165 | 1.64 | 1.36 | 0.11 | | | SDQ Conduct | 496 | 1.37 | 1.05 | 0.05 |
| SDQ Hyperactivity | 165 | 4.24 | 2.41 | 0.19 | | | SDQ Hyperactivity | 496 | 4.07 | 2.28 | 0.1 |
| SDQ Peer problems | 165 | 3.05 | 1.92 | 0.15 | | | SDQ Peer problems | 496 | 1.93 | 1.46 | 0.07 |
| SDQ Prosocial | 165 | 6.96 | 1.9 | 0.15 | | | SDQ Prosocial | 496 | 6.83 | 1.91 | 0.09 |
| SDQ Behaviour problems | 165 | 12.87 | 6.44 | 0.5 | | | SDQ Behaviour problems | 496 | 10.46 | 5.3 | 0.24 |
| Depression | 165 | 5.28 | 4.42 | 0.34 | | | Depression | 496 | 3.96 | 3.8 | 0.17 |
| General Anxiety | 165 | 10.2 | 9.01 | 0.7 | | | General Anxiety | 496 | 8.55 | 6.71 | 0.3 |
| Self Harm | 163 | 1.15 | 0.58 | 0.05 | | | Self Harm | 490 | 1.04 | 0.24 | 0.01 |
| **T5** | | | | | | | **T5** | | | | |
| | n | mean | sd | se | | | | n | mean | sd | se |
| SDQ Emotional problems | 167 | 3.92 | 2.71 | 0.21 | | | SDQ Emotional problems | 520 | 3.14 | 2.6 | 0.11 |
| SDQ Conduct | 167 | 1.82 | 1.3 | 0.1 | | | SDQ Conduct | 520 | 1.26 | 1.02 | 0.04 |

| | | | | | | | | | | | |
|---|---|---|---|---|---|---|---|---|---|---|---|
| SDQ Hyperactivity | 167 | 4.19 | 2.21 | 0.17 | | | SDQ Hyperactivity | 520 | 3.85 | 2.3 | 0.1 |
| SDQ Peer problems | 167 | 2.84 | 1.78 | 0.14 | | | SDQ Peer problems | 520 | 2.04 | 1.65 | 0.07 |
| SDQ Prosocial | 167 | 6.75 | 2.03 | 0.16 | | | SDQ Prosocial | 520 | 6.79 | 1.88 | 0.08 |
| SDQ Behaviour problems | 167 | 12.77 | 6.06 | 0.47 | | | SDQ Behaviour problems | 520 | 10.28 | 5.39 | 0.24 |
| Depression | 167 | 5.26 | 4.29 | 0.33 | | | Depression | 520 | 3.96 | 3.84 | 0.17 |
| General Anxiety | 167 | 9.75 | 8.72 | 0.68 | | | General Anxiety | 520 | 8.12 | 6.91 | 0.3 |
| Self Harm | 164 | 0.16 | 0.61 | 0.05 | | | Self Harm | 512 | 0.05 | 0.32 | 0.01 |

Tables S19. MANOVA results for in low socioeconomic group (-1SD SES) compared to the rest of the sample

| | **Time** | | | **SES** | | | **Time * SES** | | |
|---|---|---|---|---|---|---|---|---|---|
| | **F** | **p** | **Partial Eta squared** | **F** | **p** | **Partial Eta squared** | **F** | **p** | **Partial Eta squared** |
| SDQ Emotional problems | 3.983 | 0.001 | 0.004 | 2.226 | 0.136 | 0.003 | 0.206 | 0.935 | 0 |
| SDQ Conduct | 0.877 | 0.472 | 0.001 | 11.775 | 0.001 | 0.013 | 0.823 | 0.504 | 0.001 |
| SDQ Hyperactivity | 20.772 | 0.001 | 0.023 | 6.922 | 0.009 | 0.008 | 1.582 | 0.572 | 0.001 |
| SDQ Peer problems | 2.666 | 0.035 | 0.003 | 27.051 | 0.001 | 0.03 | 0.952 | 0.428 | 0.001 |
| SDQ Prosocial | 18.286 | 0.001 | 0.02 | 6.155 | 0.013 | 0.007 | 2.008 | 0.091 | 0.002 |
| SDQ Behaviour problems | 2.516 | 0.04 | 0.003 | 13.887 | 0 | 0.015 | 0.049 | 0.993 | 0 |
| Depression | 1.095 | 0.357 | 0.001 | 7.889 | 0.005 | 0.009 | 1.141 | 0.335 | 0.001 |

Tables S20. Sample characteristics across data collection points

| | TEDS 21 phase 1 twin study | Covid phase 1 twin study sample for May 2020 dataset | Covid phase 1 twin study, final sample | Covid phase 2 twin study, final sample | Covid phase 3 twin study, final sample | National statistic (from TRHG paper) |
|---|---|---|---|---|---|---|
| N: full pairs (both twins with data) | 4084 | 1182 | 1493 | 1236 | 1111 | - |
| N: unpaired (one twin with data) | 1562 | 1797 | 1909 | 1569 | 1454 | - |
| N: total pairs with some data | 5646 | 2979 | 3402 | 2805 | 2565 | - |
| % white | 93.6% | 93.6% | 94.0% | 94.4% | 94.4% | 93% |

| | | | | | | |
|---|---|---|---|---|---|---|
| % twins with 3+ full A-levels (rcqaln1/2>=3), for individual twins not pairs | 53.3% | 57.5% | 57.1% | 58.3% | 58.7% | 42.1% |
| % mothers with A-levels or higher (amohqual >= 4) | 43.4% | 46.4% | 46.7% | 47.6% | 47.5% | 35% |
| % fathers with A-levels or higher (afahqual >= 4) | 51.1% | 54.3% | 54.9% | 55.0% | 55.6% | 47% |
| % mother employed | 46.5% | 46.6% | 46.7% | 46.5% | 46.4% | 50% |
| % father employed | 93.6% | 94.1% | 94.1% | 93.8% | 94.2% | 91% |
| % female (sex1/2=0) for individual twins not pairs | 59.1% | 63.3% | 62.8% | 64.6% | 65.5% | - |
| % MZ ( | 35.0% | 36.6% | 36.2% | 36.7% | 37.8% | 33% |
| % going on to university | 54.7% | 58.5% | 58.1% | 59.0% | 59.4% | - |
| GCSE results from the TEDS 16 year GCSE dataset: | | | | | | |
| Mean grade in all GCSEs * | 9.03 | 9.15 | 9.15 | 9.17 | 9.16 | - |
| Mean grade in GCSE core subjects (English, maths and science) * | 9.06 | 9.18 | 9.19 | 9.21 | 9.20 | - |
| Mean number of GCSE results (all grades) | 9.4 | 9.5 | 9.5 | 9.6 | 9.6 | - |
| Mean number of GCSE passes at grades A* to C | 8.5 | 8.7 | 8.7 | 8.8 | 8.8 | - |
| % with 5 or more GCSE passes at grades A* to C | 87.8% | 89.3% | 90.1% | 89.9% | 89.7% | - |
| | * GCSE grades are coded 11=A*, 10=A, 9=B, etc | | | | | |

### Figures

Figure S1. Patterns of individual variability across timepoints for all mental health measures. Individual trajectories are presented as coloured lines and the average mean trajectory as a black line.

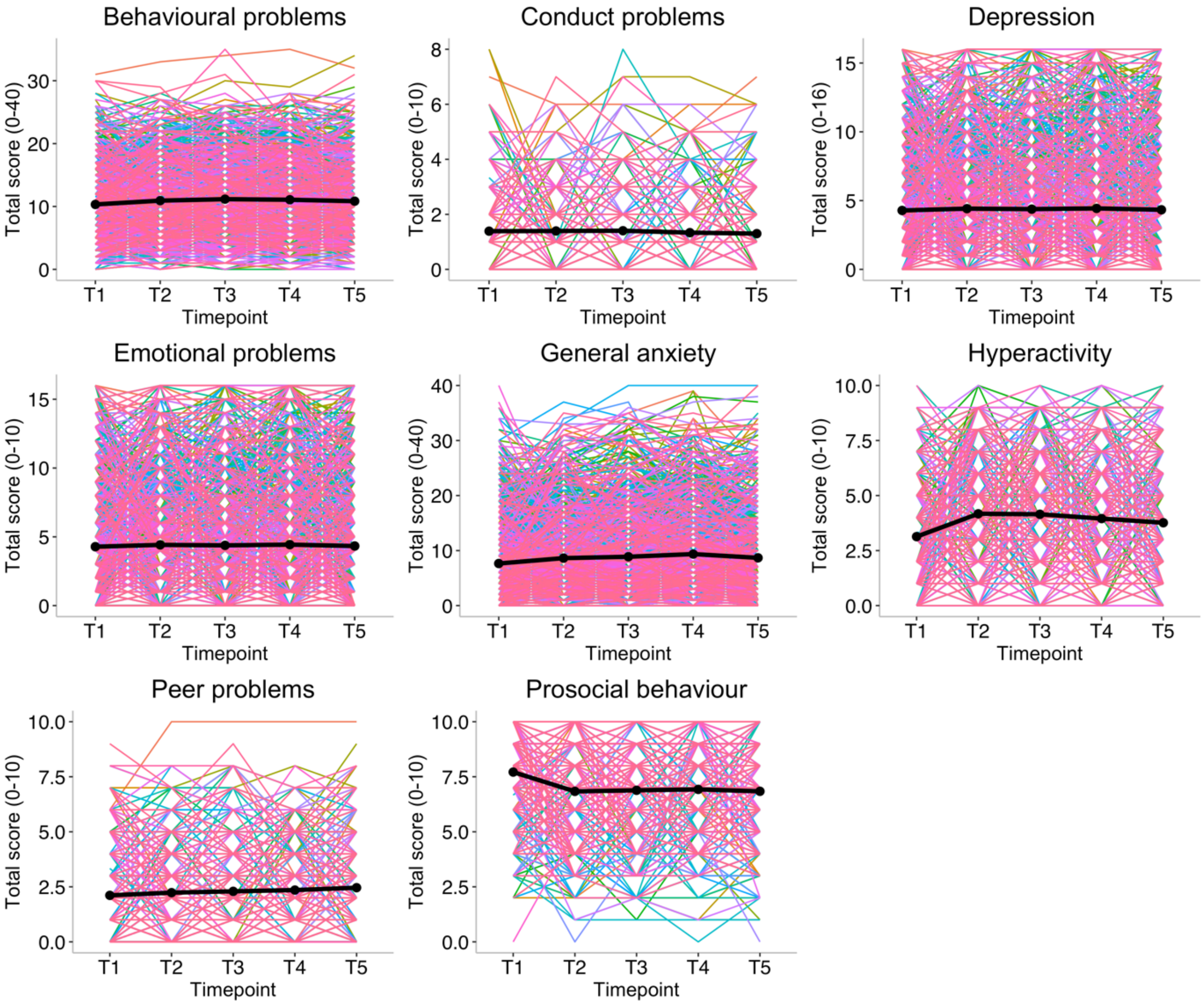

Figure S2. Twin model-fitting results illustrating the unstandardised variance components

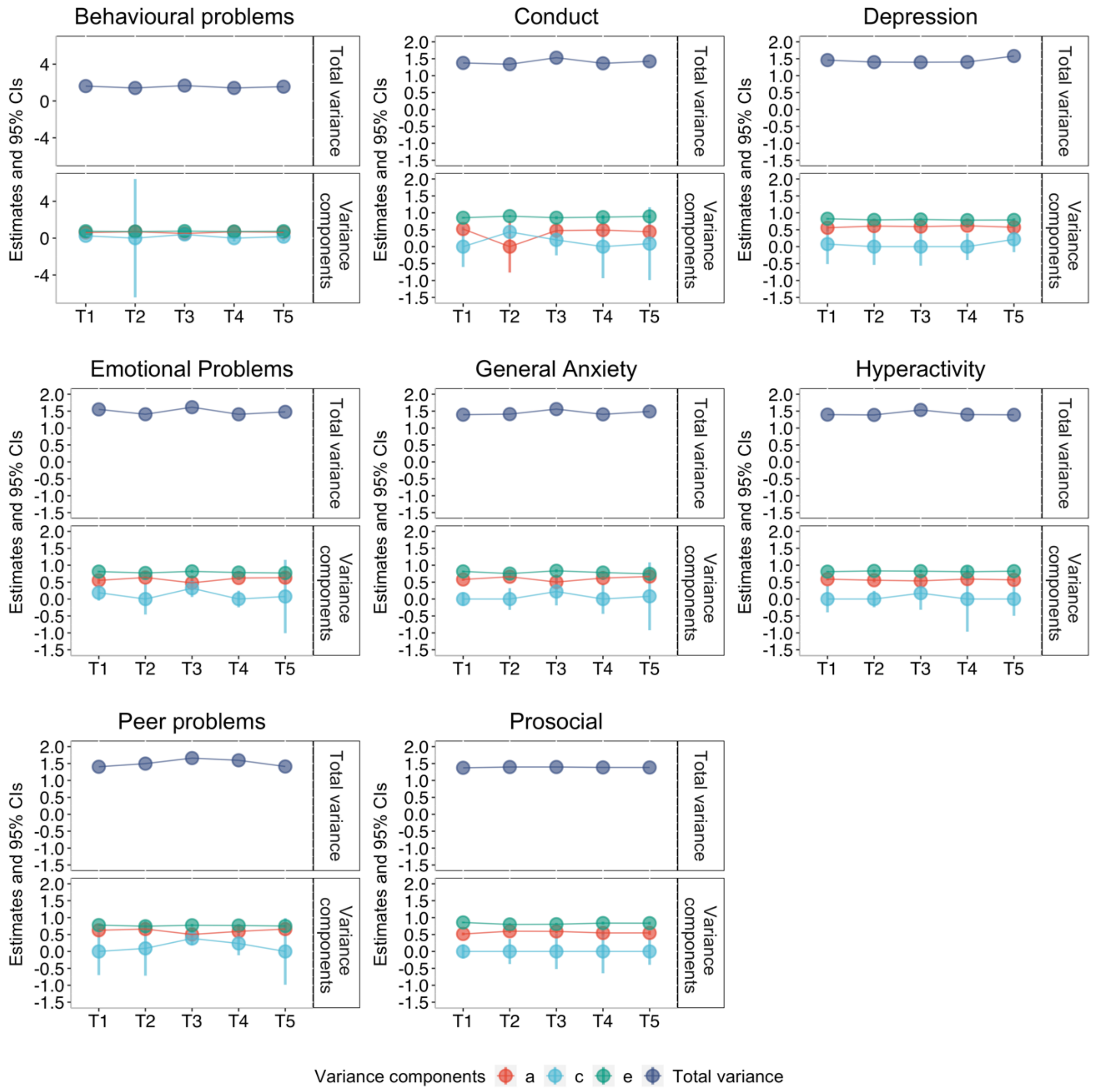

Figure S3. Patterns of individual variability across timepoints for all mental health measures separated by -/+ 1 SD on genomic p-factor. Individual trajectories are presented in coloured lines and average mean trajectory in black line.

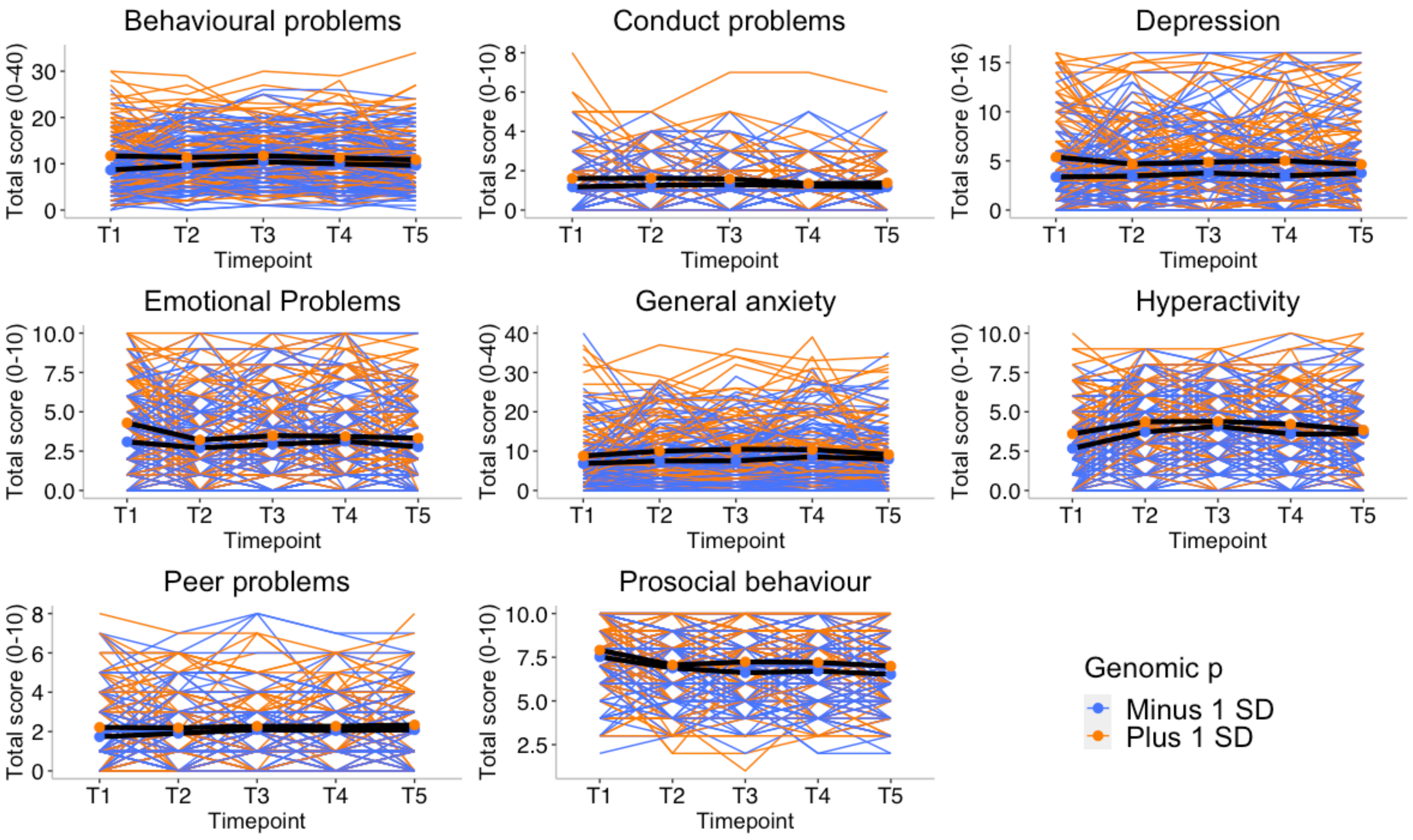

Figure S4. Patterns of individual variability across timepoints for all mental health measures separated by -/+ 1 SD on cross-disorder polygenic score (GPS). Individual trajectories are presented in coloured lines and average mean trajectory in black line.

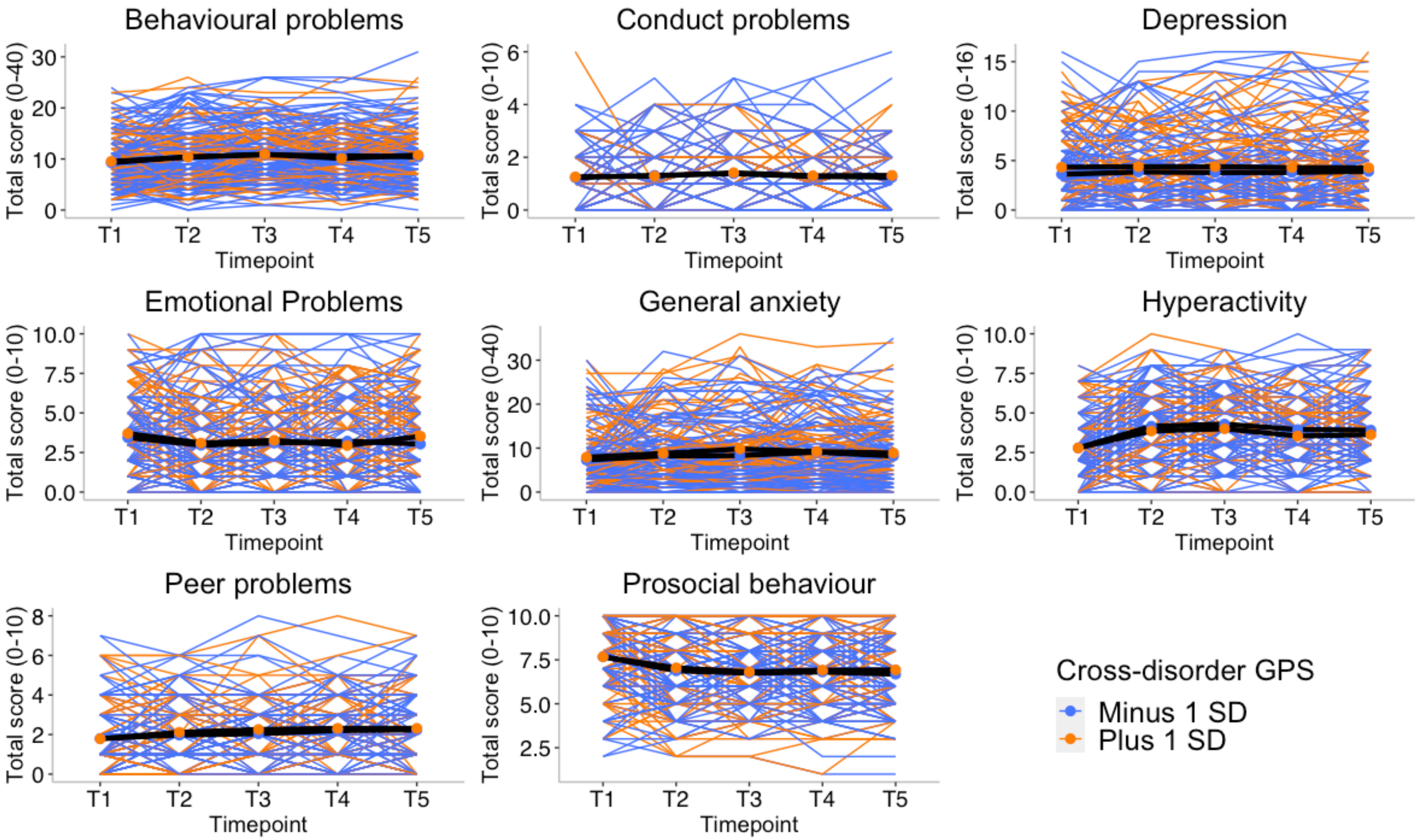

Figure S5. Patterns of individual variability across timepoints for all mental health measures separated by -/+ 1 SD on depression polygenic score (GPS). Individual trajectories are presented in coloured lines and average mean trajectory in black line.

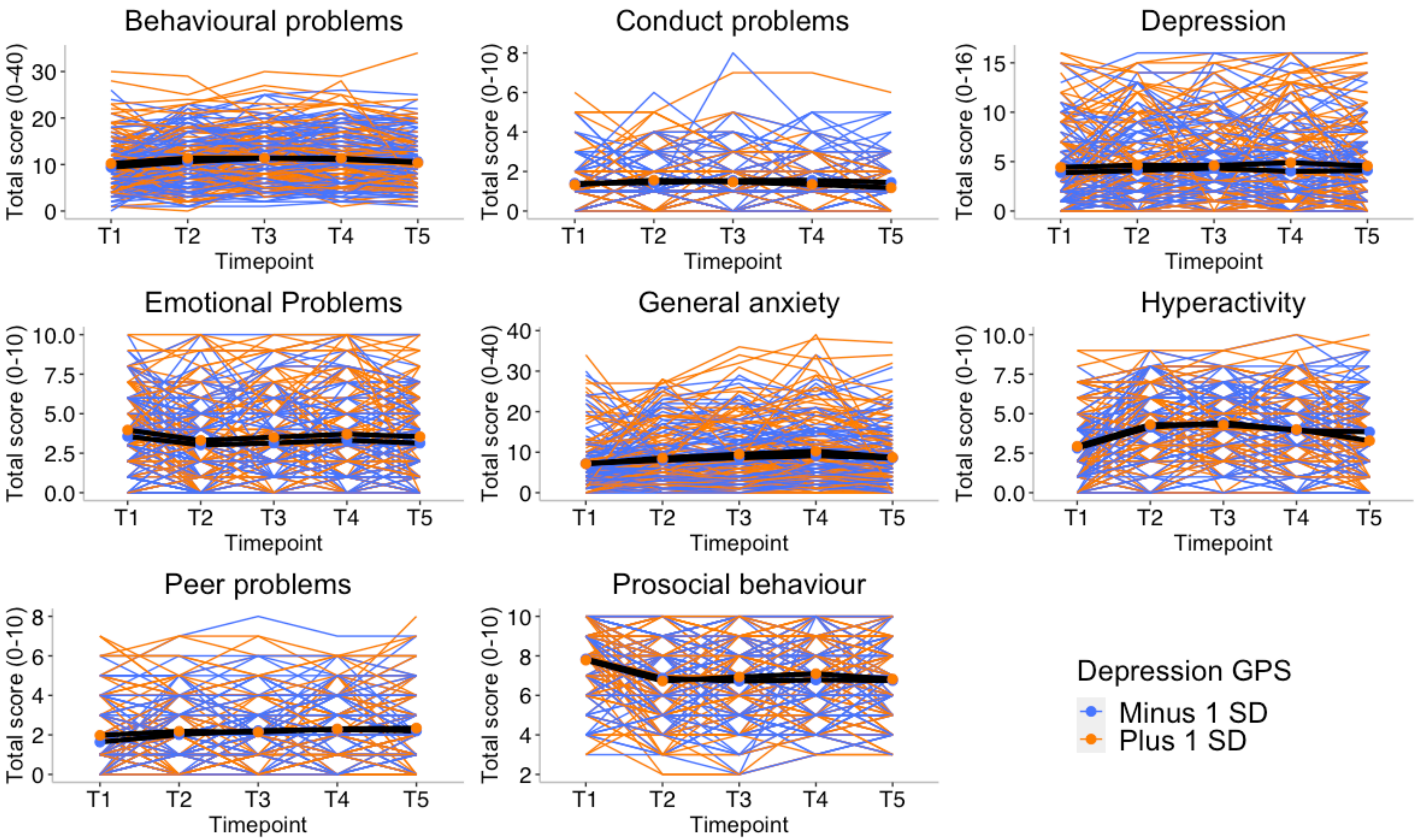

Figure S6. Patterns of individual variability across timepoints for all mental health measures separated by -/+ 1 SD on anxiety polygenic score (GPS). Individual trajectories are presented in coloured lines and average mean trajectory in black line.

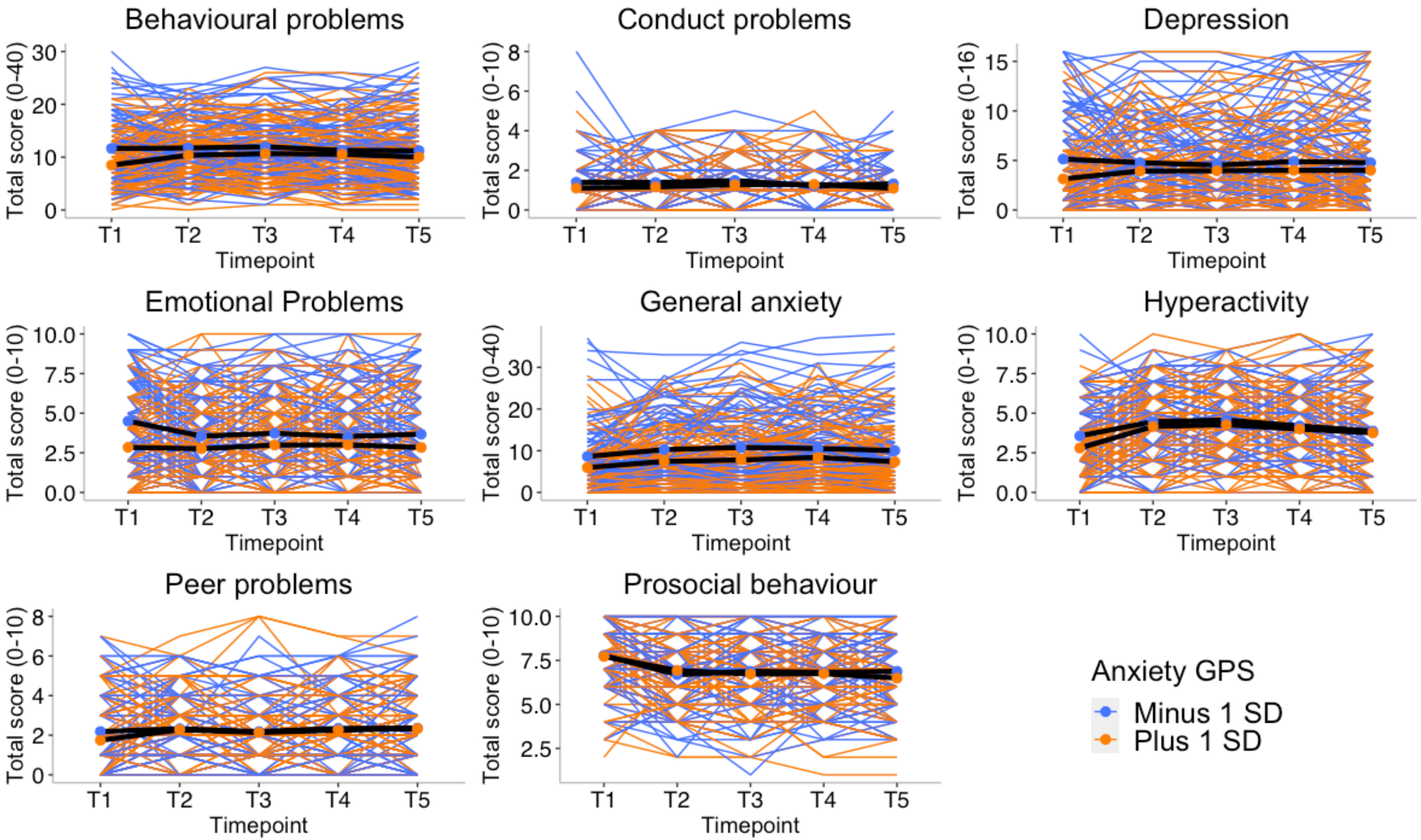

Figure S7. Patterns of individual variability across timepoints for all mental health measures separated by -/+ 1 SD on risk tolerance polygenic score (GPS). Individual trajectories are presented in coloured lines and average mean trajectory in black line.

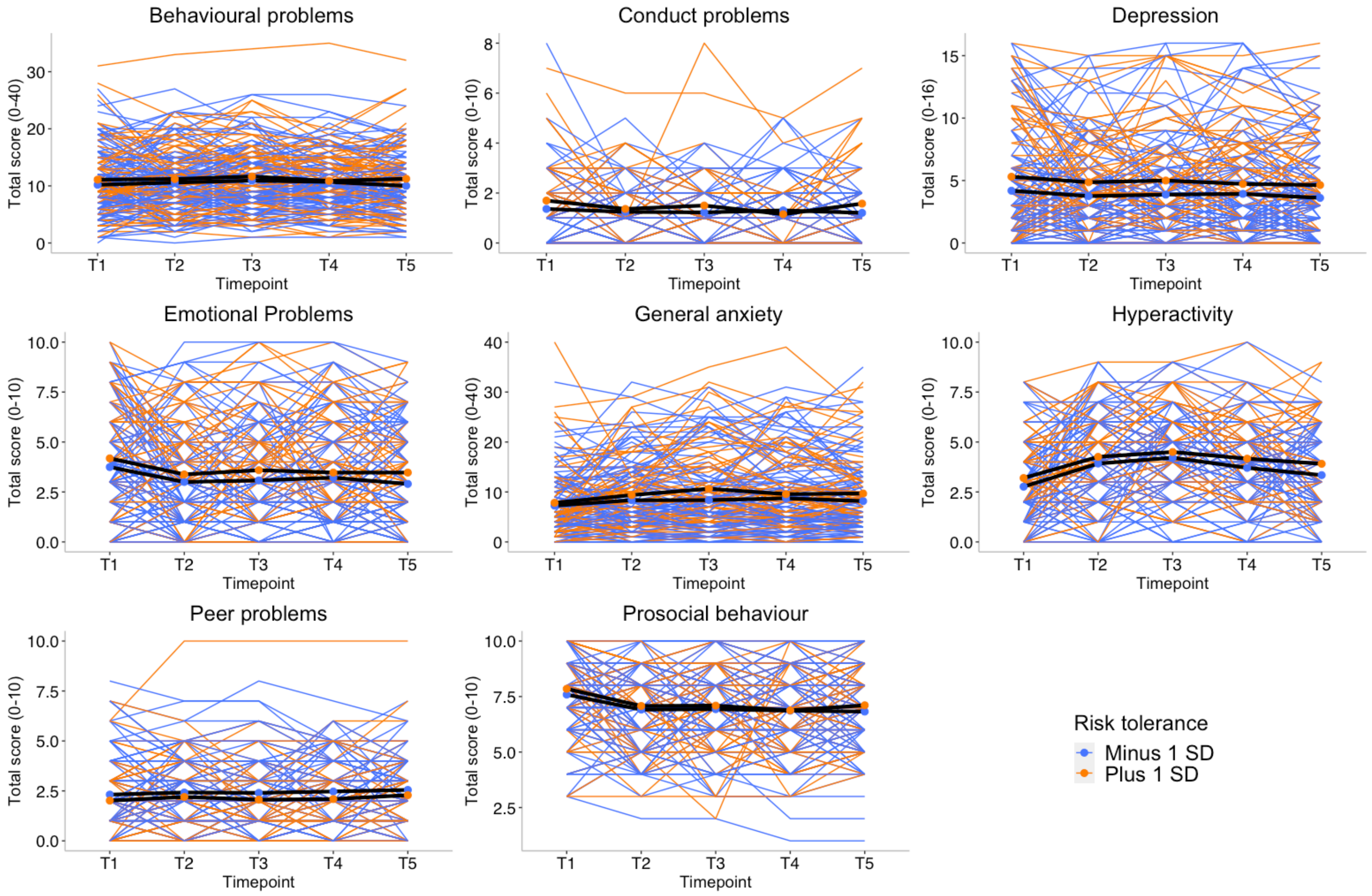

Figure S8. Patterns of individual variability across timepoints for all mental health measures separated by -/+ 1 SD on educational attainment (EA) polygenic score (GPS). Individual trajectories are presented in coloured lines and average mean trajectory in black line.

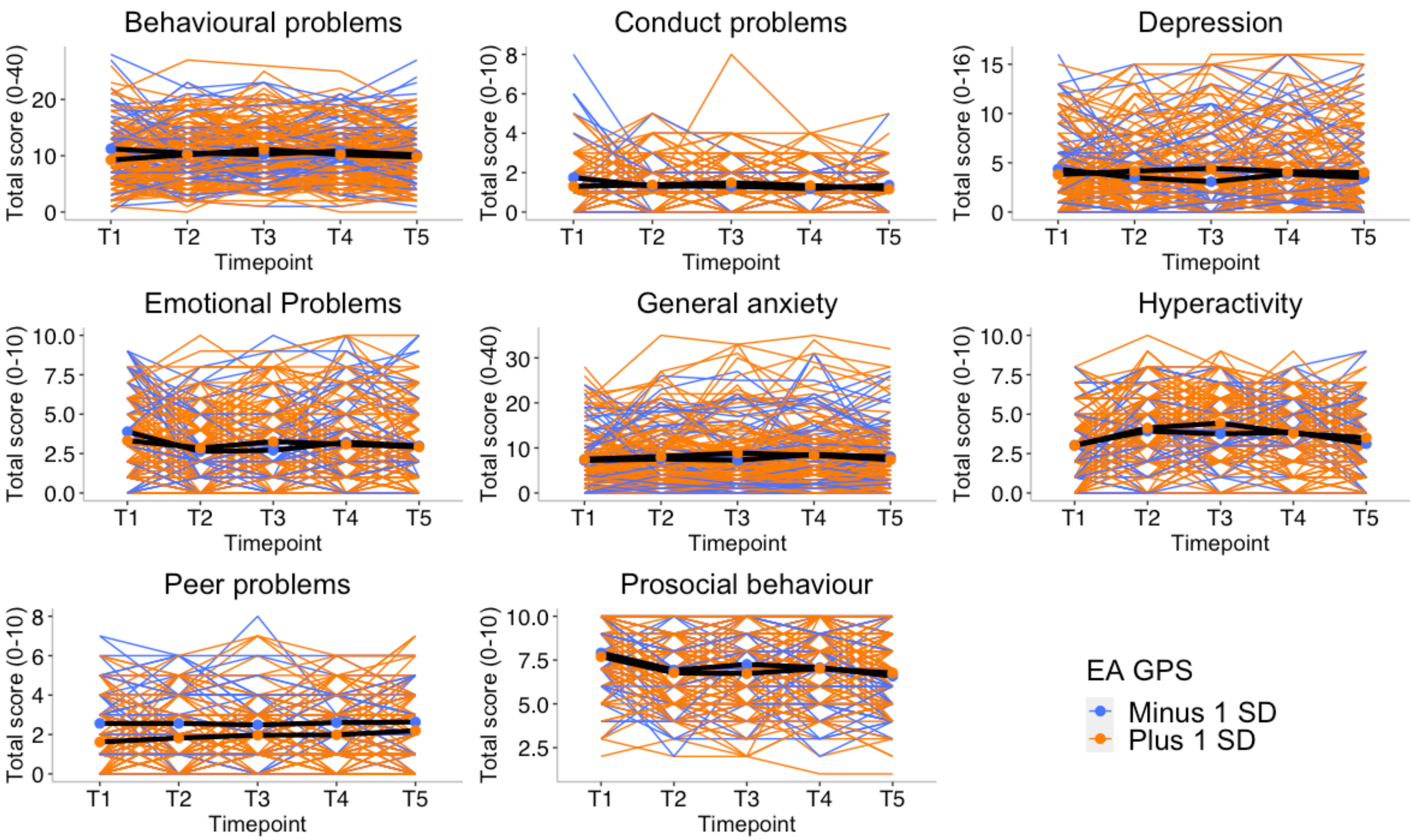

Figure S9. Patterns of individual variability across timepoints for all mental health measures separated by -/+ 1 SD on family socioeconomic status (collected at first contact). Individual trajectories are presented in coloured lines and average mean trajectory in black line.

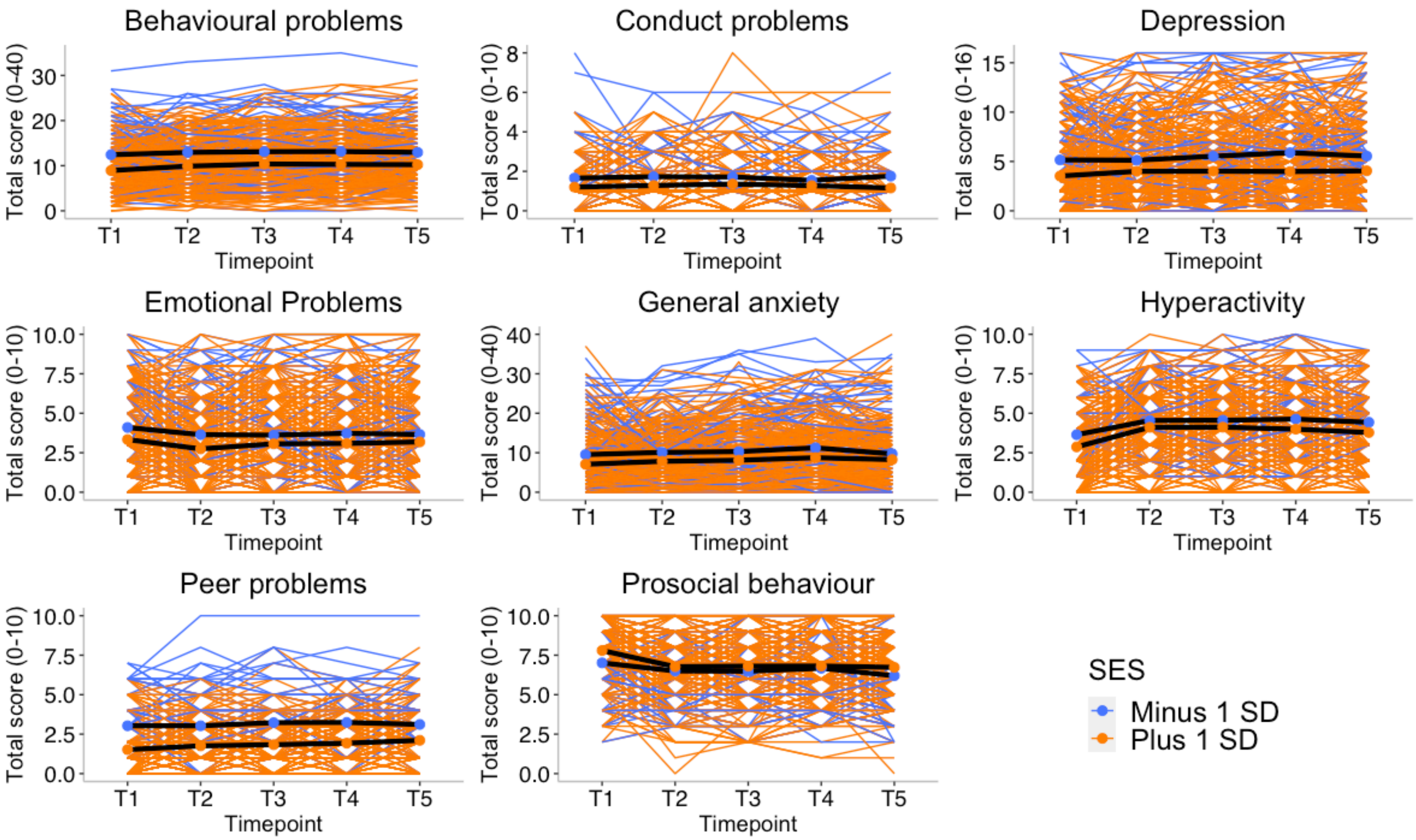

Figure S10. Patterns of individual variability across timepoints for all mental health measures separated by those having children and those who do not. Individual trajectories are presented in coloured lines and average mean trajectory in black line.

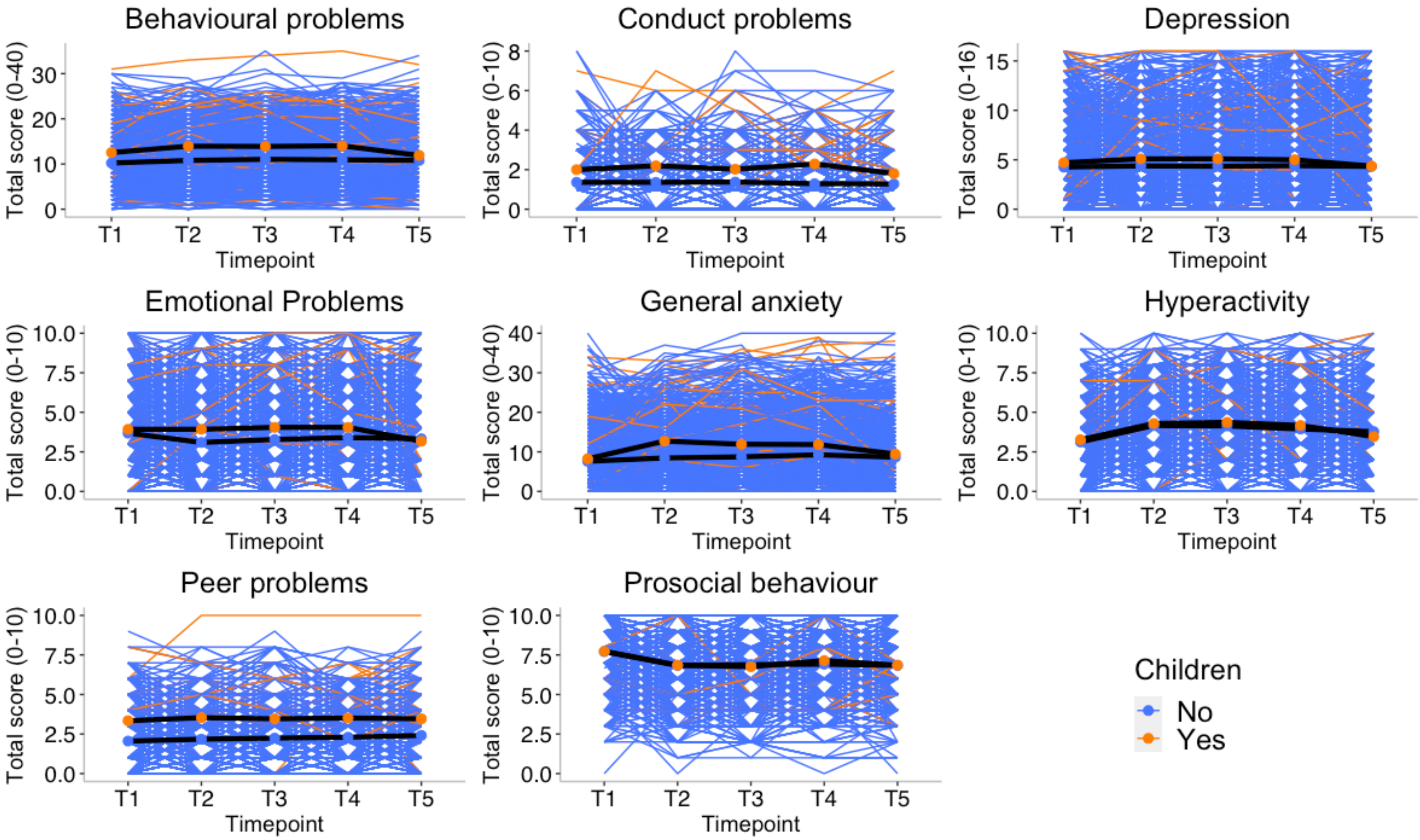

Figure S11. Patterns of individual variability across timepoints for all mental health measures separated by those having access to garden/green space from those who did not. Individual trajectories are presented in coloured lines and average mean trajectory in black line.

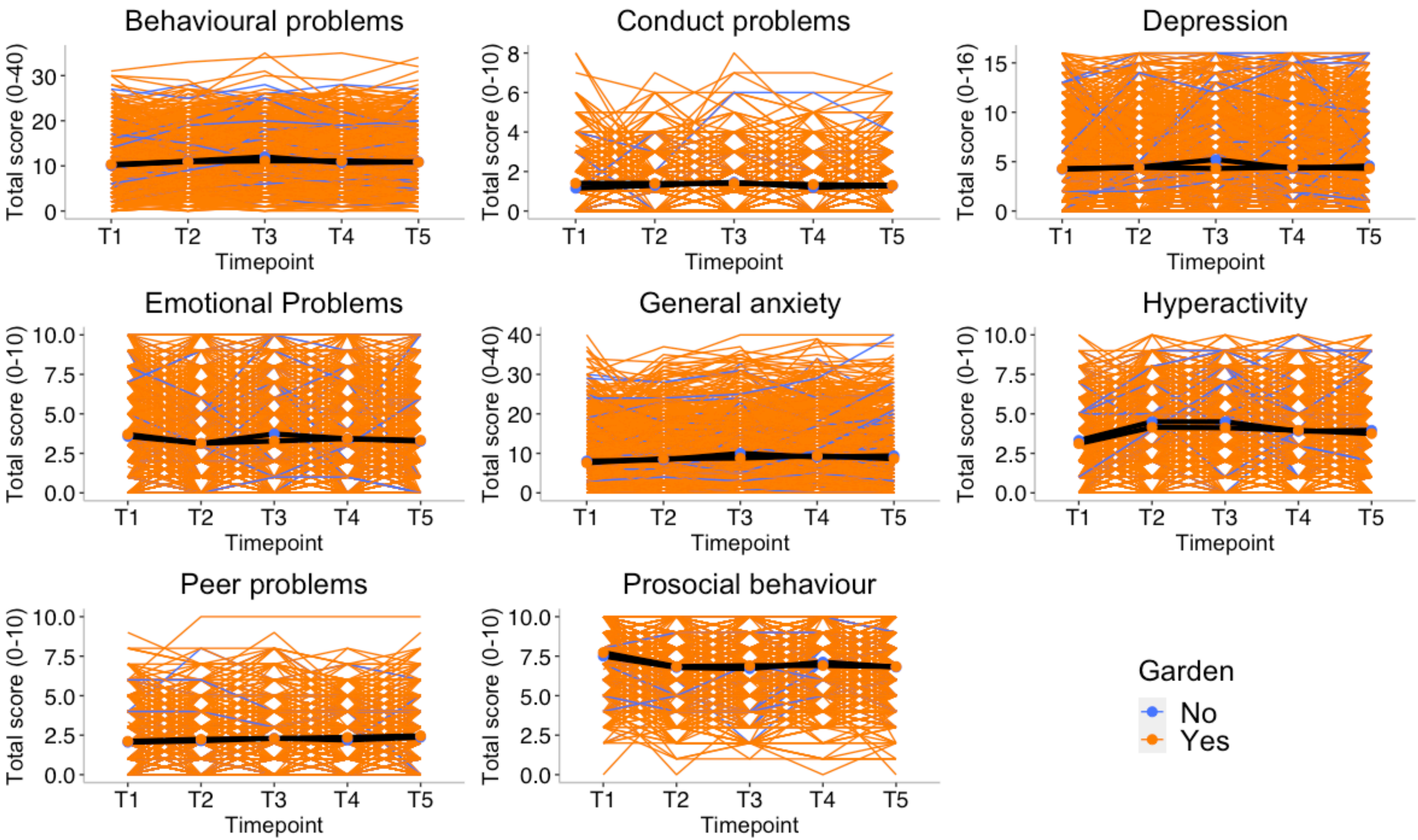

Figure S12. Patterns of individual variability across timepoints for all mental health measures separated by those whose family member lost a job during the lockdown compared to those who did not. Individual trajectories are presented in coloured lines and average mean trajectory in black line.

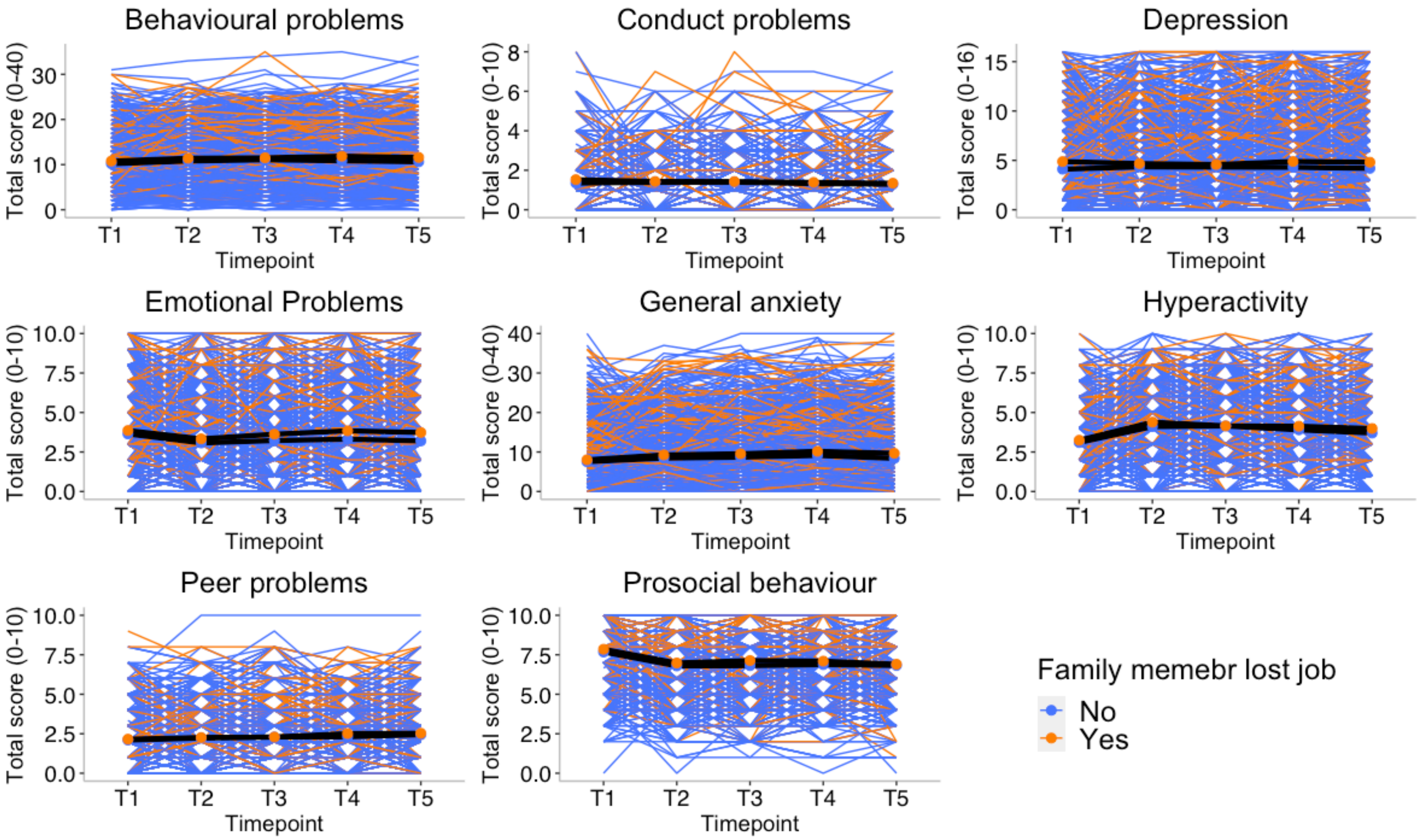

Figure S13. Patterns of individual variability across timepoints for all mental health measures separated by worrying about paying for food at any point during the pandemic from those who did not.  Individual trajectories are presented in coloured lines and average mean trajectory in black line.

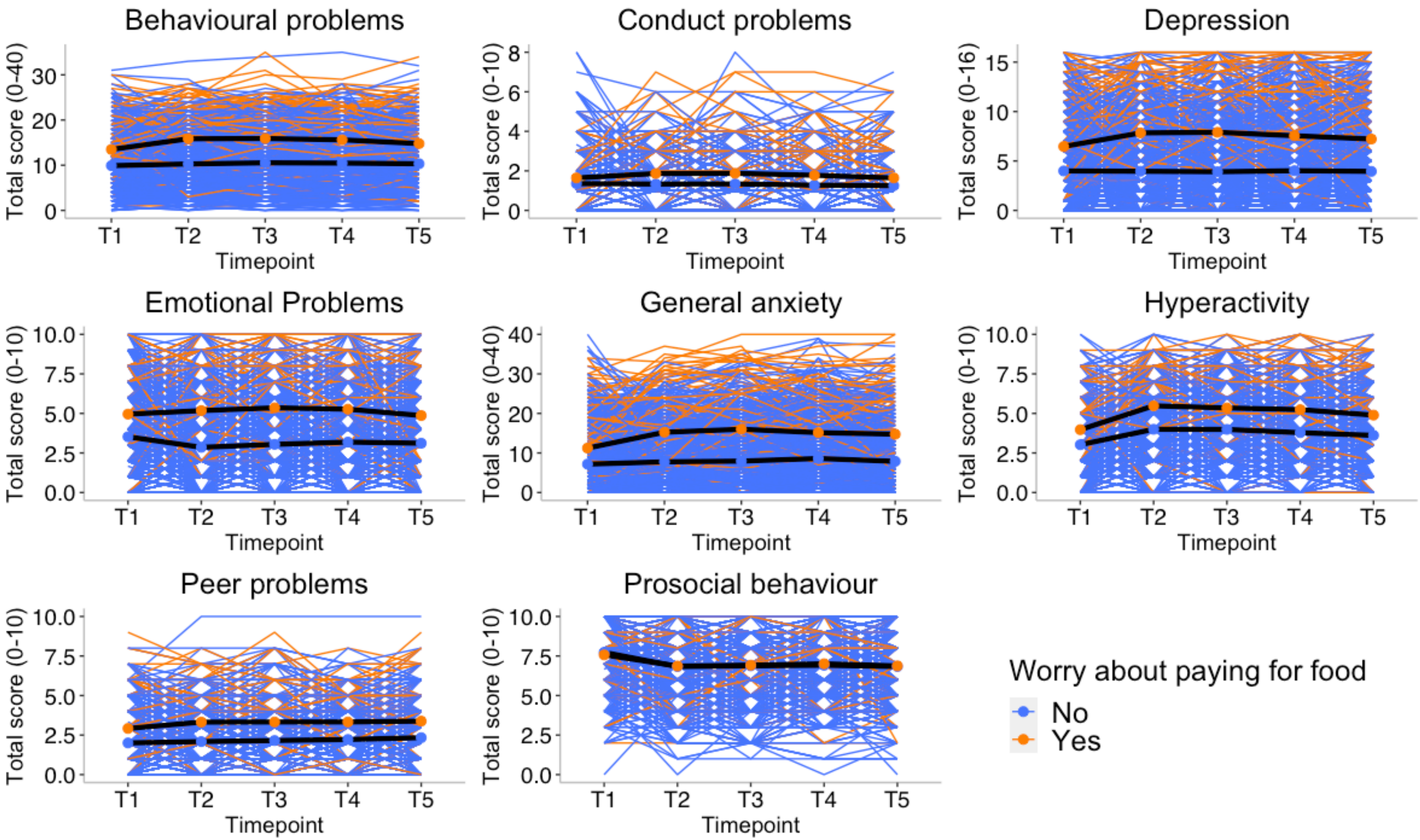

Figure S14. Patterns of individual variability across timepoints for all mental health measures separated by having COVID-19 diagnoses or symptoms at any point during the pandemic from those who did not.  Individual trajectories are presented in coloured lines and average mean trajectory in black line.

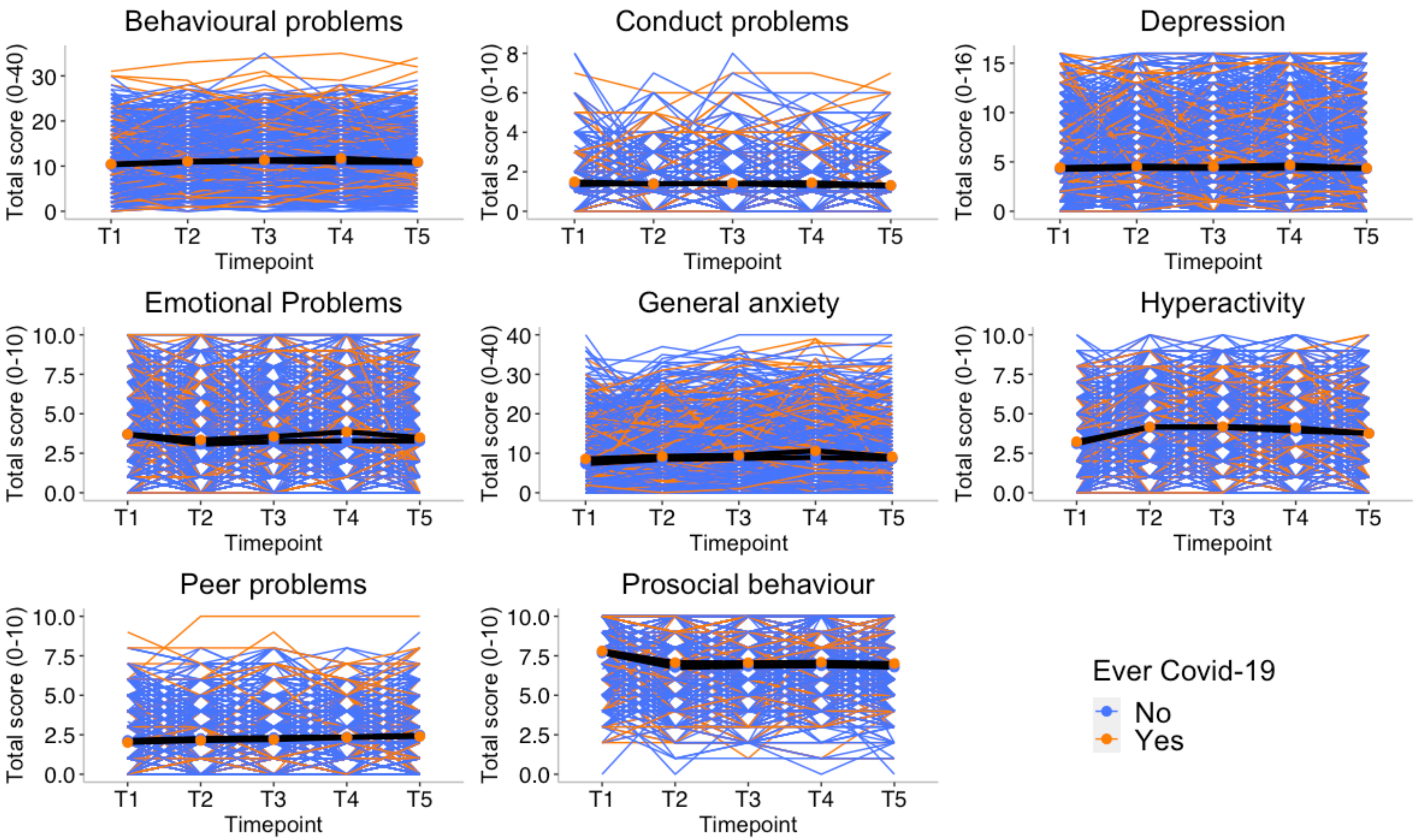

Figure S15. Patterns of individual variability across timepoints for all mental health measures separated by those with possible long COVID (symptom lasting longer than 1 months) from those without. Individual trajectories are presented in coloured lines and average mean trajectory in black line.

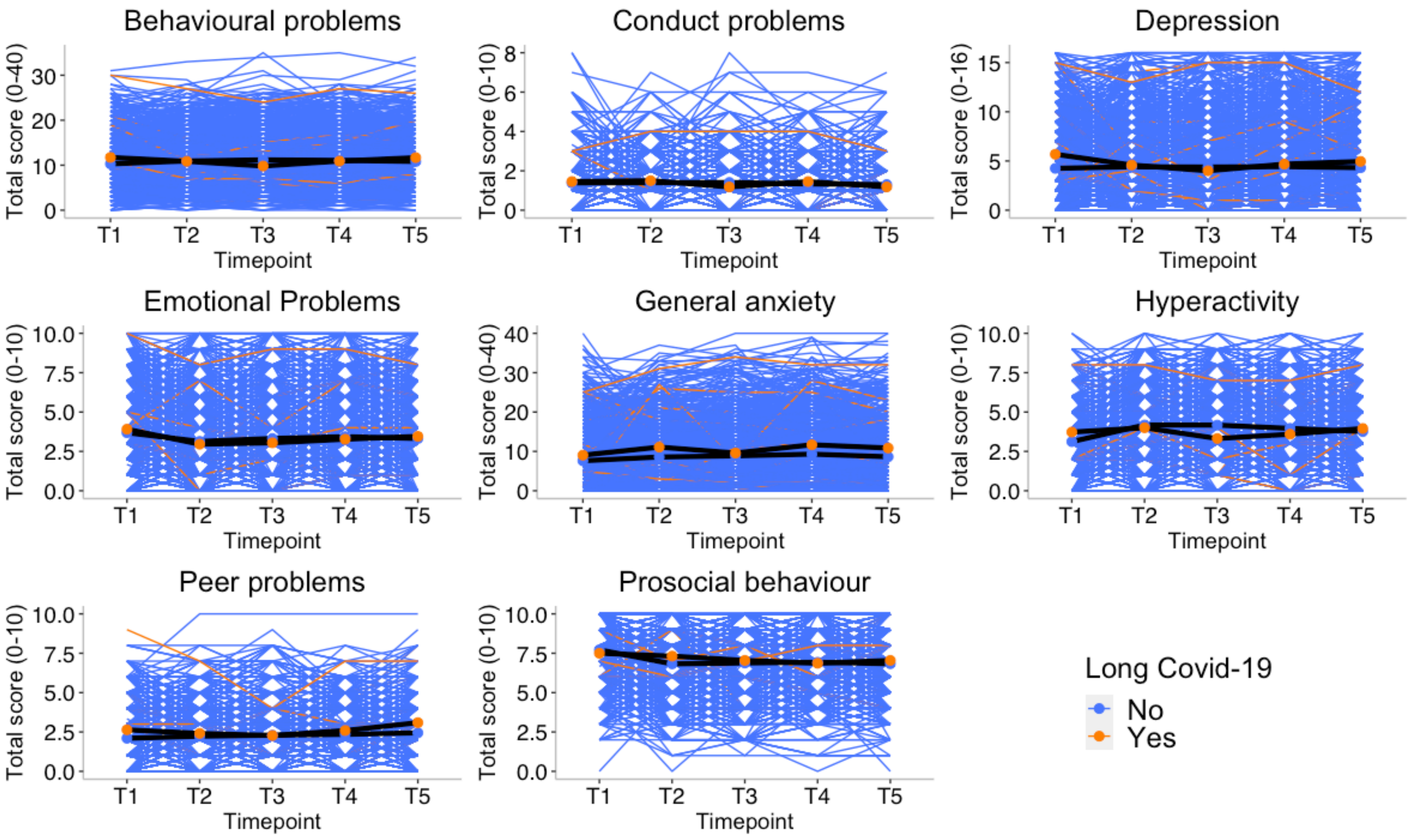

Figure S16. Correlations between mental health symptoms with worries and family effect.

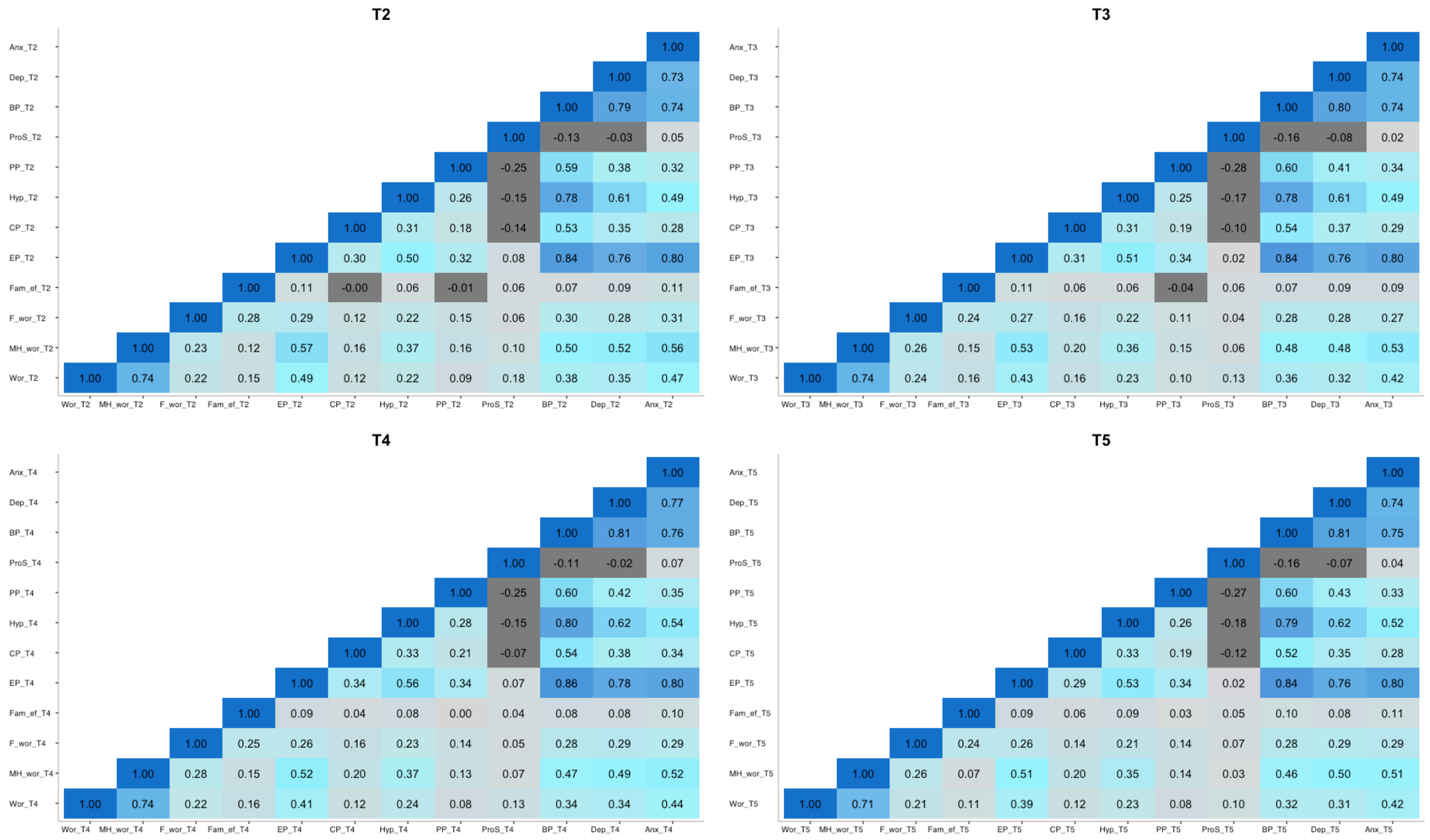

Note: Anx= General anxiety; Dep=depression; BP= Behavioural Problems; ProS=Prosocial behaviour; PP= Peer Problems; CP= conduct problems; EP= emotional problems; Fam_ef= Family Effect (sum score of family effects, losing a job, getting infected, being hospitalised, lost family member); F_wor= financial worries; MH_wor= mental health worries; Wor= worries (composite score of mental health worries, financial worries and pandemic related worries).
